# Estimating the Prevalence of Generative AI Use in Medical School Application Essays

**DOI:** 10.1101/2024.10.21.24315868

**Authors:** Nicholas C. Spies, Valerie S. Ratts, Ian S. Hagemann

**Author notes:** Correspondence to: Ian S. Hagemann, Department of Pathology and Immunology, Mailstop 8118-04-003, 425 S. Euclid Ave., St. Louis, MO 63110.

## Abstract

**Background:** Generative artificial intelligence (AI) tools became widely available to the public in November 2022. The extent to which these tools are being used by aspiring medical school applicants during the admissions process is unknown.

**Methods:** We retrospectively analyzed 6,000 essays submitted to a U.S. medical school in 2021– 2022 (baseline, before wide availability of AI) and in 2023–2024 (test year) to estimate the prevalence of AI use and its relation to other application data. We used GPTZero, a commercially available detection tool, to generate a metric for the likelihood that each essay was human-generated, P_human_, ranging from 0 (entirely AI) to 1 (entirely human).

**Results:** Fully human-generated negative controls demonstrated a median P_human_ of 0.93, while AI-generated positive controls demonstrated a median P_human_ of 0.01. Personal Comments essays submitted in the ‘23–‘24 cycle had a median human-generated score of 0.77 (95% confidence interval 0.76–0.78), versus 0.83 (95% CI 0.82–0.85) during the ‘21–‘22 cycle. Approximately 12.3 and 2.7% of essays were evaluated as having P_human_ < 0.5 in the test and baseline year, respectively. Secondary essays demonstrated lower P_human_ than Personal Comments essays, suggesting more AI use. In multivariate analysis, younger age, visa requirement, and higher GPA were significantly associated with lower P_human_. No differences were observed in gender, MCAT score, undergraduate major, or socioeconomic status. P_human_ was not predictive of admissions outcomes in uni- or multivariate analyses.

**Conclusions:** An AI detection algorithm estimated significantly increased use of generative AI in 2023-2024 medical school admission applications, as compared to the 2021-2022 baseline. Estimated AI use demonstrated no significant differences in admissions decisions. While these results provide information about the applicant pool as a whole, AI detection is imperfect. We recommended exercising caution before deploying any AI detection tools on individual applications in live admissions cycles.

**Description:** Medical school applicants increased their use of generative AI to write application essays in the most recent admissions cycle, but this use did not confer an admissions advantage.

## Introduction

Generative artificial intelligence (AI) has exploded in popularity in recent years, with “chatbots” driven by large language models (LLMs) providing a means to rapidly accomplish text-oriented tasks. These tools have a myriad of potential applications in medical education, including information retrieval^1^ and the generation of practice questions^2^, clinical vignettes, and simulations^3^. However, they also threaten the pedagogical use of writing assignments by allowing students to produce responses with minimal effort or understanding^4^. AI’s role in medical education remains a largely unexplored and rapidly evolving field.

One setting where AI chatbots may have far-reaching implications is the admissions process. Medical school application essays are intended to give admissions officers information about the applicant’s interests, experiences, attributes and motivations. Moreover, the quality of the essays is often used as an approximation of the work quality that may be expected of the student if they ultimately matriculate. In November 2022, the first widely available chatbot, ChatGPT 3.0, was released to the public, introducing the possibility that medical school applicants could use this tool either to write their essays outright, to provide a first draft, or to do final editing.

There is no consensus on the extent to which AI use by medical applicants is acceptable. Applicants have long relied upon aids such as spelling and grammar checkers, formal editing services, and feedback from friends, family and advisors. Receiving help from a chatbot could level the playing field for applicants with lower writing skills or less access to other writing aids. On the other hand, AI-generated essays give less insight into the applicant’s authentic self and work performance and are therefore less fit for purpose as components of a school application. For the 2023–24 application cycle, the American Medical College Application Service (AMCAS) adopted a policy^5^ that essays must “not be written, in part or in whole, by another author and… not [be] the product of artificial intelligence.” While applicants attested to this certification statement, it is uncertain to what degree they complied.

The popularity, potential impact, and ethical considerations of AI-driven chatbots have motivated the development of methods by which AI-generated text can be distinguished from its human-generated counterparts. These methods often rely on the extent to which each word is predictable based on those that came before it, known as “perplexity”, and the degree of variation in sentence length and structure, known as “burstiness”^6^. Human-generated writing tends to have higher perplexity and burstiness than the outputs of current LLMs, representing the greater variety and spontaneity of natural human expression.

To explore the extent to which LLMs are possibly being used in medical school applications, we conducted a retrospective study of AMCAS writing samples from before and after the surge in popularity of AI-driven chatbots in 2022. We analyzed these essays using a commercial AI detection platform, GPTZero^6^, and compared the detector’s outputs across relevant demographic and essay-related metadata.

## Methods

### Data retrieval

We conducted this study at a U.S. medical school that participates in the AMCAS Data Exchange Service. Applicants were included in the study if they selected the school as part of their AMCAS application, regardless of whether they completed a secondary application. We retrieved data from the school’s applicant tracking system in aggregate form, identified only by AMCAS ID, a coded eight-digit identifier. An honest broker who was not a member of the study team provided the data to the team to eliminate the possibility of re-identification. We coded applications as “complete” if the student submitted their AMCAS file, letters of recommendation meeting the school’s requirement, and their secondary application. We retrieved the following data from the AMCAS application: AMCAS ID, age, self-reported gender, program type (MD vs. MD/PhD), visa status, socioeconomic status indicator, undergraduate major(s), undergraduate grade point average (GPA), highest Medical College Admission Test (MCAT) three-digit score, and the applicant’s personal statement. We coded socioeconomic status using the AMCAS indicator EO1/EO2 (indicating that neither parent has a college degree or an executive, managerial or professional occupation) versus other. We manually coded each applicant’s undergraduate major(s) as follows: science, technology, engineering or math (STEM), non-STEM, or both (possible only for those declaring multiple majors; **Supplemental Table 1**). We retrieved the following data from the school-specific secondary application: a supplemental essay in which the student is asked to describe a time in life when they were unsuccessful (“Failure” essay, required as part of the secondary), a free-text box where they can enter any additional information they wish to share (“Anything Else” essay, optional), date of application submission, and the student’s admission decision (accepted vs. rejected). We considered placement on the Alternate List as rejection for this study.

### Estimating the probability that essays were generated using AI

Given the reported difficulties in accurately discriminating human from AI-generated text^7^, we first performed a preliminary analysis on negative and positive controls. We defined continuation criteria for the real-world analysis as detecting a difference in predicted probability of greater than 0.5 with 95% confidence (e.g., a median human probability of >0.75 in negative controls and <0.25 in positive controls). For negative controls, we subjected five essays known to be completely human-generated, obtained from the authors’ personal files, to the workflow described below. For positive controls, we submitted essay prompts to GPT 3.5 (OpenAI) and Claude 3 Sonnet (Anthropic) through their respective online user interfaces. The responses from each chatbot were completely generated by the LLM, with no subsequent human editing. The controls are available as **Supplemental Table 2**.

We used GPTZero API v2.0.0, a commercially available web-based service, to detect AI use. We sent the entirety of each writing sample to be analyzed to the API, which returns a vector of three probabilities for each sentence: the probability that each sentence was fully generated by a human, fully generated by AI, or a mixture of both^8^. For downstream analysis, we used the human-generated probability taken from this vector. These sentence-level probabilities were averaged by GPTZero into an essay-level prediction to which we will refer as P_human_. We expect that this measure will be correlated with the proportion of the essay that was human-written, although not an exact match. In essays written with heavy use of AI, many sentences will be flagged as AI-written or mixed; these sentences will have a low detected probability of being human-written, and the overall P_human_ will be low. Essays with mainly human-written sentences will conversely have a high sentence-wise detected probability of being written by a human and, therefore, a high P_human_.

### Statistical analysis

We performed all analysis in R 4.3.0 (R Foundation for Statistical Computing, Vienna, Austria) using the *tidyverse*^9^ frameworks. We generated tables using *gtsummary*^10^. We built general linear models using the predicted probability that each essay was human-generated as the dependent variable and demographic data or essay metadata as the independent predictors. We performed both univariate and multivariate modeling and derived the effect of each predictor from the beta coefficients (e^β^) from the trained models. We corrected for multiple hypothesis testing using the false discovery rate method. The code used to perform this analysis can be found on GitHub at https://github.com/nspies13/llm_use_in_medical_school_applications.

## Results

In the 2021–2022 cycle, the school received 6,137 applications before the deadline of November 15, 2021, while in the 2023–2024 cycle, the school received 5,055 applications before the deadline of November 15, 2023. For each essay type (“Personal Comments”, “Failure Essay”, “Anything Else”), we randomly selected 1,000 essays from each year as inputs to the GPTZero AI detector (**Table 1**), for a total of 6,000 essays.

**Table 1.** Demographics of the applicant cohorts.

| **Characteristic** | **2022, of N = 3,000<sup>1</sup>** | **2024, of N = 3,000<sup>1</sup>** | **p-value<sup>2</sup>** |
| --- | --- | --- | --- |
| **Age** | 22.6 (21.6, 23.7) | 22.7 (21.7, 23.8) | **0.021** |
| **Self-reported gender** |  |  | **<0.001** |
| *Female* | 1,599 (53%) | 1,494 (50%) |  |
| *Male* | 1,395 (47%) | 1,474 (49%) |  |
| *Other* | 6 (0.2%) | 32 (1.1%) |  |
| **Program** |  |  | 0.069 |
| *Regular MD* | 2,675 (89%) | 2,630 (88%) |  |
| *Combined MD/PhD* | 325 (11%) | 370 (12%) |  |
| **Visa Status** |  |  | 0.7 |
| *U.S. citizen* | 2778 (92.6%) | 2771 (92.4%) |  |
| *Other visa status* | 222 (7.4%) | 229 (7.6%) |  |
| **AMCAS socioeconomic status** |  |  | **0.02** |
| *EO1/EO2* | 382 (13%) | 444 (15%) |  |
| *Other* | 2618 (87%) | 2556 (85%) |  |
| **GPA** | 3.90 (3.74, 3.97) | 3.91 (3.78, 3.98) | **<0.001** |
| **MCAT** | 518 (514, 522) | 519 (515, 521) | >0.9 |
| **Completed application** | 2897 (97%) | 2911 (97%) | 0.3 |
| **Invited to interview** | 809 (27%) | 805 (27%) | 0.9 |
| **Accepted** | 276 (9.2%) | 288 (9.6%) | 0.1 |

<sup>1</sup>Median (interquartile range) or n (%)
<sup>2</sup>Wilcoxon rank sum test; Pearson's chi-squared test

We hypothesized that AI use would be rare or absent in the baseline year (2021–2022), leading the detector to produce P_human_ probabilities near 1.00. Conversely, in the first application cycle after the widespread availability of generative AI tools (2023–2024), we expected a proportion of students to have written some or all of their essays with these tools, leading to lower P_human_ estimates.

### Human-generated negative controls and AI-generated positive controls are appropriately labeled by GPTZero

A series of negative controls known to be completely human-generated demonstrated a median P_human_ of 0.93 (95% CI = 0.86–1.00). Conversely, a series of completely AI-generated positive controls, for which no downstream editing was performed, demonstrated a median P_human_ of 0.01 (95% CI = 0.00–0.05). These results met our continuation criteria and we proceeded to our planned study.

### Higher proportion of predicted AI-generated text in 2024 Personal Comments essays than in pre-ChatGPT baseline year

We compared the P_human_ for 1,000 randomly sampled Personal Comments essays in the 2023– 2024 application cycle to the pre-ChatGPT 2021–2022 cycle (**Figure 1**). The median P_human_ was 0.77 in the ‘23-24 cycle (95% confidence interval 0.76–0.78), and 0.83 in the ‘21-22 baseline (95% CI 0.82–0.85). 2.7% of essays had P_human_ < 0.5 in the baseline cycle. In contrast, 12.3% of essays had P_human_ < 0.5 in the ‘23-24 cycle, significantly more than in the baseline (p < 0.001, Fisher’s exact test).

**Figure 1:**
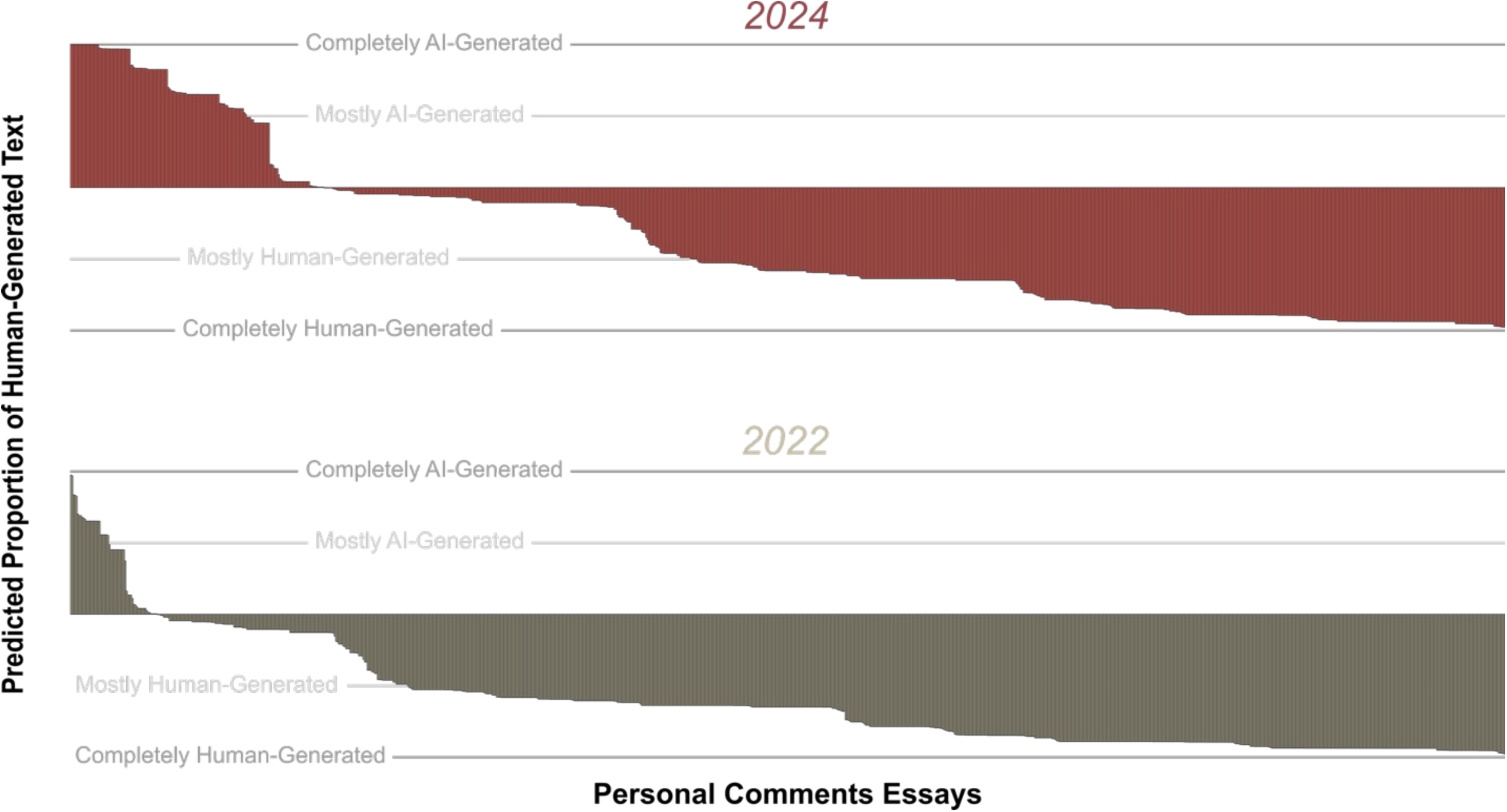
Waterfall plot of GPTZero’s estimated probabilities that 1,000 randomly sampled Personal Comments essays were AI- or human-generated. The 2024 application cycle (red, top) is compared to the pre-ChatGPT baseline of the 2022 cycle (tan, bottom).

### Analysis of factors associated with estimated human-generated probabilities from GPTZero

We analyzed 1,000 randomly selected Personal Comments, Failure, and Anything Else essays from the 2024 cycle using GPTZero’s AI detection algorithm. We then used average predicted probability that each sentence was entirely human-generated (P_human_) as the target label for univariate and multivariate linear models that included essay type, age, self-reported gender, program type, and visa status (used as a surrogate for native English proficiency).

In univariate analysis (**Table 2**), the essays in the secondary application had lower P_human_ than the universal Personal Comments essay by an average of 5–9% (p < 0.001). For every one-year increase in applicant age, there was an average of a 1% increase in P_human_ (p < 0.001). The essays from applicants requiring a visa had P_human_ of 8% less than U.S. citizens (p < 0.001). For each tenth of a point increase in grade point average, the P_human_ was 1% lower (p < 0.001). Incomplete applications had a 9% higher P_human_ than complete ones (p = 0.014). No significant differences were observed based on program type, self-reported gender, socioeconomic status, or MCAT score.

**Table 2:** Univariate analysis of application-related factors as predictors of P_human_, interpreted as the estimated percentage of the essay that was written by a human. Effect size is calculated as exp(beta). For age, effect size gives the increase in odds associated with a unit increase of one year; for MCAT, with a one-point increase in three-digit score; for GPA, with a 0.1-quality point increase in undergraduate GPA.

| Characteristic | N | Effect<sup>1</sup> | 95% CI<sup>2</sup> | p-value | q-value<sup>3</sup> |
| --- | --- | --- | --- | --- | --- |
| **Essay** |  |  |  |  |  |
| *Personal Comments* | 1,000 | — | — |  |  |
| *Failure Essay* | 1,000 | 0.91 | 0.88, 0.93 | <0.001 | **<0.001** |
| *Anything Else Essay* | 1,000 | 0.95 | 0.92, 0.97 | <0.001 | **<0.001** |
| **Age** | 3,000 | 1.01 | 1.01, 1.01 | <0.001 | **<0.001** |
| **Self-Reported Gender** |  |  |  |  |  |
| *Female* | 1,494 | — | — |  |  |
| *Male* | 1,474 | 1.00 | 0.98, 1.03 | 0.7 | 0.7 |
| *Other* | 32 | 1.11 | 0.99, 1.24 | 0.081 | 0.10 |
| **Program** |  |  |  |  |  |
| *Regular MD* | 2,630 | — | — |  |  |
| *Combined MD/PhD* | 370 | 1.02 | 0.98, 1.05 | 0.4 | 0.5 |
| **Visa** |  |  |  |  |  |
| *U.S. citizen* | 2,771 | — | — |  |  |
| *Other visa status* | 229 | 0.92 | 0.88, 0.96 | <0.001 | **<0.001** |
| **MCAT** | 2,998 | 1.00 | 1.00, 1.00 | 0.040 | 0.060 |
| **GPA** | 3,000 | 0.99 | 0.98, 1.0 | <0.001 | **<0.001** |
| **Application completion** |  |  |  |  |  |
| *Completed* | 2,911 | — | — |  |  |
| *Not completed* | 89 | 1.08 | 1.02, 1.14 | 0.014 | **0.026** |

<sup>1</sup>Effect = $e^{\beta}$ in the regression model, representing the average increase in estimated $P_{\text{human}}$ associated with a unit increment in each feature.
<sup>2</sup>CI = Confidence interval
<sup>3</sup>False discovery rate correction for multiple testing

**Table 3:** Multivariate analysis of factors correlated with admissions outcomes in 815 applicants with complete data from the 2023–2024 admissions cycle. “Probability Human” refers to the determination made by GPTZero for the Personal Comments essay. For age, the odds ratio gives the incremental increase in odds of each outcome associated with a unit increase of one year; for MCAT, with a one-point increase in three-digit score; for GPA, with a 0.1-quality point increase in undergraduate GPA.

| Characteristic | *Interviewed*<br>(142 of 815, 17%) |  |  |  |  | *Accepted*<br>(79 of 815, 10%) |  |  |  |
| --- | --- | --- | --- | --- | --- | --- | --- | --- | --- |
|  | OR<sup>1</sup> | 95% CI<sup>1</sup> | p-value | q-value<sup>2</sup> |  | OR<sup>1</sup> | 95% CI<sup>1</sup> | p-value | q-value<sup>2</sup> |
| **Probability Human** | 1.61 | 0.87, 3.02 | 0.13 | 0.2 |  | 1.14 | 0.46, 2.96 | 0.8 | 0.8 |
| **Age** | 1.09 | 1.00, 1.18 | 0.038 | 0.061 |  | 1.17 | 1.03, 1.30 | 0.010 | **0.019** |
| **Self-Reported Gender** |  |  |  |  |  |  |  |  |  |
| Female | — | — |  |  |  | — | — |  |  |
| Male | 0.51 | 0.37, 0.72 | <0.001 | **<0.001** |  | 0.59 | 0.35, 0.97 | 0.040 | 0.064 |
| **Program** |  |  |  |  |  |  |  |  |  |
| Regular MD | — | — |  |  |  | — | — |  |  |
| Combined MD/PhD | 0.76 | 0.45, 1.25 | 0.3 | 0.3 |  | 1.79 | 0.95, 3.27 | 0.063 | 0.084 |
| **Visa** |  |  |  |  |  |  |  |  |  |
| U.S. citizen | — | — |  |  |  | — | — |  |  |
| Other visa status | 1.95 | 0.99, 3.75 | 0.049 | 0.066 |  | 1.40 | 0.50, 3.41 | 0.5 | 0.6 |
| **SES** |  |  |  |  |  |  |  |  |  |
| Other | — | — |  |  |  | — | — |  |  |
| EO1/EO2 | 3.31 | 2.08, 5.29 | <0.001 | **<0.001** |  | 3.36 | 1.80, 6.15 | <0.001 | **<0.001** |
| **MCAT** | 1.19 | 1.14, 1.24 | <0.001 | **<0.001** |  | 1.18 | 1.11, 1.26 | <0.001 | **<0.001** |
| **GPA** | 1.81 | 1.50, 2.22 | <0.001 | **<0.001** |  | 1.61 | 1.24, 2.19 | <0.001 | **0.002** |

<sup>1</sup>OR = Odds ratio
<sup>2</sup>False discovery rate correction for multiple testing

In multivariate analysis including the same variables (**Figure 2**), the associations between P_human_ and essay type, age, and visa status remained statistically significant, while the association with GPA and completeness of application moved outside the significance threshold of 0.05. There was still no significant relationship between P_human_ and self-reported gender, program type, socioeconomic status, or MCAT score.

**Figure 2:**
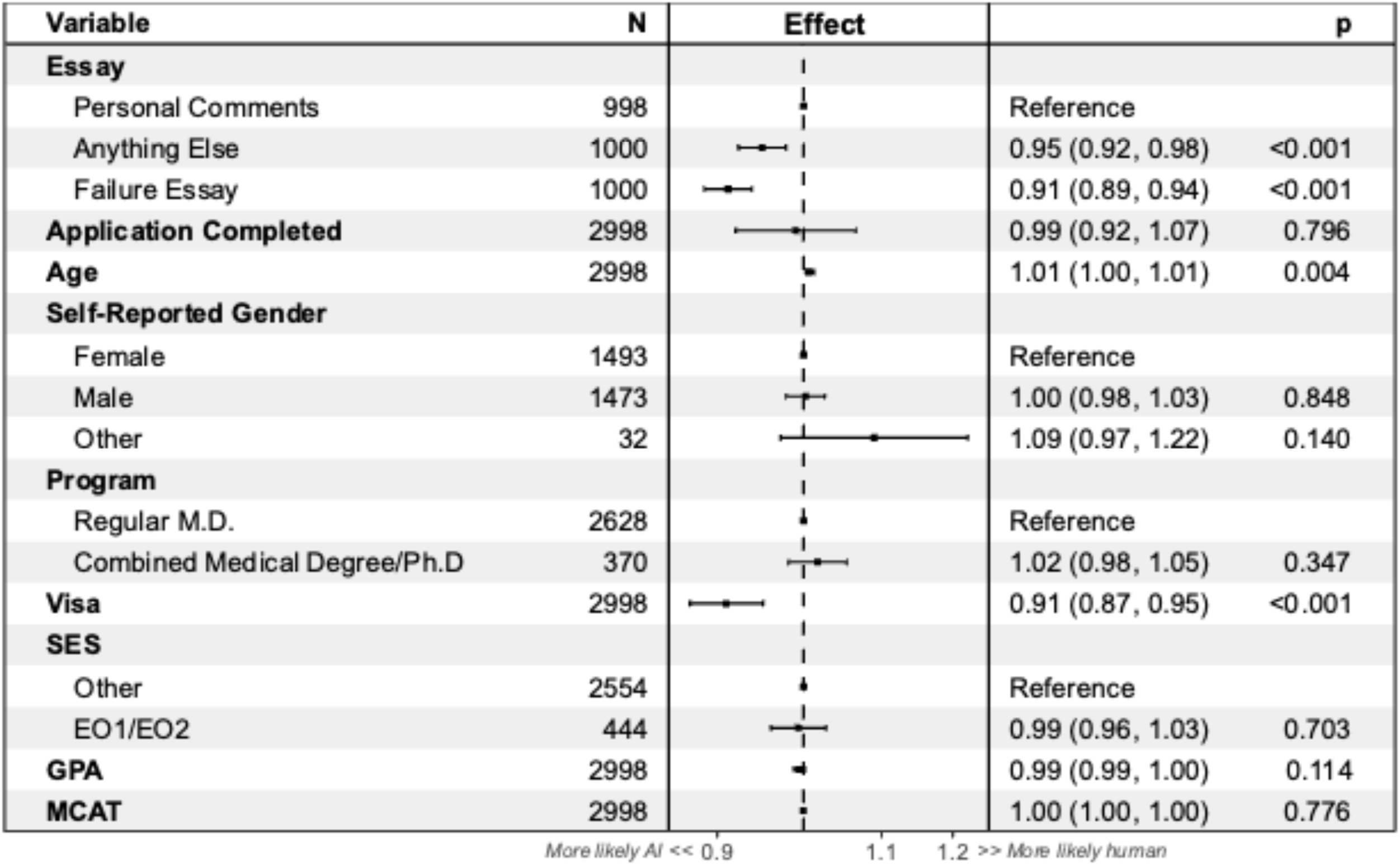
Multivariate analysis of the association between application parameters and GPTZero-predicted probabilities that each essay was human-generated in the 2024 application cycle. The magnitude of the effect is calculated as exp(beta). An effect greater than 1 indicates that the factor was associated with an increased P_human_. For age, effect gives the increase in odds associated with a unit increase of one year; for MCAT, with a one-point increase in three-digit score; for GPA, with a 0.1-quality point increase in undergraduate GPA.

We hypothesized that students who applied late might have been more likely to use AI in order to meet the deadline. Moreover, we hypothesized that applicants declaring only majors in science, technology, engineering or mathematics might be less comfortable with written expression and more likely to use AI. However, in exploratory analyses (data not shown), we found no correlation between P_human_ and date of application submission (r = 0.01) or whether the applicant declared a STEM major (p = 0.5).

### AI use is not associated with admissions outcomes

At the study school, application essays are used holistically at every stage of the review process and have the potential to affect admissions decisions. We tested whether AI use was associated with a favorable admissions outcome—invitation for an interview or, ultimately, acceptance to the medical school—in the 2023–2024 application cycle. For interview invitations, male applicants had an odds ratio of 0.51 (less likely to be interviewed), and applications with an EO1/EO2 for socioeconomic status had an odds ratio of 3.3 (more likely to be interviewed). A 1-point increase in MCAT score was associated with an odds ratio of 1.19 for receiving an interview, while each 0.1 quality point increase in GPA was associated with an odds ratio of 1.81. For acceptance, self-reported gender, program type, and visa status were not associated with acceptance (**Figure 3**). Low socioeconomic status, a one-year increase in age, one-point increase in MCAT score, and one-tenth of a point increase in GPA were all statistically significantly associated with acceptance (p < 0.001), with odds ratios of 3.36, 1.17, 1.18, and 1.61, respectively. When P_human_ was added to the rest of the application metadata, AI use was not significantly associated with either of these admissions outcomes.

## Discussion

We report our analysis of a natural experiment, comparing a cohort of medical school applicants that applied in 2021–2022, before LLM-driven chatbots were widely available, to the cohort that applied in 2023–2024, after the release of ChatGPT and GPT-3/4, when chatbots had become easily accessible, free, and frequently discussed in mainstream media and academic circles.

In the baseline year, we found that 2.7% of essays were estimated to have a P_human_ of < 0.5, suggesting AI use in their preparation. While it is possible that some essays in this cohort were written using the earlier-generation generative AI tools, we believe it is more likely that this represents the population-wide lower limit of the detector due to false-positive determinations. Our major finding is that an incremental 12.3–2.7=9.6% of essays were predicted as being mostly or entirely AI-written in the 2023–2024 cycle, a statistically significant difference. We found that the estimated proportion of essays that was AI-generated was higher in secondary essays and in essays from applicants with lower GPA (but not MCAT). There was no evidence that AI use was associated with socioeconomic status or undergraduate major. We did not find that late applicants were more likely to use AI than others. AI use did not significantly correlate with admissions decisions. Specifically, use of AI neither helped nor hindered applicants in gaining an interview or being accepted.

Strengths of this paper include the large data set, and the use of multiple timepoints (bracketing the introduction of widely available generative AI), and use of control inputs. Applicants to the study school represented approximately 10% of the 49,570 individuals who applied to U.S. medical schools through AMCAS in 2023–2024. The 2021–2022 group had, at most, rare and sporadic access to AI and therefore provides a benchmark against which the 2023–2024 group can be measured. Additionally, the highly confident positive predictions on the positive controls and negative predictions on the negative controls support the validity of the methods.

Limitations of the paper include its single-site nature, and that the applicants to the study school have, on average, higher academic achievement than the AMCAS applicant pool as a whole and may otherwise be non-representative. For practical reasons we used only one AI detector, although several are available. GPTZero was chosen due to its wide adoption and availability of an application programming interface. There might also be other approaches, such as applicant surveys, to learn about AI use. In our analysis of AI use in relation to application outcome, we only know the admissions actions at the study school; some applicants rejected at the study school were undoubtedly accepted at other schools.

A methodologic limitation for this project is that the distinction between human-written and AI-written text is excessively dualistic. While some applicants may blithely copy AI output directly into their application materials, it seems more plausible that they will adapt the AI text to their own situation, thus moving some or all sentences away from being purely AI-generated. Such edited text could pass for human-written although AI would have played a part in producing it. For the present study, we assume that at least some AI-derived text remains detectable after human editing.

An important additional caveat is that AI detection is known to be imperfect^11,12^. The sensitivity and specificity of AI detectors can be tuned, and generally is set so that the specificity is high at the expense of sensitivity^13^. This tradeoff is chosen because the consequences of a false-positive error (false detection of AI use, potentially leading to wrongful accusations of academic malfeasance or policy noncompliance) are less palatable than the consequences of a false-negative error. The result, however, is that some AI-generated text will be classified as human. Moreover, AI detectors may be more effective in identifying earlier and less advanced iterations of AI chatbots^7^, whereas newer models generate more human-like output and in some cases have been specifically designed to evade detection. Prompt engineering can be used to direct chatbots to write more like a human and the resulting text is less readily detected^12,14^. Since some chatbots are marketed on a freemium model (free tier/paid tier), students with higher resources may have access to more sophisticated versions. Anecdotally, some types of inputs, such as lists, may be erroneously flagged as AI-generated. This failure mode could lead to inaccurate detection in medical school application materials, which sometimes include lists of a student’s activities or publications. Together, these factors could lead to unjust outcomes if AI detection were deployed with real-world consequences (e.g., disqualifying applicants for detected use of AI).

AI detectors may misclassify the authentic work of non-native English writers as AI-generated^12^ due to more restricted vocabulary and syntax. Indeed, we found that applicants requiring a visa to study in the United States had a lower probability of human-generated text, which may reflect higher AI use, but could also reflect this failure mode of AI detection. English-language learners may also be more likely to use AI to proofread and correct authentic human-written essays. This use could be especially relevant for secondary essays where time is perceived to be short, and the penalty is high for making an error. We do not know how “proofreading” edits by AI affect P_human_, as compared with outright composition of writing samples by AI for which we have benchmarked P_human_.

As a final caveat, the importance placed on application essays may vary from one school to another. The impact of AI use on admissions outcomes, and the appropriate response, would vary accordingly.

Given the difficulty of definitively identifying AI use in applicant essays and the lack of clarity as to the appropriate response, we recommend continuing to study such data only in aggregate and on an informational basis. If there were evidence to suggest widespread AI use in answering a specific item, for example, the appropriate institutional response might be to design a new item that is less amenable to AI, rather than to penalize applicants who appear to be providing AI-generated answers. Similarly, we would hesitate to adopt software that would automatically flag AI use at the level of an individual applicant, although such software is likely to be available in the future. Playing a cat-and-mouse game around AI can only induce cynicism and erode overall trust in the application process, with potential to exacerbate existing disparities across socioeconomic or demographic factors.

It is unlikely that AI use can or should be entirely eliminated from medical school applications. Indeed, the 2025 AMCAS application contains a revised certification statement in which the applicant must agree that although AI use is permitted, the final product must be “a true reflection of [their] own work and represents [their] experiences.”^15^ Applicants’ reliance on AI could be a symptom of the heavy cognitive burden associated with preparing a complete medical school application. Since secondary applications showed more evidence of AI use than the common AMCAS application, schools should consider whether there is a benefit of adding writing samples to their school-specific secondary. These one-off tasks are numerous and are completed on a shorter timeline than the AMCAS essay, which could put applicants under pressure to take shortcuts. In turn, pre-health advisors should inform applicants that using AI deprives them of the opportunity to tell their story and highlight the unique contribution they will make in medicine.

## Data Availability

Due to the nature of the study, the data are not directly available but may be made available on reasonable request.

https://github.com/nspies13/llm_use_in_medical_school_applications

## Acknowledgments

The authors thank Christina Twist, MEd, MAcc, for serving as the honest broker for data retrieval.

## Ethical Approval

The Human Research Protection Office at Washington University School of Medicine determined that this study does not represent human subjects research (202401024). Moreover, permission was obtained from the Association of American Medical Colleges to use AMCAS data for this study.

## Disclaimers

None

## Previous Presentations

The manuscript was previously posted on MedRxiv.

## References

1. Singhal K, Azizi S, Tu T, et al. Large language models encode clinical knowledge. Nature. 2023;620(7972):172–180. doi:10.1038/s41586-023-06291-2

2. Qiu J, Xiong D. Generating Highly Relevant Questions. Published online 2019. doi:10.48550/ARXIV.1910.03401

3. Li J, Wang S, Zhang M, et al. Agent Hospital: A Simulacrum of Hospital with Evolvable Medical Agents. Published online 2024. doi:10.48550/ARXIV.2405.02957

4. Cotton DRE, Cotton PA, Shipway JR. Chatting and cheating: Ensuring academic integrity in the era of ChatGPT. Innovations in Education and Teaching International. 2024;61(2):228–239. doi:10.1080/14703297.2023.2190148

5. 2024 AMCAS Application Workbook. Published online 2023. Accessed April 3, 2024. https://students-residents.aamc.org/media/14376/download

6. GPTZero. Our Detection Technology. Accessed June 17, 2024. https://gptzero.me/technology

7. Elkhatat, A.M., Elsaid, I., Almeer, S. Evaluating the efficacy of AI content detection tools in differentiating between human and AI-generated text. International Journal for Educational Integrity. 2023;19:17.

8. “Document Predictions”, in GPTZero API Documentation. Published online 2024. Accessed April 3, 2024. https://gptzero.stoplight.io/docs/gptzero-api/707838f7e089d-document-predictions

9. Wickham H, Averick M, Bryan J, et al. Welcome to the Tidyverse. JOSS. 2019;4(43):1686. doi:10.21105/joss.01686

10. Sjoberg DD, Whiting K, Curry M, Lavery JA, Larmarange J. Reproducible Summary Tables with the gtsummary Package. The R Journal. 2021;13(1):570–580. doi:10.32614/RJ-2021-053

11. Rashidi HH, Fennell BD, Albahra S, Hu B, Gorbett T. The ChatGPT conundrum: Human-generated scientific manuscripts misidentified as AI creations by AI text detection tool. J Pathol Inform. 2023;14:100342. doi:10.1016/j.jpi.2023.100342

12. Liang W, Yuksekgonul M, Mao Y, Wu E, Zou J. GPT detectors are biased against non-native English writers. Patterns. 2023;4(7):100779. doi:10.1016/j.patter.2023.100779

13. Habibzadeh F. GPTZero Performance in Identifying Artificial Intelligence-Generated Medical Texts: A Preliminary Study. J Korean Med Sci. 2023;38(38):e319. doi:10.3346/jkms.2023.38.e319

14. Perkins M, Roe J, Vu BH, et al. GenAI Detection Tools, Adversarial Techniques and Implications for Inclusivity in Higher Education. Published online March 28, 2024. doi:10.48550/arXiv.2403.19148

15. 2025 AMCAS Applicant Guide. Published online 2024. Accessed July 6, 2024. https://students-residents.aamc.org/applying-medical-school-amcas/publication/2025-amcas-applicant-guide

