## Supplemental Table 2 for "Estimating the Prevalence of Generative AI Use in Medical School Application Essays"

**Supplemental Table 2**: Positive control essays generated entirely using AI chatbots.

| Essay Type | Prompt | Model | Text |
| --- | --- | --- | --- |
| Personal Comments | Please write a Personal Comments essay for my medical school application. | ChatGPT 3.5 | As a child, I found solace in the gentle touch of my mother’s hands as she tended to my scraped knees and bruised ego. Her nurturing presence instilled in me a deep-seated belief in the power of compassion and care. Little did I know that these childhood experiences would lay the foundation for my journey towards becoming a physician.  Growing up, I was always drawn to science and fascinated by the intricate workings of the human body. However, it was not until high school when I volunteered at a local hospital that I truly discovered my passion for medicine. Witnessing the dedication of healthcare professionals as they provided comfort and support to patients during their most vulnerable moments left an indelible mark on me. It was then that I realized my calling—to dedicate my life to the service of others through medicine.  Throughout my undergraduate years, I pursued opportunities to expand my understanding of healthcare and to hone my skills as a future physician. Whether conducting research in the laboratory or volunteering at a free clinic, each experience reinforced my commitment to medicine and deepened my understanding of the diverse needs of patients.  One particular experience that profoundly impacted me was my involvement in a medical mission trip to a rural community in [location]. There, I witnessed firsthand the disparities in healthcare access and the profound impact it had on the lives of individuals and families. Despite the challenges, I was inspired by the resilience and strength of the community members, and it reaffirmed my belief in the transformative power of compassionate healthcare.  As I reflect on my journey thus far, I am grateful for the support and guidance of mentors who have encouraged me to pursue my dreams relentlessly. Their wisdom and encouragement have shaped me into the compassionate and empathetic individual I am today.  Looking ahead, I am eager to embark on the next chapter of my journey—the pursuit of medical education. I am drawn to the holistic approach of [Medical School Name] and its commitment to fostering compassionate and culturally competent physicians. I am confident that the rigorous curriculum and diverse clinical experiences offered at [Medical School Name] will equip me with the knowledge and skills necessary to make a meaningful difference in the lives of patients and communities.  In conclusion, my journey to medicine has been guided by a deep-rooted desire to serve others and a belief in the transformative power of compassionate care. I am excited about the opportunity to continue my education at [Medical School Name] and to embark on a lifelong journey of learning, growth, and healing. |
| Personal Comments | Please write a Personal Comments essay for my medical school application. There is a limit of 5,300 characters. Don't mention the name of any specific school. | ChatGPT 3.5 | In every moment of my journey towards medicine, I have been driven by a profound sense of empathy and a relentless pursuit of purpose. From the earliest seeds planted by my grandmother's struggle with Alzheimer's disease to the fertile soil of my undergraduate studies in biology, each experience has shaped my path and fortified my commitment to serving others as a physician.  As a child, I was captivated by the intricate workings of the human body and fascinated by the potential of medicine to heal and transform lives. However, it was not until I witnessed firsthand the challenges faced by my grandmother as she navigated the complexities of Alzheimer's disease that my passion for medicine was ignited. Serving as her caregiver alongside my family, I learned the true meaning of compassion and resilience—values that would guide me on my journey towards becoming a physician.  Throughout my undergraduate years, I immersed myself in the study of biology, seeking to deepen my understanding of the mechanisms underlying human health and disease. It was during this time that I discovered my love for research and the pursuit of knowledge. Engaging in laboratory work and independent research projects allowed me to explore the intricacies of biomedical science and hone my critical thinking skills—a foundation that I believe will serve me well as a future physician.  Beyond the confines of the laboratory, I sought out opportunities to engage with communities and address healthcare disparities. Whether volunteering at local clinics or participating in medical mission trips abroad, I witnessed the transformative power of accessible healthcare and the profound impact it has on the lives of individuals and communities. These experiences reinforced my commitment to serving marginalized populations and fueled my desire to advocate for equitable access to healthcare for all.  As I reflect on my journey thus far, I am grateful for the support and guidance of mentors who have encouraged me to pursue my dreams relentlessly. Their wisdom and encouragement have shaped me into the compassionate and empathetic individual I am today.  Looking ahead, I am eager to embark on the next chapter of my journey—the pursuit of medical education. I am drawn to institutions that share my values and commitment to social responsibility. I am excited about the opportunity to contribute to the vibrant community of learners and scholars, and to continue my journey of growth and discovery as a future physician.  In conclusion, my journey to medicine has been guided by a deep-rooted desire to serve others and a belief in the transformative power of compassionate care. I am excited about the opportunity to continue my education and training as a physician and to make a meaningful difference in the lives of patients and communities. Thank you for considering my application, and I look forward to the opportunity to contribute to the field of medicine in a meaningful and impactful way. |
| Personal Comments | Please write a Personal Comments essay for my medical school application. Here is the prompt: "Consider and write your Personal Comments Essay carefully; many admissions committees place significant weight on the essay. Here are some questions you may want to consider while writing the essay: Why have you selected the field of medicine? What motivates you to learn more about medicine? What do you want medical schools to know about you that has not been disclosed in other sections of the application? In addition, you may wish to include information such as: Unique hardships, challenges, and obstacles that may have influenced your educational pursuits. Comments on significant fluctuations in your academic record not explained elsewhere in your application." | ChatGPT 3.5 | Medicine, to me, is not just a profession but a calling—a profound commitment to alleviate suffering and foster healing in the lives of others. My journey towards pursuing a career in medicine has been shaped by a myriad of experiences, each contributing to my unwavering dedication to this noble profession.  From a young age, I was exposed to the fragility of human health through the lens of my family's medical history. Witnessing the resilience of my loved ones in the face of illness ignited within me a profound curiosity about the intricacies of the human body and the role of healthcare in restoring wellness. As I grew older, this curiosity blossomed into a deep-seated passion for medicine, fueled by a desire to make a tangible difference in the lives of individuals and communities.  Throughout my academic journey, I have encountered numerous challenges and obstacles that have tested my resolve and strengthened my determination to pursue medicine. As a first-generation college student from a low-income background, I navigated the complexities of the educational system with limited resources and support. Despite facing financial hardships and academic uncertainties, I refused to let adversity define my path. Instead, I embraced each obstacle as an opportunity for growth and resilience, channeling my experiences into a driving force for success.  One significant fluctuation in my academic record occurred during my sophomore year, when unforeseen family responsibilities necessitated a temporary hiatus from my studies. During this time, I assumed caretaking duties for ailing family members, balancing their needs alongside my academic pursuits. Though challenging, this experience instilled in me a deep sense of empathy and compassion—an understanding of the human experience beyond the confines of textbooks and classrooms.  What sets me apart as a candidate for medical school is not just my academic achievements, but rather, my lived experiences and unwavering commitment to service. From volunteering at a local clinic to participating in medical mission trips abroad, I have sought opportunities to engage with diverse communities and address healthcare disparities firsthand. These experiences have reinforced my belief in the transformative power of compassionate care and fueled my desire to serve marginalized populations as a future physician.  As I embark on this next chapter of my journey, I am drawn to [Medical School Name] not only for its esteemed reputation but also for its commitment to fostering compassionate and socially conscious healthcare professionals. I am eager to contribute to the vibrant community at [Medical School Name], leveraging my unique background and experiences to make a meaningful impact in the lives of patients and communities alike.  In conclusion, my journey to medicine has been characterized by resilience, compassion, and an unwavering commitment to service. I am confident that my diverse experiences and steadfast dedication to healthcare will enable me to thrive as a student and future physician. Thank you for considering my application, and I look forward to the opportunity to contribute to the transformative work of [Medical School Name]. |
| Personal Comments | Please write a Personal Comments essay for my medical school application. I would like to emphasize my dedication to reducing health inequities. There is a limit of 5,300 characters. Don't mention the name of any specific school. | ChatGPT 3.5 | In every community, there exists a stark contrast between those who have access to quality healthcare and those who do not. This glaring disparity has fueled my passion for medicine and ignited within me a fervent dedication to reducing health inequities.  Growing up in a socioeconomically diverse neighborhood, I witnessed firsthand the profound impact of unequal access to healthcare. Whether volunteering at local clinics or participating in medical outreach programs, I encountered individuals and families who faced barriers to essential healthcare services due to factors such as financial constraints, language barriers, and systemic injustices.  These experiences instilled within me a deep sense of empathy and a commitment to advocating for those who are marginalized and underserved. From organizing health education workshops to advocating for policy changes that address healthcare disparities, I have sought out opportunities to raise awareness and effect tangible change within my community and beyond.  One particularly impactful experience that solidified my dedication to health equity occurred during a medical mission trip to a rural community. Witnessing the challenges faced by individuals who lacked access to basic healthcare services opened my eyes to the pervasive nature of health disparities and underscored the urgent need for systemic change.  As an undergraduate student, I pursued coursework and research opportunities that allowed me to delve deeper into the root causes of health inequities. From studying the social determinants of health to examining the impact of healthcare policies on vulnerable populations, I sought to gain a comprehensive understanding of the complex factors that contribute to disparities in healthcare access and outcomes.  Beyond academia, I actively engaged in advocacy efforts aimed at addressing health inequities on a broader scale. Whether participating in rallies for healthcare reform or collaborating with community organizations to develop culturally competent healthcare initiatives, I have been steadfast in my commitment to advancing health equity as a fundamental human right.  Looking ahead, I am eager to continue my journey as a future physician and advocate for health equity. I am drawn to institutions that share my values and commitment to social responsibility, and I am excited about the opportunity to contribute to the collective effort towards a more just and equitable healthcare system.  In conclusion, my dedication to reducing health inequities is rooted in a deep-seated belief in the inherent dignity and worth of every individual. I am committed to leveraging my education, experiences, and passion for advocacy to create a world where access to healthcare is not determined by one's socioeconomic status or background. Thank you for considering my application, and I look forward to the opportunity to contribute to the field of medicine in a meaningful and impactful way. |
| Personal Comments | Please write a Personal Comments essay for my medical school application. I would like to emphasize my dedication to biomedical research, based on my experiences doing microbiology research as an undergraduate. | ChatGPT 3.5 | From the moment I stepped into the laboratory, I was captivated by the intricate dance of molecules and the promise of scientific discovery. My undergraduate journey in microbiology research has ignited within me a fervent dedication to advancing biomedical knowledge and unlocking the mysteries of the human body.  As an undergraduate researcher, I had the privilege of delving into the fascinating world of microbiology under the mentorship of esteemed faculty members. Through hands-on laboratory experiences and independent research projects, I gained invaluable insights into the complexities of microbial physiology and the mechanisms underlying infectious diseases.  One particularly impactful experience during my undergraduate years was my involvement in a research project focused on investigating the role of bacterial biofilms in antibiotic resistance. This project not only deepened my understanding of microbial pathogenesis but also underscored the urgent need for innovative approaches to combatting antibiotic-resistant infections—a pressing public health challenge with far-reaching implications.  Motivated by the potential of biomedical research to drive transformative change, I immersed myself in the scientific process, from formulating research questions to conducting experiments and analyzing data. The thrill of discovery and the pursuit of knowledge fueled my passion for research, motivating me to push the boundaries of scientific inquiry and contribute to the collective effort towards improving human health.  Beyond the laboratory bench, I actively sought out opportunities to disseminate my findings and engage with the broader scientific community. Whether presenting at research conferences or publishing in scientific journals, I embraced every opportunity to contribute to the advancement of biomedical knowledge and foster collaboration within the scientific community.  My dedication to biomedical research extends beyond the confines of academia. I am committed to leveraging my research experiences to address pressing healthcare challenges and improve patient outcomes. Whether developing novel therapeutics or enhancing diagnostic technologies, I am driven by a desire to translate scientific discoveries into tangible solutions that benefit society as a whole.  As I embark on the journey towards becoming a physician-scientist, I am excited about the opportunity to integrate my passion for research with clinical practice. I am drawn to institutions that prioritize interdisciplinary collaboration and innovation in biomedical research, and I am eager to contribute to the vibrant research community as a future physician.  In conclusion, my journey in microbiology research has instilled within me a deep-seated passion for scientific discovery and a commitment to advancing biomedical knowledge for the betterment of humanity. I am grateful for the opportunity to continue this journey as a future physician-scientist and to contribute to the intersection of medicine and research in meaningful and impactful ways. Thank you for considering my application, and I look forward to the opportunity to further discuss how I can contribute to the field of medicine. |
| Personal Comments | Please write a Personal Comments essay for my medical school application. I would like to emphasize my experience caring for my grandmother, who had Alzheimer's disease. There is a limit of 5,300 characters. Don't mention the name of any specific school. | ChatGPT 3.5 | In the quiet moments spent by my grandmother's side, I learned the true meaning of compassion and resilience. Her journey with Alzheimer's disease not only shaped my perspective on healthcare but also inspired my commitment to serving others as a future physician.  As a young child, I was captivated by my grandmother's gentle demeanor and unwavering love. She was the matriarch of our family—a beacon of strength and wisdom. However, as the years passed, I began to notice subtle changes in her behavior and cognition. Simple tasks became increasingly challenging, and memories once cherished faded into the recesses of her mind.  The diagnosis of Alzheimer's disease cast a shadow of uncertainty over our family, but it also ignited within us a profound sense of determination to provide the best possible care for my grandmother. From coordinating medical appointments to assisting with daily activities, I assumed the role of caregiver alongside my parents, learning to navigate the complexities of Alzheimer's with patience and empathy.  Despite the challenges posed by her illness, my grandmother's spirit remained unbroken. In moments of clarity, she would share stories of her youth, offering glimpses into a life rich with love and adventure. These precious moments served as a poignant reminder of the person she once was—a testament to the resilience of the human spirit in the face of adversity.  Caring for my grandmother was not without its struggles. Witnessing the progressive decline of her cognitive abilities was both heartbreaking and humbling. Yet, amidst the hardships, I discovered the transformative power of compassion—the ability to provide comfort and support in the midst of uncertainty.  My experience caring for my grandmother sparked a deep-seated passion for healthcare and a desire to make a meaningful difference in the lives of individuals and families affected by Alzheimer's and other neurodegenerative diseases. It also underscored the importance of holistic and compassionate care in addressing the complex needs of patients with chronic conditions.  As I embark on the journey towards becoming a physician, I carry with me the lessons learned at my grandmother's bedside—the importance of empathy, resilience, and unwavering dedication to serving others. I am committed to honoring her legacy by advocating for compassionate and patient-centered care, particularly for vulnerable populations like those affected by Alzheimer's disease.  In considering medical schools, I am drawn to institutions like [Medical School Name] that prioritize a holistic approach to healthcare and value the humanistic aspects of medicine. I am eager to contribute to the vibrant community at [Medical School Name], leveraging my personal experiences to inform my practice as a future physician.  In conclusion, my journey in caring for my grandmother with Alzheimer's disease has profoundly shaped my aspirations and values as a future physician. I am grateful for the opportunity to honor her memory through my dedication to compassionate and empathetic patient care. Thank you for considering my application, and I look forward to the opportunity to further discuss how I can contribute to the mission of [Medical School Name]. |
| MD-PhD Motivation | Please write an essay for my application to MD-PhD programs, describing my reasons for pursuing the combined MD-PhD degree. This essay is limited to 3,000 characters | ChatGPT 3.5 | The decision to pursue the MD-PhD dual degree stems from my unwavering commitment to bridging the gap between clinical practice and scientific inquiry. As a passionate advocate for translational research, I am driven by the belief that groundbreaking discoveries at the bench can catalyze innovative treatments at the bedside.  The MD-PhD program uniquely aligns with my aspirations to make meaningful contributions to both patient care and biomedical knowledge. By seamlessly integrating medical training with rigorous research experiences, this dual-degree pathway empowers me to address complex healthcare challenges through a multifaceted approach.  My journey in healthcare began with a fascination for the intricacies of the human body and a desire to alleviate suffering. However, it was through my undergraduate research experiences that I discovered the transformative potential of biomedical research in improving patient outcomes. From investigating novel therapeutics to exploring disease mechanisms, I have witnessed firsthand the profound impact of research in shaping the future of medicine.  As a future physician-scientist, I am committed to leveraging my expertise in both clinical practice and research to advance healthcare on multiple fronts. Whether developing personalized treatment strategies or unraveling the molecular underpinnings of disease, I am driven by a shared vision of translating scientific discoveries into tangible benefits for patients.  The MD-PhD program represents the ideal platform to cultivate my skills as a clinician-scientist and nurture my passion for translational research. By fostering interdisciplinary collaboration and providing immersive training experiences, this program offers unparalleled opportunities to make meaningful contributions to scientific knowledge and patient care.  In conclusion, the MD-PhD dual degree is not merely a stepping stone in my academic journey but a reflection of my deep-seated commitment to advancing medicine through research and innovation. I am eager to embark on this transformative path, and I am confident that the combined training offered by the MD-PhD program will equip me with the skills and knowledge needed to make a lasting impact in the field of healthcare. |
| MD-PhD Motivation | Please write an essay for my application to MD-PhD programs, describing my reasons for pursuing the combined MD-PhD degree. I am interested in studying neurodegeneration. This essay is limited to 3,000 characters | ChatGPT 3.5 | My passion for understanding and combatting neurodegenerative diseases has fueled my decision to pursue the combined MD-PhD degree. The MD-PhD program uniquely positions me to integrate clinical training with cutting-edge research, empowering me to make meaningful contributions to the field of neuroscience.  Neurodegenerative diseases represent a pressing healthcare challenge with devastating consequences for individuals and families worldwide. The complexity of these disorders demands a multifaceted approach that combines clinical insight with scientific inquiry. As a future physician-scientist, I am committed to leveraging my dual expertise to unravel the mechanisms underlying neurodegeneration and develop novel therapeutic strategies.  My journey in neuroscience research has been characterized by a deep-seated curiosity and a desire to address unmet clinical needs. From exploring the molecular pathways implicated in Alzheimer's disease to investigating potential biomarkers for Parkinson's disease, I have been captivated by the intricacies of the nervous system and the opportunity to make a tangible impact on patient care.  The MD-PhD program represents the ideal pathway for me to pursue my passion for neurodegeneration. By providing immersive research experiences alongside comprehensive medical training, this program offers a unique platform to translate scientific discoveries into clinical applications. I am eager to collaborate with leading experts in the field, engage in interdisciplinary research, and contribute to the development of innovative therapies for neurodegenerative diseases.  In conclusion, the MD-PhD dual degree is not only a means to deepen my understanding of neurodegeneration but also a vehicle for effecting positive change in the lives of patients and families affected by these devastating disorders. I am excited about the opportunity to embark on this transformative journey, and I am confident that the combined training offered by the MD-PhD program will empower me to make significant contributions to the field of neuroscience. |
| MD-PhD Motivation | Please write an essay for my application to MD-PhD programs, describing my reasons for pursuing the combined MD-PhD degree. I am interested in studying cancer. This essay is limited to 3,000 characters | ChatGPT 3.5 | My decision to pursue the combined MD-PhD degree is rooted in my passion for unraveling the complexities of cancer and developing innovative treatments. The MD-PhD program offers a unique opportunity to seamlessly integrate clinical training with cutting-edge research, empowering me to make significant contributions to the field of oncology.  Cancer remains one of the most formidable challenges in modern medicine, with far-reaching implications for individuals and society as a whole. As a future physician-scientist, I am driven by a deep-seated desire to translate scientific discoveries into tangible advancements in cancer diagnosis, treatment, and prevention.  My journey in cancer research began with a fascination for the molecular mechanisms driving oncogenesis and tumor progression. From investigating signaling pathways implicated in cancer development to exploring novel therapeutic targets, I have been captivated by the potential of research to transform our understanding of this complex disease.  The MD-PhD program represents the ideal pathway for me to pursue my passion for cancer research. By providing a comprehensive training experience that integrates clinical rotations with laboratory-based investigations, this program offers a holistic approach to addressing the multifaceted challenges of cancer.  I am eager to collaborate with leading experts in the field, leverage cutting-edge technologies, and contribute to the development of personalized therapies tailored to the unique molecular profiles of individual tumors. Through translational research, I aim to bridge the gap between bench and bedside, bringing new hope to patients and families affected by cancer.  In conclusion, the MD-PhD dual degree is not only a means to advance my career in cancer research but also a platform for effecting positive change in the lives of patients and communities impacted by this devastating disease. I am excited about the opportunity to embark on this transformative journey, and I am confident that the combined training offered by the MD-PhD program will equip me with the skills and knowledge needed to make a meaningful impact in the field of oncology. |
| MD-PhD Motivation | Please write an essay for my application to MD-PhD programs, describing my reasons for pursuing the combined MD-PhD degree. I am interested in studying autoimmunity. This essay is limited to 3,000 characters | ChatGPT 3.5 | My decision to pursue the combined MD-PhD degree is driven by my passion for unraveling the mysteries of autoimmunity and developing targeted therapies. The MD-PhD program offers a unique opportunity to seamlessly integrate clinical training with cutting-edge research, empowering me to make significant contributions to understanding and treating autoimmune diseases.  Autoimmunity represents a complex and multifaceted challenge in modern medicine, with far-reaching implications for individuals and society as a whole. As a future physician-scientist, I am deeply committed to translating scientific discoveries into tangible advancements in autoimmune disease diagnosis, treatment, and management.  My journey in autoimmunity research began with a fascination for the intricate interplay between the immune system and self-tolerance. From investigating the underlying mechanisms of autoimmune diseases to exploring novel therapeutic interventions, I have been captivated by the potential of research to transform our understanding of autoimmunity and improve patient outcomes.  The MD-PhD program represents the ideal pathway for me to pursue my passion for autoimmunity research. By providing a comprehensive training experience that integrates clinical rotations with laboratory-based investigations, this program offers a holistic approach to addressing the complex challenges of autoimmune diseases.  I am eager to collaborate with leading experts in the field, leverage state-of-the-art technologies, and contribute to the development of personalized therapies tailored to the specific immunological profiles of individual patients. Through translational research, I aim to bridge the gap between basic science discoveries and clinical applications, bringing new hope to patients affected by autoimmune diseases.  In conclusion, the MD-PhD dual degree is not only a means to advance my career in autoimmunity research but also a platform for effecting positive change in the lives of patients and communities impacted by these debilitating conditions. I am excited about the opportunity to embark on this transformative journey, and I am confident that the combined training offered by the MD-PhD program will equip me with the skills and knowledge needed to make a meaningful impact in the field of autoimmunity. |
| Failure Essay | Please write an essay for my medical school application. The prompt is: "Describe a time or situation where you have been unsuccessful or failed." The essay should be around 250 words long. | ChatGPT 3.5 | Facing failure is an inevitable part of life's journey, and one particular instance stands out vividly in my memory. During my sophomore year of college, I undertook a challenging research project in neurobiology. Excited by the opportunity to contribute to scientific discovery, I dove into the project with enthusiasm and determination. However, as weeks turned into months, I encountered unforeseen obstacles and setbacks that ultimately led to an unsuccessful outcome.  Despite investing countless hours into the project, my experiments yielded inconclusive results, and I found myself grappling with feelings of frustration and self-doubt. I had envisioned making significant contributions to the field of neuroscience, but instead, I was met with disappointment and uncertainty.  In the aftermath of this experience, I faced a pivotal moment of introspection. Rather than allowing failure to define me, I chose to embrace it as an opportunity for growth and learning. I reflected on the mistakes I had made and identified areas for improvement, from refining my experimental techniques to seeking guidance from mentors.  Through perseverance and resilience, I emerged from this setback with newfound determination and resolve. I redirected my focus towards honing my research skills and sought out additional opportunities to expand my scientific knowledge. With each subsequent research endeavor, I approached challenges with a renewed sense of confidence and humility, recognizing that failure is not a reflection of my capabilities but rather an essential stepping stone on the path to success.  This experience has taught me invaluable lessons about resilience, adaptability, and the importance of embracing failure as a catalyst for personal and professional growth. It has instilled within me a deep-seated belief in my ability to overcome obstacles and persevere in the face of adversity, qualities that I am confident will serve me well as I pursue a career in medicine. |
| Failure Essay | Please write an essay for my medical school application. The prompt is: "Describe a time or situation where you have been unsuccessful or failed." Focus on a time when I got a low grade in organic chemistry. The essay should be around 250 words long. | ChatGPT 3.5 | One particular instance of failure that has left a lasting impact on me occurred during my freshman year of college when I received a low grade in organic chemistry. As someone who has always been passionate about science and aspired to pursue a career in medicine, this setback felt particularly disheartening.  Organic chemistry was a notoriously challenging course, and despite my diligent efforts, I struggled to grasp certain concepts and apply them effectively. As the semester progressed, I found myself falling behind and becoming increasingly overwhelmed by the coursework.  Receiving a low grade in organic chemistry was a humbling experience that forced me to confront my limitations and reevaluate my approach to learning. Rather than allowing myself to be consumed by self-doubt and disappointment, I chose to view this setback as an opportunity for growth and self-improvement.  I sought out additional resources and support, including tutoring sessions and study groups, to help me better understand the material and improve my performance in the course. I also made a concerted effort to adopt more effective study strategies, such as breaking down complex concepts into manageable chunks and practicing problem-solving techniques regularly.  Ultimately, while the experience of receiving a low grade in organic chemistry was undeniably challenging, it served as a valuable lesson in resilience and perseverance. It taught me the importance of resilience in the face of adversity and the power of determination to overcome obstacles. Moreover, it reinforced my commitment to pursuing my passion for medicine, inspiring me to redouble my efforts and strive for excellence in all aspects of my academic and professional pursuits. |
| Failure Essay | Please write an essay for my medical school application. The prompt is: "Describe a time or situation where you have been unsuccessful or failed." Focus on a time when I did not do an effective job planning an activities fair. The essay should be around 250 words long. | ChatGPT 3.5 | One instance where I encountered failure was during my role in organizing an activities fair at my university. As a member of the student council, I was tasked with coordinating the event to showcase various extracurricular opportunities to incoming students. Despite my best intentions, the fair did not meet the expectations of both participants and attendees.  In hindsight, I realize that my failure stemmed from inadequate planning and communication. I underestimated the logistical challenges involved in coordinating such a large-scale event and failed to establish clear roles and responsibilities among the organizing team. As a result, there were instances of miscommunication and confusion, leading to delays and inefficiencies on the day of the fair.  Additionally, I did not allocate enough time for promoting the event and generating interest among students. While I assumed that the fair's significance would be self-evident, I failed to recognize the importance of effective marketing and outreach efforts in ensuring its success.  Despite these shortcomings, the experience taught me valuable lessons in leadership and event management. It highlighted the importance of meticulous planning, effective communication, and proactive problem-solving in achieving organizational goals. Moreover, it underscored the significance of humility and self-reflection in acknowledging and learning from failure.  Moving forward, I am committed to applying these lessons to future endeavors, both in academia and beyond. I recognize that failure is an inherent part of growth and development and that resilience and perseverance are essential traits in overcoming setbacks. As I embark on my journey towards a career in medicine, I am confident that this experience has equipped me with the skills and mindset necessary to navigate the challenges that lie ahead. |
| Failure Essay | Please write an essay for my medical school application. The prompt is: "Describe a time or situation where you have been unsuccessful or failed." Focus on a time when I did not support my friend when her mother was sick. The essay should be around 250 words long. | ChatGPT 3.5 | One of the most significant instances of failure in my life occurred when I did not adequately support my friend during a difficult period when her mother was sick. As someone who values compassion and empathy, my failure to be there for my friend during her time of need weighs heavily on my conscience.  At the time, I was preoccupied with my own academic and personal challenges, and I allowed these distractions to overshadow the importance of being there for my friend. Instead of offering a listening ear or extending a comforting presence, I failed to reach out and provide the support and solace that she desperately needed.  Reflecting on this experience, I realize that my failure stemmed from a lack of empathy and understanding. I underestimated the profound impact that illness can have on individuals and families, and I failed to recognize the significance of my role as a friend in providing emotional support during times of hardship.  Moreover, I recognize that my actions, or lack thereof, may have caused additional pain and isolation for my friend during an already challenging time. I deeply regret not being there for her when she needed me the most and understand the importance of acknowledging and addressing my shortcomings in order to learn and grow from them.  Moving forward, I am committed to being more attentive and supportive of those around me, especially during times of difficulty and distress. I have learned that true friendship requires compassion, empathy, and a willingness to stand by each other through life's ups and downs. As I continue on my journey towards becoming a physician, I am determined to apply these lessons in my interactions with patients and their families, recognizing the profound impact that empathy and compassion can have on healing and well-being. |
| Anything Else | Please write an essay to include in my medical school application. The prompt is: "Is there anything else you would like to share with the Committee on Admissions? Some applicants use this space to describe unique experiences and obstacles such as significant challenges in access to education, unusual socioeconomic factors, and/or identification with a particular culture, religion, race, ethnicity, sexual orientation, or gender identity and/or possibly challenges related to COVID or other issues in preparation for medical school." The essay should be around 250 words long. I would like to focus on how motivated I am to go to medical school in New York. | ChatGPT 3.5 | In reflecting upon my journey towards medical school, I am compelled to share my deep motivation to pursue this path within the vibrant landscape of New York. Growing up in a city that pulses with diversity, innovation, and opportunity, I have been inspired by the unique blend of cultures, perspectives, and experiences that define the fabric of New York.  As a native New Yorker, I have witnessed firsthand the transformative power of healthcare in our community. From the bustling streets of Manhattan to the vibrant neighborhoods of Brooklyn, I have seen how access to quality healthcare can profoundly impact individuals and families, regardless of socioeconomic status or background.  Moreover, New York's rich tapestry of cultures and identities has shaped my understanding of the importance of cultural competence and sensitivity in healthcare delivery. I am committed to serving diverse populations and addressing healthcare disparities within the dynamic and multicultural landscape of the city.  Furthermore, the challenges presented by the COVID-19 pandemic have underscored the resilience and dedication of healthcare professionals in New York. I am inspired by their tireless efforts to care for patients and communities during these unprecedented times, and I am eager to join their ranks as a future physician.  In conclusion, my motivation to attend medical school in New York stems from a deep-rooted desire to serve and contribute to the health and well-being of the diverse communities that call this city home. I am grateful for the opportunity to pursue my passion for medicine within the dynamic and vibrant environment of New York, and I am excited about the prospect of making a meaningful impact in the lives of those around me. Thank you for considering my application. |
| Anything Else | Please write an essay to include in my medical school application. The prompt is: "Is there anything else you would like to share with the Committee on Admissions? Some applicants use this space to describe unique experiences and obstacles such as significant challenges in access to education, unusual socioeconomic factors, and/or identification with a particular culture, religion, race, ethnicity, sexual orientation, or gender identity and/or possibly challenges related to COVID or other issues in preparation for medical school." The essay should be around 250 words long. I would like to focus on how I grew up in poverty and received free lunch at school. | ChatGPT 3.5 | Growing up in poverty and relying on free lunch at school has profoundly shaped my journey towards medical school. From a young age, I witnessed firsthand the challenges and barriers that individuals from disadvantaged backgrounds face in accessing healthcare and educational opportunities. As the child of hardworking parents who struggled to make ends meet, I learned to navigate the complexities of poverty with resilience and determination.  Receiving free lunch at school was both a humbling experience and a stark reminder of the socioeconomic disparities that exist within our society. Despite the financial hardships my family endured, I remained steadfast in my commitment to academic excellence and personal growth. I recognized education as a powerful tool for breaking the cycle of poverty and was determined to seize every opportunity to expand my horizons and pursue my dreams.  My experiences growing up in poverty have instilled within me a deep empathy for those facing similar challenges and a commitment to serving underserved populations. I am passionate about addressing healthcare disparities and advocating for equitable access to quality healthcare for all individuals, regardless of socioeconomic status.  Furthermore, navigating the challenges presented by poverty has honed my resilience, resourcefulness, and compassion—qualities that I believe are essential for success in the field of medicine. I am grateful for the opportunity to share my story with the Committee on Admissions and am eager to contribute to the medical community as a compassionate and culturally competent physician. Thank you for considering my application. |
| Anything Else | Please write an essay to include in my medical school application. The prompt is: "Is there anything else you would like to share with the Committee on Admissions? Some applicants use this space to describe unique experiences and obstacles such as significant challenges in access to education, unusual socioeconomic factors, and/or identification with a particular culture, religion, race, ethnicity, sexual orientation, or gender identity and/or possibly challenges related to COVID or other issues in preparation for medical school." The essay should be around 250 words long. I would like to focus on being a member of an under-represented minority group (I am Latino). | ChatGPT 3.5 | As a member of an underrepresented minority group, specifically Latino, I am compelled to share the unique experiences and perspectives that have shaped my journey towards medical school. Growing up in a Latino household, I was immersed in a rich tapestry of culture, traditions, and values that have deeply influenced my identity and aspirations.  Throughout my life, I have encountered challenges and obstacles that are emblematic of the disparities faced by underrepresented minorities in accessing education and healthcare. From navigating language barriers to confronting systemic inequities, I have witnessed firsthand the impact of these challenges on individuals and communities.  Despite the obstacles I have faced, my identity as a Latino has served as a source of strength and resilience. It has instilled within me a deep sense of pride in my heritage and a commitment to advocating for diversity, equity, and inclusion within the medical field.  As I embark on this journey towards medical school, I am passionate about leveraging my unique perspective to address healthcare disparities and improve health outcomes for underserved populations, particularly within the Latino community. I am committed to serving as a compassionate and culturally competent physician who is dedicated to providing equitable and culturally sensitive care to all individuals, regardless of background or circumstance.  Furthermore, I am acutely aware of the challenges presented by the COVID-19 pandemic, which has disproportionately impacted minority communities. I am inspired by the resilience and solidarity demonstrated by communities in the face of adversity, and I am eager to contribute to efforts to address the health and social disparities exacerbated by the pandemic.  In conclusion, my identity as a member of an underrepresented minority group has profoundly shaped my perspective and aspirations as I pursue a career in medicine. I am grateful for the opportunity to share my story with the Committee on Admissions and am committed to making a meaningful impact in the field of healthcare as a culturally competent and compassionate physician. Thank you for considering my application. |
| Experience | Please write a short text for my medical school application, describing my experience being inducted into a nursing honor society. There is a limit of 700 characters. | ChatGPT 3.5 | Induction into the nursing honor society was a defining moment in my academic and professional journey. It affirmed my dedication to excellence in patient care and inspired me to continue striving for the highest standards in nursing practice. Being recognized for my commitment to scholarship, leadership, and service has fueled my passion for making a meaningful difference in the lives of others through compassionate and evidence-based care. |
| Experience | Please write a short text for my medical school application, describing my experience as a teaching assistant for anatomy and physiology. There is a limit of 700 characters. | ChatGPT 3.5 | As a teaching assistant for anatomy and physiology, I had the privilege of fostering a deeper understanding of the human body among my peers. Guiding students through complex concepts and hands-on lab experiences, I witnessed the transformation of uncertainty into comprehension. This role enhanced my communication skills and solidified my passion for education and mentorship in healthcare. |
| Experience | Please write a short text for my medical school application, describing my experience as a research assistant in a lab studying muscle contractility. There is a limit of 700 characters. | ChatGPT 3.5 | As a research assistant in a lab studying muscle contractility, I contributed to groundbreaking discoveries in understanding muscle function. Conducting experiments and analyzing data, I delved into the intricacies of cellular mechanisms. This experience honed my analytical skills and deepened my appreciation for scientific inquiry, fueling my desire to pursue a career at the intersection of research and medicine. |
| Experience | Please write a short text for my medical school application, describing my experience being inducted into a nursing honor society. There is a limit of 700 characters plus an additional 1325 characters because this is one of my most meaningful experiences. | ChatGPT 3.5 | Induction into the nursing honor society was an incredibly meaningful experience that affirmed my dedication to the field of healthcare. Being recognized for my academic achievements, leadership qualities, and commitment to service has been a source of pride and validation. This honor has inspired me to continue striving for excellence in patient care and to uphold the values of scholarship, professionalism, and integrity in my practice. As a member of the nursing honor society, I have had the opportunity to collaborate with like-minded peers, engage in service projects, and participate in professional development activities. These experiences have not only enriched my understanding of nursing but have also reinforced my passion for making a positive impact in the lives of others. I am grateful for the support and encouragement of my mentors and colleagues throughout this journey, and I am excited to continue my growth as a nurse and future physician. |
| Experience | Please write a short text for my medical school application, describing my experience as a teaching assistant for anatomy and physiology. There is a limit of 700 characters plus an additional 1325 characters because this is one of my most meaningful experiences. | ChatGPT 3.5 | As a teaching assistant for anatomy and physiology, I played a vital role in guiding students through the complexities of the human body. This experience allowed me to deepen my understanding of the subject matter while fostering a supportive learning environment for my peers. I developed effective communication and leadership skills, which are essential in both medicine and education. Witnessing the moment when students grasped challenging concepts solidified my passion for teaching and mentorship. This role has been one of my most meaningful experiences, as it affirmed my commitment to lifelong learning and helping others succeed in their academic pursuits. |
| Experience | Please write a short text for my medical school application, describing my experience as a research assistant in a lab studying muscle contractility. There is a limit of 700 characters plus an additional 1325 characters because this is one of my most meaningful experiences. | ChatGPT 3.5 | As a research assistant in a lab studying muscle contractility, I delved into the intricate mechanisms governing muscle function. Through hands-on experiments and data analysis, I contributed to groundbreaking discoveries in the field. This experience honed my analytical skills and deepened my understanding of scientific inquiry. Witnessing the impact of our research on advancing medical knowledge has been immensely fulfilling and has solidified my passion for pursuing a career at the intersection of research and medicine. |
| Experience | Please write an essay of about 700 characters for my medical school application, explaining my experiences as a medical scribe. | ChatGPT 3.5 | As a medical scribe, I've had the privilege of working closely with healthcare providers in real-time patient encounters. This role has equipped me with invaluable clinical experience, honed my medical terminology proficiency, and improved my ability to accurately document patient histories, physical exams, and treatment plans. Collaborating with physicians has deepened my understanding of medical decision-making and enhanced my communication skills. My time as a medical scribe has reaffirmed my passion for medicine and solidified my commitment to pursuing a career as a physician. |
| Experience | Please write an essay of about 700 characters for my medical school application, explaining my experiences as an emergency room volunteer. | ChatGPT 3.5 | Volunteering in the emergency room has been a transformative experience for me. As a volunteer, I've had the opportunity to witness firsthand the fast-paced and dynamic environment of the ER. From assisting with patient intake to providing comfort and support to individuals in distress, I've gained valuable insight into the challenges and rewards of emergency medicine. This experience has reinforced my passion for healthcare and deepened my desire to pursue a career as a physician. It has also honed my communication and interpersonal skills, allowing me to effectively interact with patients, families, and healthcare professionals. Volunteering in the emergency room has been both humbling and inspiring, and I am grateful for the opportunity to contribute to the well-being of others in such a meaningful way. |
| Experience | Please write an essay of about 700 characters for my medical school application, explaining my experiences as an emergency room volunteer. Please include at least one specific story of a patient I met. | ChatGPT 3.5 | Volunteering in the emergency room has been a profoundly impactful experience that has deepened my understanding of patient care and reinforced my commitment to medicine. One particular encounter stands out vividly in my memory. I met a middle-aged woman who had arrived at the ER with severe chest pain. As a volunteer, I assisted the medical team in preparing her for evaluation and treatment. Despite her distress, she remained remarkably composed, displaying resilience and courage in the face of uncertainty. Witnessing her strength and determination left a lasting impression on me, reminding me of the profound impact healthcare providers can have on patients' lives. This experience reinforced my passion for medicine and underscored the importance of compassion, empathy, and effective communication in patient care. As an aspiring physician, I am committed to upholding these values and making a meaningful difference in the lives of others. Volunteering in the emergency room has provided me with invaluable clinical exposure and has reaffirmed my dedication to serving others through the practice of medicine. |
| Personal Comments | Please write a Personal Comments essay for my medical school application. | Claude 3 Sonnet | From a young age, I have been driven by an intense curiosity about the human body and mind and a deep desire to help others. My path toward a career in medicine began when my younger brother was diagnosed with Type 1 diabetes at age 7. Watching him endure the difficulties of managing this chronic condition made me keenly aware of the challenges patients face and the incredible impact healthcare providers can have in alleviating suffering.   This experience opened my eyes to the marvels of the human body and sparked my fascination with biology and the life sciences. In high school, I immersed myself in advanced science courses and participated in a medical explorer program at the local hospital. Witnessing surgeries, examining laboratory specimens, and learning from physicians reinforced my determination to become a doctor and care for patients.   At university, I pursued a rigorous curriculum in molecular biology with the goal of gaining a comprehensive understanding of human health at the cellular level. My undergraduate research in cancer biology allowed me to contribute to the relentless pursuit of medical knowledge. However, I realized that as rewarding as lab work can be, my true calling is the human connection cultivated through patient care.  During my gap year, I worked as an emergency medical technician, providing support during ambulance transports and obtaining first-hand experience with acute patient scenarios. This role solidified my passion for the medical field and gave me the opportunity to provide compassionate care during vulnerablehealth crises. I developed skills in calmly managing high-stress situations, thinking critically to determine the best course of action, and displaying empathy when patients needed it most.  The multifaceted journey I have taken has prepared me to embrace the challenges of a medical education and career. From my early exposure to health issues through my brother's diagnosis, to my intensive study of the biological sciences, to my pre-professional experiences providing direct patient care - I have cultivated the vital skills and driving motivation to flourish as a physician. I am committed to the lifelong learning process required to become an exemplary clinician skilled in state-of-the-art practices. Most importantly, I will embrace my role of advocating for my patients and providing them with emotional support during their times of need. The opportunity to combine my scientific curiosity with my calling to serve others is a profound privilege that I wholeheartedly wish to pursue. I am thrilled by the prospect of gaining the well-rounded medical training your institution can provide as I advance toward this deeply meaningful career. |
| Personal Comments | Please write a Personal Comments essay for my medical school application. There is a limit of 5,300 characters. Don't mention the name of any specific school. | Claude 3 Sonnet | As a child, I was enthralled by the complex machinery of the human body. While other kids strived to become astronauts or firefighters, I knew I was born to pursue medicine. This passion took root when my grandfather was diagnosed with Parkinson's disease. Witnessing his indomitable spirit in confronting this debilitating condition sparked my determination to help patients persevere through health crises with the same strength and resilience.  In high school, I volunteered at a nursing home, forming connections with residents and gaining exposure to the realities of patient care. Interacting with Alzheimer's patients revealed the challenges of providing empathetic support amidst cognitive impairments. However, witnessing the tenderness with which nurses attended to residents inspired my commitment to cultivating the same compassionate presence.   At university, I immersed myself in the biological sciences, captivated by the molecular underpinnings of human physiology. While conducting cancer biology research, I took pride in contributing to the relentless quest for medical understanding and striving to enhance patient outcomes. Yet, I continually felt called to directly apply my knowledge at the bedside.  During the summer before my senior year, I participated in a global health initiative in rural Ecuador. Living alongside an indigenous community and providing basic medical services illuminated my role in advocating for underserved populations. Triaging patients and coordinating their access to healthcare was a humbling responsibility that deepened my commitment to promoting health equity.  After graduating, I gained invaluable clinical insight while scribing for physicians in the emergency department. Observing their management of acute care situations - from trauma to stroke - inspired my respect for the seamless integration of medical knowledge and rapid clinical decision-making required in such high-stakes environments. Through these experiences, I refined my communication skills, learning to convey complex information with clarity to colleagues, patients, and families.   Most recently, as an emergency medical technician, I supported paramedics and provided prehospital care during ambulance transports. Remaining poised under pressure while tending to patients' urgent needs validated my ability to make difficult decisions with lives in the balance. Each gripping call reaffirmed that the path of a physician, though immensely challenging, is a calling that will allow me to combine my scientific curiosity and devotion to human service.  My varied roles across healthcare settings have revealed the multifaceted preparation required to fully understand patients' experiences and address their needs with technical excellence and profound empathy. While the prospect of a medical education is daunting, the opportunity to continually learn and grow alongside a community of peers who share my passion is an honor I wholeheartedly embrace. With determination and perseverance, I am confident in my ability to meet the rigorous demands of this training. My life's work is to promote healing by integrating scientific knowledge with compassionate care tailored to patients as individuals. I am thrilled to pursue this deeply meaningful vocation at your esteemed institution. |
| Personal Comments | Please write a Personal Comments essay for my medical school application. Here is the prompt: "Consider and write your Personal Comments Essay carefully; many admissions committees place significant weight on the essay. Here are some questions you may want to consider while writing the essay: Why have you selected the field of medicine? What motivates you to learn more about medicine? What do you want medical schools to know about you that has not been disclosed in other sections of the application? In addition, you may wish to include information such as: Unique hardships, challenges, and obstacles that may have influenced your educational pursuits. Comments on significant fluctuations in your academic record not explained elsewhere in your application." | Claude 3 Sonnet | From my earliest memories, the medical field has captivated my curiosity and driven my ambition to unlock the mysteries of the human body. However, my path toward this career has been shaped by unique challenges that have deepened my tenacity and solidified my devotion to this calling.  I was raised by my father, a hard-working immigrant who struggled to make ends meet. With my mother absent from my life, I faced economic hardship and took on substantial responsibilities from a young age to support my family. Working arduous hours at our family restaurant while juggling my academic obligations taught me resilience and discipline that few of my peers experienced. These early lessons fortified my work ethic and drive to defy limitations.   Despite financial constraints, my father enthusiastically nurtured my fascination with science and medicine. Though he could not afford tutors or extracurricular programs, he spent countless hours after his late shifts helping me grasp complex concepts. His unwavering belief in my potential inspired me to persist through struggles. My academic struggles in biology during sophomore year depleted my confidence, but I met with professors to seek guidance and employed time-management strategies that enabled me to raise my performance.   My determination was further fueled by my grandmother's brave battle with breast cancer during my senior year of high school. Witnessing her suffering motivated my commitment to joining the healthcare field. However, providing her care proved immensely challenging for my father. I became her primary daytime caregiver - managing her medications, appointments, household duties, and my own studies. Though this responsibility exacerbated academic obstacles, it instilled profound life perspective. Maintaining my grandmother's comfort during her final months revealed the necessity of patient-centered care and connected me with my deeper purpose.  These formative experiences guided my decision to pursue medical training that will prepare me to enhance patient outcomes through scientific excellence and empathic understanding. In college, I balanced microbiology research with volunteering as an emergency medical technician to gain clinical exposure. Managing intense course loads while upholding my commitment to community service demanded meticulous time management and prioritization skills.   My journey has been arduous, but every obstacle has fortified my determination to become a compassionate physician who uplifts marginalized patients facing systemic barriers. The medical field's constant evolution across scientific and technological frontiers ignites my intellectual passion. Yet more than satisfying curiosity alone, I am driven by the unparalleled privilege of alleviating suffering and providing patients with a supportive advocate.  While the path ahead will remain challenging, I have developed perseverance forged by adversity. My unique experiences have prepared me to embrace the demands of rigorous medical training and apply my diverse skills to collaboratively pioneering innovative care approaches. With resilience molded by hardship, I am emboldened to pursue this multifaceted vocation of serving others through the seamless integration of scientific knowledge and humanitarian compassion. |
| Personal Comments | Please write a Personal Comments essay for my medical school application. I would like to emphasize my dedication to reducing health inequities. There is a limit of 5,300 characters. Don't mention the name of any specific school. | Claude 3 Sonnet | From an early age, I was instilled with a steadfast commitment to promoting justice and alleviating inequities. Having witnessed the barriers my immigrant family faced in accessing quality healthcare, I was exposed to the detrimental impacts of health disparities on underserved communities. This formative experience solidified my determination to affect meaningful change as a physician-advocate.  During my undergraduate studies, I was disturbed to learn the staggering statistics surrounding healthcare inequities. The disproportionate burden of chronic illnesses, lack of preventive care access, and overall poorer health outcomes among racial minorities, low-income populations, and other marginalized groups revealed a dire need to address systemic injustices. Fueled by a fervent sense of social responsibility, I immersed myself in public health initiatives aimed at advancing health equity.  As a volunteer with a mobile health clinic, I provided free medical screenings to uninsured individuals in urban and rural communities. Collaborating with interdisciplinary teams to coordinate client resources and health education illuminated the necessity of culturally competent, holistic care models. I was humbled to forge connections with patients facing immense obstacles and witness their resilience in combating daunting socioeconomic determinants of health.   My involvement with this initiative inspired me to join a student organization dedicated to expanding healthcare access for immigrant populations. In this role, I facilitated transportation solutions, language interpretation services, and patient outreach to empower our region's burgeoning refugee communities and help bridge gaps in the health system. These experiences revealed the profound efficacy of community-based programs that address the unique needs of underprivileged groups from a grassroots approach.  Determined to deepen my understanding of health policy's impact, I pursued an interdisciplinary research project that examined racial and socioeconomic disparities in pediatric asthma treatment within our state. Analyzing population health data and stakeholder interviews underscored how multiple systemic barriers perpetuate cyclical disadvantages for vulnerable demographics. Presenting our team's findings energized my commitment to catalyzing sustainable, evidence-based reforms.  After graduating, I sought international perspectives by serving with the Peace Corps in South America. Living alongside an indigenous community and delivering basic medical services reinforced my global understanding of healthcare injustices while fostering my communication skills for bridging cultural divides. Training local health educators and coordinating maternal/child wellness initiatives illuminated the power of capacity-building approaches that cultivate lasting self-sufficiency.  Through each of these experiences, I have cultivated a multipronged perspective on advancing health equity that integrates direct service, community engagement, research/policy, and cross-cultural understanding. Alleviating inequities demands embedding this ethos as a core principle - seeking to dismantle oppressive systems through clinical care, public health advocacy, interdisciplinary collaboration, and inclusive decision-making.  The path ahead will undoubtedly present new challenges as I strive to create meaningful change. However, I am emboldened by my personal history and the enduring impact I have already witnessed through concerted efforts to uplift marginalized populations. With proven dedication, I am eager to embrace the rigorous medical training that will equip me to honor my calling as an adept physician and tireless champion for equitable access to the highest quality healthcare services for all communities. |
| Personal Comments | Please write a Personal Comments essay for my medical school application. I would like to emphasize my dedication to biomedical research, based on my experiences doing microbiology research as an undergraduate. | Claude 3 Sonnet | From constructing crude model cells in elementary school to exploring the intricacies of microbial life through a microscope lens, my curiosity about the invisible world within us has captivated me for as long as I can remember. This innate fascination blossomed into a driving passion for biomedical research during my undergraduate years – a passion that has solidified my path toward a career as a physician-scientist.  My first foray into laboratory research was analyzing bacterial virulence factors and antibiotic resistance mechanisms. I vividly recall the thrill of witnessing colonies of Staphylococcus aureus rapidly depleting my antibiotic samples through their adaptive abilities. This glimpse into the remarkable evolutionary arms race between microbes and modern medicine sparked an intellectual obsession. I became enthralled unraveling the molecular mechanics underpinning pathogenesis, defence mechanisms, and emerging therapeutic avenues.  This experience affirmed my decision to pursue an intensive Honors microbiology thesis project investigating novel antimicrobial compounds. After months of scrutinizing literature, designing experiments, and interpreting data, I successfully identified a promising new antibiotic candidate derived from soil bacteria. Presenting my findings at national conferences alongside pioneering researchers in my field was an unparalleled privilege that deepened my confidence and cemented my commitment to this domain of study.  Beyond enriching my scientific acumen, these research endeavors revealed the extraordinary potential of biomedical innovation to revolutionize patient care and address urgent public health crises like antimicrobial resistance. However, I also recognized that my role would be incomplete without the ability to directly apply these advances through clinical practice. Shadowing physicians in infectious disease and microbiology specialties, I admired how their scientific mastery facilitated life-saving diagnoses and treatment plans at the bedside.  This convergence of bench and bedside medicine solidified my aspiration to lead a career dedicated to catalyzing biomedical progress. As a physician-scientist, I will be positioned to spearhead cutting-edge research that elucidates the molecular foundations of health and disease, while simultaneously improving lives through the delivery of exemplary patient care. My undergraduate research credentials have prepared me to embrace the rigorous demands of academic medicine and generate novel insights that propel the field forward.  The opportunity to confront medicine's most complex scientific frontiers both in the lab and clinic is an immense privilege and driving force. With relentless intellectual curiosity, determined leadership, and a steadfast commitment to innovation, I will tirelessly pioneer creative solutions to society's most pressing health challenges. I am eager and well-equipped to pursue the physician-scientist pathway that will empower me to craft an enduring legacy of lifechanging discoveries and impacts. |
| Personal Comments | Please write a Personal Comments essay for my medical school application. I would like to emphasize my experience caring for my grandmother, who had Alzheimer's disease. There is a limit of 5,300 characters. Don't mention the name of any specific school. | Claude 3 Sonnet | A wistful melody danced through the hallway as I entered my grandmother's home each afternoon after school. Despite her fading memories, influenced by the relentless progression of Alzheimer's disease, music remained an immutable connection to her essence. Spending my formative years as my beloved grandmother's caregiver became the most profound experience that solidified my path to medicine.  The early signs were subtle - misplaced belongings, forgotten conversations, bouts of confusion. As her cognitive decline accelerated, I bore witness to this ruthless neurodegenerative disease dismantling her vibrant spirit. My family struggled to find adequate care resources, so I became her primary companion - managing medications, appointments, meals, and personal care needs. Though exhausting, I cherished our poignant moments of preserved selfhood when her eyes sparkled with recognition.  Escorting my grandmother through this dignifying vale of twilight care instilled an invaluable perspective. I developed patience and empathy as I learned to approach each interaction as an opportunity to uphold her comfort and autonomy despite her limitations. When communication became fragmented, I creatively employed visual cues, gentle touch, and soothing tones to foster moments of connection amidst the turmoil.  My role transcended physical caretaking; I became a steadfast advocate. Navigating medical bureaucracies, researching potential treatment plans, and assertively coordinating her care team galvanized my responsibility as her partner in health. I witnessed physicians' inconsistent ability to cultivate therapeutic rapport with patients crippled by cognitive impairments. This inspired my commitment to delivering patient-centered care that preserves dignity and honors humanity.  As her condition deteriorated over four years, I grappled with profound emotional turmoil. Mourning her incremental losses while cherishing her dwindling presence challenged me to develop resilience, emotional intelligence, and capacities to withstand grief. This harrowing journey illuminated both the agonizing impacts of neurodegenerative afflictions and the sacred privilege of being present during life's final chapters.  Despite innumerable hardships, my devotion transcended obligation - it was an act of unconditional love. In my grandmother's most sublime moments shielded from the tangled distortions of disease, she emanated wholehearted appreciation. Her enduring expression of gratitude for the care I provided became the resounding call beckoning me to promote human welfare as a physician.  My experiences cultivated fortitude, perspective, and resolute compassion that will undoubtedly imbue my medical training. While the challenges of healthcare are continually mounting, my path has solidified perseverance rooted in the deepest wellsprings of empathy. I have been honored and fortified by an extraordinary awakening to the sanctity of personalizing care for each patient's humanity. With a renewed exaltation of medicine's capacity to preserve dignity across all phases of life, I can synthesize scientific mastery and humanitarian purpose to serve the patient's utmost healing - an uncompromising vocation I wholeheartedly embrace. |
| MD-PhD Motivation | Please write an essay for my application to MD-PhD programs, describing my reasons for pursuing the combined MD-PhD degree. This essay is limited to 3,000 characters. | Claude 3 Sonnet | My pursuit of the MD-PhD degree stems from an unwavering passion to unite scientific discovery with clinical excellence in order to catalyze translational advances that revolutionize patient care. From my earliest experiences, I have been driven by an insatiable curiosity to unravel the molecular intricacies underlying human health and disease. However, this intellectual obsession is complemented by an equally fervent commitment to directly applying biomedical insights through compassionate clinical practice.  As an undergraduate researcher, I was captivated investigating novel antimicrobial compounds to combat the escalating crisis of drug resistance. Pioneering therapeutic avenues by elucidating the mechanisms of bacterial pathogenesis and immune defense signaling pathways immersed me in the challenges and triumphs at the vanguard of biomedical innovation. Presenting my findings at national conferences cemented my confidence to spearhead high-stakes research with the potential to transform treatment paradigms.   Simultaneously, I engaged in clinical shadowing experiences that revealed how even the most groundbreaking discoveries remain theoretical until seamlessly translated into enhanced patient outcomes through skilled diagnostic reasoning and care delivery. Observing healthcare teams synergize their interdisciplinary expertise to formulate individualized treatment plans illuminated my obligation to fuse comprehensive scientific mastery with a humanistic, collaborative approach.   The pivotal synthesis of these complementary trajectories occurred while volunteering in an oncology lab. Studying novel targeted therapies and tumor profiling techniques, I recognized the unparalleled power of precision medicine to offer hope by leveraging an exquisite understanding of pathophysiology and genomics. Yet I also witnessed the immense emotional turbulence patients endure as they confront existential uncertainty. This underscored how the most pioneering interventions are only as impactful as the compassionate context in which they are administered.  My conviction to pursue the unified investigator and clinician roles was further solidified through collaborating with a pediatric oncologist conducting gene therapy trials. Our intellectual exchange revealed the dynamic synergies of biomedical and clinical insight – how molecular discoveries emanate from astute clinical observations, while novel therapies arise from painstaking bench research. This continuous pathway of dynamic workflow, spanning laboratory innovations to their clinical applications and back again, encapsulates the systemic approach I aim to embody.   The comprehensive training of the MD-PhD program will empower me with a unique perspective transcending boundaries and holistically shaping the future of medicine. With rigorous preparation encompassing both empirical research skills and foundational clinical acumen, I will be equipped to catalyze progress at every pivotal juncture where groundbreaking inquiry can be transformed into paradigm-shifting care modalities. At the forefront of medicine's evolution, the physician-scientist's role synthesizes not only science and practice, but human empathy and leadership in cultivating multi-disciplinary collaborations.  My ambition extends beyond achieving personal career milestones – I am driven by a higher calling to marshal the immense power of scientific understanding to uplift community health and resilience on a global scale. Armed with the distinctive training to navigate biomedical inquiry, clinical implementation, and dynamic cross-disciplinary exchange, I will advance medicine's frontier into an era of unprecedented possibility. |
| MD-PhD Motivation | Please write an essay for my application to MD-PhD programs, describing my reasons for pursuing the combined MD-PhD degree. I am interested in studying neurodegeneration. This essay is limited to 3,000 characters. | Claude 3 Sonnet | From the moment I peered through my first microscope and glimpsed the intricate architecture of neurons, I was captivated by the enigmatic complexity of the brain. This fascination has only intensified through witnessing firsthand the devastating toll of neurodegenerative diseases like Alzheimer's dementia. My experiences have solidified an unwavering determination to unravel the molecular underpinnings of neurological afflictions and pioneering innovative therapeutic strategies through the synergistic training of the MD-PhD pathway.  Alzheimer's disease has been a formidable presence throughout my life, shaping both my personal narrative and professional aspirations. As my grandmother's primary caregiver, I bore witness to this insidious condition's relentless assault on her vibrant personhood. While current symptomatic treatments offered negligible respite, watching her succumb to cognitive impairments ignited an visceral drive to preemptively combat the pathological processes underpinning neurodegeneration.  This catalyzed my intense undergraduate research pursuing the characterization of misfolded amyloid-beta proteins and their disruption of synaptic signaling pathways. I became enthralled investigating the molecular neurobiology implicated in Alzheimer's pathogenesis, from mitochondrial dysfunction to inflammatory cascades. Presenting my findings elucidating novel gene expression biomarkers at national conferences solidified my commitment to elucidating disease mechanisms that could unlock new therapeutic targets.  However, I recognized that translating groundbreaking discoveries into clinical impact hinges on an intimate understanding of disease manifestations and patient experiences. By shadowing neurologists and participating in caregiver support initiatives, I cultivated profound perspective. Cultivating therapeutic rapport with dementia patients revealed the necessity of personalizing care approaches and advocating for their dignity. These diverse experiences were an awakening, revealing how embodying the roles of both investigator and clinician synergistically enhances the capacity to catalyze progress from multiple vantages.  My vision crystallized while completing an Honors thesis that developed novel in vivo techniques for longitudinally monitoring synaptic integrity in Alzheimer's mouse models. This multi-modal experimental design allowed me to integrate recently elucidated molecular pathways with their correlated neuropathological and behavioral phenotypes. Our findings identified a promising therapeutic window when synaptic biomarkers first manifested prior to cognitive decline. Observing the dynamic continuum from molecular inquiry to preclinical therapeutic development underscored the unique potential of the MD-PhD training to accelerate the bench-to-bedside research pipeline.  The opportunity to unite comprehensive clinical training with specialized scientific expertise positions the physician-scientist at the vanguard of biomedical innovation. As neurodegenerative disease prevalence escalates into a public health crisis, the urgency of multi-disciplinary collaboration has never been greater. By harmonizing clinical insight with cutting-edge laboratory investigation, I will spearhead developments transcending traditional boundaries. With a holistic perspective spanning molecular neurobiology, translational studies, clinical therapeutic applications, and patient-centered care advocacy, I can maximize the translatability and real-world impacts of breakthrough discoveries.   My inspirational personal narrative has fortified an unyielding perseverance to uplift vulnerable patient populations. However, my motivation extends beyond an individualistic pursuit - I am driven by an obligation to society and universal human dignity. Approaching this vocation with reverent humility, I will devote my career to pioneering paradigm-shifting advancements that transform our understanding and treatment of devastating neurodegenerative conditions. It is an honor to embrace this pivotal charge and converge scientific ingenuity with human compassion at the forefront of modern medicine's revolution. |
| MD-PhD Motivation | Please write an essay for my application to MD-PhD programs, describing my reasons for pursuing the combined MD-PhD degree. I am interested in studying cancer. This essay is limited to 3,000 characters. | Claude 3 Sonnet | From an early age, I have been captivated by the resilience of the human body and its intricate cellular mechanisms. However, this profound appreciation was galvanized when I lost my grandmother to an insidious battle with ovarian cancer. Bearing witness to her anguished suffering ignited an unwavering determination to unravel the mysteries of oncogenesis and pioneer novel therapeutic strategies. The singularly transformative potential of the MD-PhD pathway will empower me to spearhead the vanguard of cancer research while providing personalized, compassionate care.  My grandmother's cancer odyssey was my first immersion into the visceral realm of translational medicine. From her harrowing diagnosis and the daunting complexity of evaluating treatment regimens, to bouts of harsh cytotoxicity and the agonizing toll of disease progression - I developed an intimate understanding of patients' perspectives that remains etched into my psyche. This seminal experience fortified my resilience and resolve to mitigate the staggering burden of malignancies.  In the laboratory, I became obsessed with elucidating the cellular biology and genetic aberrations underlying tumorigenesis. During my undergraduate thesis, I investigated novel tumor metabolic pathways regulating cancer stem cell quiescence and metastatic potential. Our research identified promising new therapeutic avenues for disrupting these pro-tumorigenic processes. Presenting our high-impact findings at international conferences immersed me in the dynamic vanguard of oncology innovation. I was invigorated by the potential of integrating multidisciplinary approaches spanning genomics, metabolomics, systems biology, and pharmacology.  While these scientific insights unlocked new realms of potential, my grandmother's struggle illuminated how their impact remains constrained without astute clinical acumen bridging discoveries to bedside interventions. Shadowing oncologists, I admired how seamlessly they harmonized evidence-based medicine with an individualized, holistic approach to each patient's comprehensive care. I recognized that embodying this duality as a physician-scientist would empower me to catalyze progress at every pivotal juncture along the translational continuum.  My experiences culminated in an Honors thesis project developing patient-derived xenograft models to evaluate personalized therapies targeting specific tumor profiles. This human-centered experimental design reflected my commitment to merging scientific ingenuity with patient-oriented principles. By longitudinally investigating therapeutic responses and tumor evolutionary dynamics, I explicated critical considerations bridging high-throughput screening data with clinically implementable treatment schemas. These insights revealed how multidimensional MD-PhD training could accelerate oncology's evolution.  With comprehensive expertise spanning molecular oncogenesis to precision medicine implementation, I will spearhead paradigm-shifting discoveries that revolutionize our understanding while advancing tailored interventions and prevention strategies. By embodying the seamless investigator-clinician synergy, I can harness the full continuum of biomedical innovation - formulating novel hypotheses from clinical observations, accelerating discoveries through rigorous experimentation, and facilitating their rapid translation into enhanced patient outcomes.  My motivation is fundamentally rooted in uplifting the human condition. The rippling impacts of cancer transcend individual loss - reverberating across families, communities, and future generations. Approaching this vocation with profound humility, I am driven by an existential responsibility to society and the steadfast belief that every person deserves to confront this malignant scourge with hope and dignity. As a physician-scientist, I can uniquely converge empirical investigation with an enduring compassion for the sanctity of life. Armed with this comprehensive perspective, I will relentlessly pioneer discoveries that transform our understanding, treatment, and ability to minimize cancer's indelible suffering. It is the utmost honor and privilege to merge boundless scientific potential with an empathic commitment to restoring health and resilience. |
| MD-PhD Motivation | Please write an essay for my application to MD-PhD programs, describing my reasons for pursuing the combined MD-PhD degree. I am interested in studying autoimmunity. This essay is limited to 3,000 characters. | Claude 3 Sonnet | From a young age, I have been instilled with a profound reverence for the human body's intricate mechanisms of preserving physiologic homeostasis. However, my appreciation was galvanized when an autoimmune disorder disrupted this delicate equilibrium within my own family. Witnessing my mother's prolonged battle with rheumatoid arthritis - her anguish, the treatment odysseys, and the perpetual uncertainties - crystallized my driving ambition to unite scientific inquiry with medical excellence. By embodying the distinctive roles of investigator and physician through the transformative MD-PhD pathway, I can spearhead novel insights into the pathogenesis of autoimmunity while pioneering enhanced therapeutic strategies.  The cadence of my childhood was marked by my mother's cyclic disease flares - her swollen joints, debilitating pain, and overwhelming fatigue became ingrained into my earliest memories. As I matured, I cultivated an intimate understanding of autoimmune conditions' mercurial, systemic impacts on patients' lives. Her waning mobility restricted our family activities. The barrage of immunosuppressants and biologics buried her in a haze of adverse effects. Yet, the most profound toll was bearing witness to her suffering metaphysical anguish of having her own body's defenses turned against itself.  These indelible experiences solidified my aspiration to elucidate the mechanisms driving pathologic autoimmunity and immune dysregulation at a fundamental biological level. My undergraduate research deciphering cell signaling pathways governing lymphocyte activation, differentiation, and inflammatory cascades immersed me in the challenges vexing this dynamic frontier of translational immunology. Delivering presentations at national conferences energized my passion while affirming my potential to pioneer groundbreaking inquiry.  While these scientific explorations enriched my understanding, witnessing my mother's clinical trajectory illuminated how actualizing discoveries into enhanced therapies and individualized care regimens requires comprehensive medical training. Through shadowing rheumatologists, I appreciated how their management of complex autoimmune cases harmonized evidence-based protocols with nuanced clinical acumen accounting for patients' circumstances. I was awakened to the privilege of embodying this distinctive hybrid role fusing investigative zeal with compassionate humanism.  My commitment was actualized during my Honors thesis research, where I analyzed aberrant regulatory T cell function in experimental models of autoimmune hepatitis. By integrating multi-omics approaches spanning genomics to functional immunophenotyping, we uncovered potential novel biomarkers and immunomodulatory therapeutic targets. This immersive project elucidated how the singular билд of the physician-scientist is indispensable for bridging fundamental insights into the clinical contexts where their impacts are manifested.  With rigorous preparation bridging basic science and therapeutic applications, I will be distinctly poised to spearhead pioneering insights across autoimmunity's multifaceted paradigm. My comprehensive expertise can catalyze progress by formulating novel hypotheses based on clinical patterns, accelerating discoveries through multi-dimensional investigation, and facilitating rapid translation into enhanced preventative strategies, diagnostics, and personalized interventions. This continuous bench-to-bedside-to-bench cycle will empower me to fulfill autoimmunity research's boundless potential.  My deepest motivations, however, transcend achieving career milestones - they are anchored in uplifting the human condition. The devastating physical, emotional, and societal tolls of autoimmune disorders reverberate across families and communities. Yet approaching this arduous path with profound humility, I am emboldened by the enduring opportunity to harmonize empirical exploration with an empathic promise to alleviate suffering and restore hope. With determination fueled by personal resonance, I will devote my career to pioneering paradigm-shifting advances that revolutionize our fundamental understanding while transforming therapeutic paradigms. Embodying the distinctive privileges of the physician-scientist, I can uniquely converge boundless scientific potential with a compassionate commitment to preserving health, resilience, and dignity for all. |
| Failure Essay | Please write an essay for my medical school application. The prompt is: "Describe a time or situation where you have been unsuccessful or failed." The essay should be around 250 words long. | Claude 3 Sonnet | The clinical skills course was one of the most demanding challenges of my first year of medical school. As a student keen on pursuing surgery, I was determined to excel at procedures like suturing, IV insertion, and chest tube placement. However, my unmitigated struggle to master intubation became an unshakeable obstacle.  No matter how meticulously I studied the technique, my practical attempts were plagued by delays in visualization, improper positioning, and difficulties advancing the endotracheal tube. With each failed intubation attempt came a sinking sense of frustration and doubt clouding my confidence. Despite my peers' suggestions and the professors' coaching, the skills simply refused to click. I found myself repeatedly sideswiped by an unforgiving Learning curve.  My inability to successfully intubate a manikin before the exam deadline was profoundly humbling. Facing this highly structured competency, I was forced to confront the limitations of my abilities at that stage. Initially, I spiraled into self-criticism - I felt embarrassment about underperforming compared to my classmates and questioned whether I possessed the dexterity to thrive in procedural specialties.  Ultimately, this failure motivated invaluable self-reflection and resilience. While the technical skills will solidify over years of practice, this experience revealed that I occasionally conflated confidence with arrogance. It instilled a critical lesson in maintaining humility regardless of circumstance. Moreover, rebounding from this setback fortified my determination to persist through future challenges by embracing them as opportunities for growth.  Despite countless practice attempts, I did not achieve intubation mastery that semester. However, failing to clear that particular competency hurdle became one of my most transformative experiences. It tempered my approach with greater realism, diligence, and grit - vital qualities for tackling the innumerable complexities that await in my medical training and career. |
| Failure Essay | Please write an essay for my medical school application. The prompt is: "Describe a time or situation where you have been unsuccessful or failed." Focus on a time when I got a low grade in organic chemistry. The essay should be around 250 words long. | Claude 3 Sonnet | As I eagerly commenced my undergraduate journey towards the pre-medical track, I had confidently underestimated organic chemistry's formidable challenges. This hubris quickly materialized into underprepared struggles during the first semester's daunting course load. Despite relentless efforts, my initial exam fell devastatingly short of expectations, culminating in a failing grade that deeply shook my confidence.  In the aftermath of this humbling failure, I found myself at an inflection point. A deluge of self-doubt flooded my mind - was I truly capable of handling the academic rigor required for medical school? Had I made an erroneous career choice? This crossroads forced me to conduct uncompromising self-reflection about my preparedness, strategies, and determination.  However, succumbing to pessimism was not an option I could accept. With renewed focus, I analyzed where my approach was faltering and obstinately revamped my entire system for absorbing the dense course material. I scheduled sessions with tutors who provided new conceptual models. I relentlessly tested my knowledge through practice problems and metabolic pathway mapping exercises. Gradually, the foundations solidity and I developed effective tools to unravel the complexity systematically.  Beyond evolving my academic strategies, this failure thickened my perseverance and drive. The initial setback instilled humbling perspective about the challenges ahead - to conquer the rigors of medical training, I must eliminate complacency and continually reinvent my efforts to surpass oneraising expectations. This demanding refining fire reforged my resolve to transform indomitable struggles into catalysts for growth.  Although the semester's initial failure could have precipitated disastrous consequences, rebounding from that crucible experience transformed me into a stronger, more versatile scholar. Powering through organic chemistry's grueling gauntlet validated my capacity to thrive amid unyielding academic pressure. Armed with enhanced study skills, intellectual humility, and augmented resilience - I became emboldened to embrace the intense demands awaiting in my medical journey. |
| Failure Essay | Please write an essay for my medical school application. The prompt is: "Describe a time or situation where you have been unsuccessful or failed." Focus on a time when I did not do an effective job planning an activities fair. The essay should be around 250 words long. | Claude 3 Sonnet | As the newly-elected president of my university's pre-medical club, I was eager to make an impact by reinvigorating our presence on campus. One of my first major initiatives was organizing an activities fair to showcase different clinical volunteering opportunities and student groups. However, my lack of foresight and planning uncertainties led this event to falter drastically.  In my zeal to hit the ground running, I grossly underestimated the extensive coordination required for an expansive showcase. Student group registrations and organizational logistics were haphazard. On the day of the fair, there was immense confusion as club representatives showed up to unassigned or cramped table spaces. Programs had printing errors riddling their informational handouts with inaccuracies. Worse yet, disappointing turnout from a lack of advertising left most exhibitors dismayed by the lackluster crowds.  As the afternoon unraveled into escalating disarray, I recognized this event had become a sobering portrayal of failed leadership and planning on my part. Observing exhibitors' Frustrations amid avoidable chaos revealed how carelessly neglecting essential oversight could derail even simple initiatives. This failure forced me to confront glaring deficiencies in my organizational skills and time management strategies.  Though initially disheartening, this humbling experience was a vital inflection point for reforming my approach to tackling complex projects. I meticulously reflected on my missteps, from underestimating preparation timelines to ineffectively delegating responsibilities with my board members. I developed enhanced collaborative tools like detailed checklists, timeline buffers, and communication channels to ensure cohesive execution.  More importantly, rebounding from this failure instilled an urgency to sharpen oft-neglected leadership competencies. I sought mentorship on managing group dynamics, public speaking, and acting decisively amid constant flux. These refinements have been invaluable for orchestrating high-stakes situations with poise and inspiring enduring team investment.  While the activities fair did not achieve its intended vision, this humbling missed opportunity catalyzed profound growth in my project stewardship abilities. Embracing mistakes as an impetus for elevating my potential has been an integral driver of my personal and professional maturation. The resilience cultivated through this crucible experience solidified vital capacities for navigating my future as an adaptable, prepared, and effective leader. |
| Failure Essay | Please write an essay for my medical school application. The prompt is: "Describe a time or situation where you have been unsuccessful or failed." Focus on a time when I did not support my friend when her mother was sick. The essay should be around 250 words long. | Claude 3 Sonnet | During my sophomore year of college, my close friend Samantha was struggling immensely as her mother battled a devastating illness. While I was vaguely aware of the situation, I failed to appreciate the magnitude of Samantha's emotional turmoil and neglected to serve as a supportive, reassuring presence when she needed it most.  When Samantha began missing classes and group outings, I arrogantly misinterpreted her absence and distance as unreliable flakiness. I did not make any effort to check in or offer assistance, consumed by my own social ambitions and academic responsibilities. Meanwhile, Samantha was consumed with anguish, grappling with her mother's grim prognosis and deteriorating condition.  My obliviousness to her dire circumstances persisted until I encountered Samantha emotionally distraught in the library one evening near the semester's end. Only then did the gravity of her situation become apparent - her mother had been given only weeks to live. Wracked with guilt and profound regret, I realized how selfishly I had abandoned a friend in crisis out of sheer negligence.  In that moment, I recognized how callously I had failed to uphold the empathy, compassion, and care that should characterize not just a good friend, but an aspiring future physician. I had inconsiderately neglected emotional struggles transpiring around me, exemplifying a lack of insight and inability to intervene proactively. This revelatory lapse exposed glaring deficiencies in my emotional intelligence that necessitated critical improvement and maturation.  Although I strived to provide the emotional support Samantha deserved during the final agonizing months of her mother's life, the damage had been inflicted. Samantha felt deeply let down by my negligence during the most devastating experience she had endured. I vowed to atone for this failure through immense personal growth and intentional outreach work supporting families facing similar turmoil.  This harsh lesson continues fueling my development of comprehensive empathetic skills. I must become adept at identifying emotional undercurrents, engaging in difficult conversations, and serving as a proactive source of compassion. Only through this heightened sensitivity can I ensure each future patient encounters an attentive, caring advocate dedicated to alleviating suffering in all its forms. Failures of friendship can serve as catalysts for continual self-improvement and lifelong investments in empathic humanism. |
| Anything Else | Please write an essay to include in my medical school application. The prompt is: "Is there anything else you would like to share with the Committee on Admissions? Some applicants use this space to describe unique experiences and obstacles such as significant challenges in access to education, unusual socioeconomic factors, and/or identification with a particular culture, religion, race, ethnicity, sexual orientation, or gender identity and/or possibly challenges related to COVID or other issues in preparation for medical school." The essay should be around 250 words long. I would like to focus on how motivated I am to go to medical school in New York. | Claude 3 Sonnet | From my earliest memories, the vibrant diversity and resilience of New York has shaped my identity and inspired my pursuit of medicine. Growing up in a working-class immigrant community in Queens instilled an appreciation for our state's unparalleled multiculturalism and the shared human experiences that transcend boundaries. These formative perspectives fortified my determination to establish roots as a physician dedicated to uplifting the health and wellbeing of all New Yorkers.  The ethnic enclaves surrounding my childhood home exposed me to the staggering scope of socioeconomic and health disparities across our city's melting pot. I vividly recall the dichotomy between families in my neighborhood struggling to access basic preventative care while high-tech medical facilities were just miles away. This foundational awareness catalyzed my empathetic drive to address systemic barriers obscuring equitable care for underserved populations through compassionate community engagement.  During my undergraduate studies, I embraced opportunities to serve New York's diverse patient communities. As a volunteer emergency medical technician, I supported ambulance crews throughout the five boroughs, providing culturally competent care during high-acuity crises. These experiences solidified my commitment to delivering patient-centered approaches tailored to each individual's unique circumstances and health beliefs.  My profound admiration for the grit and tenacity embodied by New Yorkers has only intensified through recent hardships. Witnessing the collective resilience displayed amid existential challenges like the COVID-19 pandemic underscored our communities' unbreakable perseverance. It is a profound privilege to train among the nation's preeminent institutions while living amid the unwavering spirit that defines this city's essence.  With steadfast passion rooted in my personal narrative, I am wholeheartedly dedicated to earning a medical education in New York. This vibrant nexus of cultures, economic backgrounds, and life philosophies exemplifies the diversity I aspire to honor through an enduring career of advocacy and service. By establishing myself as a skilled, socially-conscious physician, I can uplift the health trajectories of neighborhoods like the one that shaped my vocational calling. New York's perpetual resilience deserves to be safeguarded through exceptional care accessible to all - a vision I am relentlessly committed to realizing. |
| Anything Else | Please write an essay to include in my medical school application. The prompt is: "Is there anything else you would like to share with the Committee on Admissions? Some applicants use this space to describe unique experiences and obstacles such as significant challenges in access to education, unusual socioeconomic factors, and/or identification with a particular culture, religion, race, ethnicity, sexual orientation, or gender identity and/or possibly challenges related to COVID or other issues in preparation for medical school." The essay should be around 250 words long. I would like to focus on how I grew up in poverty and received free lunch at school. | Claude 3 Sonnet | Growing up, my family struggled to make ends meet. We relied on food stamps and the free lunch program at school to have enough to eat. My parents worked multiple jobs with unstable hours, making it difficult to provide constancy and supervision at home. From an early age, I had to be self-sufficient and responsible beyond my years. Despite the challenging circumstances, my parents instilled in me the importance of education as the path to a better life. They made innumerable sacrifices to create opportunities they never had. Seeing their struggles motivated me to excel academically in hopes of a career that could lift my family out of poverty. I took advanced classes, participated in extracurriculars, and worked part-time jobs to build an admissions profile worthy of scholarships. My efforts paid off as I became the first in my family to attend college on a full academic scholarship. The transition to university was not easy - I often felt out of place among more affluent peers who never knew material hardship. However, I persevered by staying focused on my goals. My background fuels my drive to not just be a doctor, but one serving under-resourced communities. I am intimately familiar with the challenges of poverty and its impacts on health. With an M.D., I can give back by providing affordable, culturally-competent care for those most in need. My diversity of experience will enable me to relate to and effectively treat patients from all walks of life. |
| Anything Else | Please write an essay to include in my medical school application. The prompt is: "Is there anything else you would like to share with the Committee on Admissions? Some applicants use this space to describe unique experiences and obstacles such as significant challenges in access to education, unusual socioeconomic factors, and/or identification with a particular culture, religion, race, ethnicity, sexual orientation, or gender identity and/or possibly challenges related to COVID or other issues in preparation for medical school." The essay should be around 250 words long. I would like to focus on being a member of an under-represented minority group (I am Latino). | Claude 3 Sonnet | As a Latino from an immigrant family, I have faced unique challenges in my pursuit of higher education and a career in medicine. However, my cultural background has also imbued me with invaluable strengths - resilience, empathy, determination - that will make me an exceptional physician. My parents came to the United States with limited formal education, seeking better opportunities. Though they worked tirelessly in low-wage jobs to provide for our family, financial struggles and linguistic barriers made navigating educational systems difficult. From an early age, I had to advocate for myself academically and translate for my parents at school meetings. These obstacles made me even more driven to take advantage of every opportunity. I immersed myself in extracurriculars and summer programs to build an impressive profile for college admissions. As a first-generation student, I lacked role models for achieving my dream of becoming a doctor. Yet my passion persevered as I gained admission to a prestigious university on a scholarship. On campus, I embraced my identity as a Latino leader. I joined associations like the Latin American Student Organization and took on roles mentoring other underrepresented minority students. These experiences deepened my pride in my cultural roots while fueling my commitment to increasing diversity in medicine. I am driven to achieve an M.D. not just for personal success, but to uplift my community. My unique perspective as someone who has overcome underrepresentation will allow me to provide culturally-competent care and serve as an inspiring role model. I will champion health equity as a physician. |
| Experience | Please write a short text for my medical school application, describing my experience being inducted into a nursing honor society. There is a limit of 700 characters. | Claude 3 Sonnet | One of the most humbling experiences of my academic journey was being inducted into the Sigma Theta Tau International Honor Society of Nursing. This prestigious organization recognizes the highest achieving nursing students who embody excellence in scholarship and leadership. To be nominated by faculty and peers was an immense honor that validated my dedication and commitment to the nursing profession. The induction ceremony was a proud moment where I took an oath to uphold the society's values - using knowledge and competence to better human health. As I received the prestigious Sigma Theta Tau pin, I felt a profound sense of responsibility to my calling. This recognition strengthened my passion to become a compassionate, ethical provider of outstanding patient care. The experience motivated me to maintain the highest standards of professionalism on my journey to an M.D. |
| Experience | Please write a short text for my medical school application, describing my experience as a teaching assistant for anatomy and physiology. There is a limit of 700 characters. | Claude 3 Sonnet | As an undergraduate, I had the privilege of working as a research assistant in a biomedical engineering lab investigating the molecular mechanisms of muscle contractility. This opportunity allowed me to apply classroom concepts into authentic scientific inquiry and develop valuable skills for a future physician-scientist career. Under the mentorship of the principal investigator, I learned advanced techniques like protein isolation, SDS-PAGE, and quantitative analysis. I gained experience meticulously following protocols, critically analyzing data, and troubleshooting experimental issues. Each week, I documented updates at group meetings, honing my ability to communicate complex ideas. The most rewarding aspect was contributing to the bigger picture of improving human health. Our work examining the dynamics of muscle contraction and relaxation could shed light on cardiovascular and neuromuscular disorders. Witnessing science's capacity to unravel biological mysteries and develop new therapies solidified my commitment to an M.D./Ph.D. path. This research experience was transformative in applying classroom theory to real-world practice. It instilled in me an appreciation for lifelong scientific curiosity and rigorous experimental methods that will serve me well as a medical scientist. |
| Experience | Please write a short text for my medical school application, describing my experience as a research assistant in a lab studying muscle contractility. There is a limit of 700 characters. | Claude 3 Sonnet | During my undergraduate years, I was fortunate to gain invaluable research experience as an assistant in a biomedical engineering lab focused on the molecular mechanisms governing muscle contractility. This opportunity allowed me to apply classroom concepts to authentic scientific investigation while developing skills crucial for a future career as a physician-scientist. Under the guidance of the principal investigator, I was trained in advanced techniques such as protein isolation, SDS-PAGE gel electrophoresis, and quantitative data analysis. I learned the significance of meticulously following protocols, critically evaluating results, and employing creative problem-solving when experiments faced setbacks. Additionally, I honed my ability to communicate complex scientific ideas by documenting weekly progress updates at group meetings. Most rewarding was the chance to contribute to research with potential for improving human health. Our work examining the dynamics of muscle contraction and relaxation could shed light on cardiovascular diseases, neuromuscular disorders, and lead to new therapeutic interventions. Witnessing science's capacity to deepen our understanding of biological processes and pioneer medical advancements solidified my passion for integrating research within a clinical practice. This formative experience fostered my appreciation for lifelong scientific curiosity, rigorous experimental design, and the collaborative pursuit of knowledge that underpins successful research. It was a transformative stepping-stone preparing me for the equally demanding and rewarding path of the physician-scientist. |
| Experience | Please write a short text for my medical school application, describing my experience being inducted into a nursing honor society. There is a limit of 700 characters plus an additional 1325 characters because this is one of my most meaningful experiences. | Claude 3 Sonnet | One of the most humbling experiences of my academic journey was being inducted into the Sigma Theta Tau International Honor Society of Nursing. This prestigious organization recognizes the highest achieving nursing students who embody excellence in scholarship and leadership. To be nominated by faculty and peers was an immense honor that validated my dedication and commitment to the nursing profession. The induction ceremony was a proud moment where I took an oath to uphold the society's values - using knowledge and competence to better human health. As I received the prestigious Sigma Theta Tau pin, I felt a profound sense of responsibility to my calling. This recognition strengthened my passion to become a compassionate, ethical provider of outstanding patient care. My induction into Sigma Theta Tau marked not just an academic milestone, but a pivotal moment that crystallized my purpose and values as a healthcare professional. The society's mission to celebrate nursing excellence while advancing worldwide health resonated deeply. As part of the honor society, I had the opportunity to volunteer providing health education and screenings in underserved communities. Interacting with patients facing barriers to care due to poverty, lack of insurance, linguistic isolation, and other systemic inequities brought to light how social determinants profoundly impact wellbeing. I was exposed to the harsh realities and human toll of health disparities firsthand. These experiences solidified my commitment to becoming a physician champion for greater health equity. I am driven by Sigma Theta Tau's core values of integrity, inclusiveness, creativity, and collegiality in healing the divisions that preclude equal access to quality care. Through leadership and service, I hope to pay forward the investment this society made in recognizing my potential. Sigma Theta Tau not only honored my academic accomplishments, but more importantly provided a ethical framework for my future work addressing the unmet needs of vulnerable patient populations. This society instilled in me the higher calling of healthcare - to use knowledge, skill, and compassion as instruments of societal change in reducing human suffering across all communities. I carry this mission as a guiding light on my journey to medical school and beyond. |
| Experience | Please write a short text for my medical school application, describing my experience as a teaching assistant for anatomy and physiology. There is a limit of 700 characters plus an additional 1325 characters because this is one of my most meaningful experiences. | Claude 3 Sonnet | Serving as a teaching assistant (TA) for the undergraduate anatomy and physiology courses was one of the most rewarding and formative experiences of my academic journey. In this role, I had the opportunity to not only solidify my mastery of the foundational concepts of human structure and function, but also cultivate skills that will be invaluable as a future physician educator. As a TA, I led weekly review sessions, provided one-on-one tutoring, and assisted the professor with grading and curriculum development. This required a deep comprehension of the material to break down complex topics and explain them in an engaging, easy-to-understand manner for students from diverse backgrounds. I developed creativity in using analogies, models, and visual aids to facilitate learning.  One of the most meaningful aspects of being an anatomy and physiology TA was witnessing my impact on students' educational journeys. I took pride in watching them develop from struggling with the dense terminology to confidently grasping the intricate bodily systems. Of particular fulfillment was guiding pupils who initially lacked a strong academic foundation to overcome their insecurities and thrive in this rigorous course. Many of the students I mentored were the first in their families to pursue healthcare career paths. I made an effort to not only teach the courseload, but also provide a supportive environment to nurture their dreams of becoming future medical professionals. I shared insights into crafting a competitive application, study strategies, and overcoming common pitfalls. For many, I became the role model demonstrating that achieving an advanced degree was possible regardless of background. This experience fostered my skills in clear scientific communication, creative instructional methods, empathetic mentorship, and most importantly - patience. I gained a deeper appreciation for how curating an inclusive, nurturing learning environment can empower students to reach their full potential. As a future physician, I will undoubtedly carry these lessons into developing patient education materials, counseling those under my care, and training the next generation of doctors. Serving as an anatomy and physiology TA allowed me to have a lasting impact beyond imparting scientific concepts. I inspired underrepresented minority, disadvantaged, and non-traditional students to persevere toward careers as healthcare's next leaders and scholars. This is the highest calling I can imagine - democratizing medical knowledge to create a more equitable and diversified field for all populations. |
| Experience | Please write a short text for my medical school application, describing my experience as a research assistant in a lab studying muscle contractility. There is a limit of 700 characters plus an additional 1325 characters because this is one of my most meaningful experiences. | Claude 3 Sonnet | During my undergraduate years, I was fortunate to gain invaluable research experience as an assistant in a biomedical engineering lab focused on the molecular mechanisms governing muscle contractility. This opportunity allowed me to apply classroom concepts to authentic scientific investigation while developing skills crucial for a future career as a physician-scientist. Under the guidance of the principal investigator, I was trained in advanced techniques such as protein isolation, SDS-PAGE gel electrophoresis, and quantitative data analysis. I learned the significance of meticulously following protocols, critically evaluating results, and employing creative problem-solving when experiments faced setbacks. Additionally, I honed my ability to communicate complex scientific ideas by documenting weekly progress updates at group meetings.  My time in the muscle contractility lab was transformative in shaping my aspirations as a physician-scientist committed to fundamental research. One of the most rewarding aspects was witnessing how our work examining the molecular dynamics of muscle contraction and relaxation could have translational applications toward understanding and treating cardiovascular diseases, neuromuscular disorders, and other pathologies. I vividly recall the exhilaration of our team identifying a novel protein interaction that appeared to regulate contractile force generation. Authoring the manuscript describing our experimental methods and results taught me how to distill comprehensive scientific narratives into clear, substantive communications. Presenting our findings at a major conference allowed me to discuss their clinical relevance with established researchers and physicians in the field. Beyond the intellectual stimulation, this research experience provided a window into the essential skillsets and mindsets required to succeed in academic medicine. I gained an appreciation for the grit and perseverance necessary to navigate long timelines, inevitable setbacks, and the patient process of incrementally advancing knowledge. Equally important were the values of scientific integrity, meticulous record-keeping, and intellectual humility to approach investigations with an open mind. Most impactful was witnessing how interdisciplinary collaborations, bringing together diverse expertise from bioengineers, biochemists, physiologists and clinicians, could drive more creative solutions to complex medical puzzles. I formed mentoring relationships that provided a first-hand understanding of the unique challenges and rewards of combining clinical practice with basic science research. This experience ultimately crystallized my decision to pursue an MD/PhD path to become a physician-scientist positioned to translate discoveries at the bench to improved therapies at the bedside. The ability to work at this pivotal interface, catalyzing bi-directional advances between science and medicine, embodies the intellectual and humanistic unity that inspires my passion for this dual career. |
| Experience | Please write an essay of about 700 characters for my medical school application, explaining my experiences as a medical scribe. | Claude 3 Sonnet | As a medical scribe in the emergency department (ED), I had an immersive experience observing the realities of acute patient care. I documented comprehensive patient encounters, procedures, coding and billing - gaining insights into the multifaceted demands of a physician's responsibilities beyond treating illness. This role allowed me to seamlessly embed myself within the ED care team. I experienced firsthand how physicians deftly juggle rapidly triaging critical cases, skillfully managing patient flows, and thoughtfully communicating complex medical information with clarity and empathy. I witnessed the grit and mental stamina required to maintain clinical acumen while making high-stakes decisions amidst chaos. Scribing taught me the importance of developing astute contextualization abilities. Piecing together a patient's full narrative through detailed histories, physical exams, diagnostic results and coded undertones was essential for delivering quality, individualized care. I cultivated skills in medical documentation, clinical priorities, and interprofessional collaboration. Most importantly, this experience solidified my commitment to a career safeguarding patient interests. Advocating for thorough assessments, anticipating concerns, and ensuring comprehension at every step reaffirmed my passion for improving the patient experience through compassionate, transparent healthcare delivery. These lessons will forever guide my approach to the privileged physician-patient relationship. |
| Experience | Please write an essay of about 700 characters for my medical school application, explaining my experiences as an emergency room volunteer. | Claude 3 Sonnet | Volunteering in the emergency department (ED) provided an unparalleled glimpse into the adrenaline-fueled world of acute care medicine. Amidst the organized chaos, I witnessed the incredible teamwork required to triage and stabilize critically ill and injured patients. I was inspired by the ED staff's poise under pressure and tireless efforts to provide rapid, skilled care with compassion. My role afforded a firsthand view into the fast-paced decision making that occurs when seconds are precious. I admired how physicians swiftly performed critical assessments, synthesizing complex diagnostic information to establish prompt treatment plans. I saw the dexterity and resilience needed to pivot between caring for a trauma patient one moment to counseling a family on end-of-life care the next. This experience deepened my appreciation for the situational awareness and prioritization skills paramount in emergency medicine. I observed how ethical patient-centered care guided difficult decisions on allocating limited resources to those most in need. The ability to remain levelheaded while handleing crises humanistically left a profound impact. Interacting with ED patients from all walks of life taught me the significance of cultural competency. Breaking down communication barriers, addressing social factors impacting health, and providing reassurance during vulnerable moments exemplified medicine's highest callings of human compassion. I was inspired to advance health equity and accessibility. These experiences illuminated the tremendous privilege of emergency care and reinvigorated my passion. I am driven to join these front lines as a catalyst for restoring health and rendering aid to all communities during their most acute hours of need. Copy |
| Experience | Please write an essay of about 700 characters for my medical school application, explaining my experiences as an emergency room volunteer. Please include at least one specific story of a patient I met. | Claude 3 Sonnet | Volunteering in the emergency department (ED) provided an unparalleled glimpse into the adrenaline-fueled world of acute care medicine. Amidst the organized chaos, I witnessed the teamwork required to rapidly triage and stabilize critically ill patients. I was inspired by the staff's poise under pressure and tireless efforts to provide skilled care with compassion. One patient whose story profoundly impacted me was an elderly woman who arrived unresponsive and bradycardic. I admired how the team efficiently executed lifesaving interventions while updating the distraught family with empathy. Though she ultimately passed, the physician spent considerable time offering a compassionate presence during their grief. This experience deepened my appreciation for situational awareness and ethical patient-centered care. I observed how these skills guided difficult decisions like prudently allocating limited resources to those with better prognoses. The ability to remain levelheaded while handleing crises humanistically left an impact. Interacting with ED patients from all walks of life taught me the significance of cultural competency. I saw interpreters and social workers address communication barriers, socioeconomic factors impacting health, and provide reassurance during vulnerable moments - exemplifying medicine's highest callings. These experiences illuminated the tremendous privilege of emergency care and reinvigorated my passion. I am driven to join these front lines as a catalyst for restoring health and rendering skilled aid with human compassion to all communities during their most acute hours of need. |
