## Supplemental Table 1 for "Estimating the Prevalence of Generative AI Use in Medical School Application Essays"

**Supplemental Table 1**: Undergraduate major subjects were manually coded by the investigators as science, technology, engineering and math (STEM) versus non-STEM. Each row represents a major or combination of majors self-declared by one or more applicants. There are four sub-tables providing a full list of declared majors classified as STEM, non-STEM, both (possible only for double/triple-majors), or undetermined. Practically oriented majors such as “Clinical professions” and “Healthcare management” were not considered STEM, while social sciences such as economics and sociology were considered STEM.

| **STEM majors** | **Number reporting this major** |
| --- | --- |
| Aerospace Engineering | 2 |
| Anatomy & Cell Biology | 1 |
| Anatomy and Cell Biology | 3 |
| Animal Science | 1 |
| Animal Science; Biochemistry | 1 |
| Anthropology | 24 |
| Anthropology (Medical Anthropology); Health and Human Biology | 1 |
| Anthropology (Social) | 1 |
| Anthropology and Human Biology | 5 |
| Anthropology modified with Biology | 1 |
| Anthropology Modified with History | 1 |
| Anthropology; Biochemistry | 1 |
| Anthropology; Biological Sciences | 1 |
| Anthropology; Biology | 1 |
| Anthropology; Chemistry | 2 |
| Anthropology; Global Health Studies | 1 |
| Anthropology; Neurobiology and Physiology | 1 |
| Anthropology: Global Health and Environment | 8 |
| Anthropology: Global Health and Environment | 1 |
| Anthropology: Global Health and the Environment | 1 |
| Anthropology: Global Health Environment | 1 |
| Anthropology: Human Health and Biology | 1 |
| Anthropology/Global Health | 1 |
| Applied & Computational Mathematics and Statistics | 1 |
| Applied Economics and Management | 1 |
| Applied Math and Statistics; Chemistry | 1 |
| Applied Mathematics | 2 |
| Applied Mathematics and Statistics | 1 |
| Applied Medicine | 1 |
| Applied Psychology | 1 |
| Archaeology; Biochemistry | 1 |
| Astronomy | 1 |
| Astrophysics; Physics | 1 |
| B.S. in Biological Sciences, Research Honors; Biological Chemistry | 1 |
| BA-Psychology; BS-Biochemistry and Molecular Biology | 1 |
| Behavioral Biology; Molecular and Cellular Biology | 1 |
| Behavioral Neuroscience | 7 |
| Behavioral Neuroscience Psychology | 1 |
| Bio-Behavioral Health | 1 |
| Bio-Engineering; Biochemistry | 1 |
| Biochemical and Biophysical Sciences | 1 |
| Biochemistry | 309 |
| Biochemistry - Bachelor of Science | 1 |
| Biochemistry ; Global Health | 1 |
| Biochemistry (Chemistry) | 1 |
| Biochemistry (Medicinal Chemistry) | 2 |
| Biochemistry (Medicinal Chemistry); Biomedical Sciences | 1 |
| Biochemistry & Biomedical Sciences (Health Stream) | 1 |
| Biochemistry & Biophysics | 1 |
| Biochemistry & Cell Biology | 1 |
| Biochemistry & Cellular and Molecular Biology | 1 |
| Biochemistry & Chemical Biology | 1 |
| Biochemistry & Chemical Biology; Medicine, Health, and Society | 1 |
| Biochemistry & Molecular Biology | 15 |
| Biochemistry & Molecular Biology | 1 |
| Biochemistry & Molecular Biology (Pre-Med) | 1 |
| BIOCHEMISTRY & MOLECULAR BIOLOGY; DATA SCIENCE: STATISTICS CONCENTRATION | 1 |
| Biochemistry & Molecular Biology; Economics | 1 |
| Biochemistry & Molecular Biology; Statistics | 1 |
| Biochemistry and Biomedical Sciences | 2 |
| Biochemistry and Biotechnology | 1 |
| Biochemistry and Cell Biology | 6 |
| Biochemistry and Cell Biology; Mathematics; Chemistry | 1 |
| Biochemistry and Cellular and Molecular Biology | 1 |
| Biochemistry and Molecular Biology | 45 |
| Biochemistry and Molecular Biology | 1 |
| Biochemistry and Molecular Biology ; Health and Exercise Science | 1 |
| Biochemistry and Molecular Biology; Chemistry | 1 |
| Biochemistry and Molecular Biology; Health Science | 1 |
| Biochemistry and Molecular Biology; Psychology | 2 |
| Biochemistry and Molecular Biology; Psychology - Neuroscience Track | 1 |
| Biochemistry and Molecular Biology/Biotechnology | 1 |
| Biochemistry and Nutrition; Economics - Political Economy Track | 1 |
| Biochemistry and Nutrition; Mathematics | 1 |
| Biochemistry Honors | 3 |
| Biochemistry with College Honors; Neuroscience with College Honors | 1 |
| Biochemistry with Neuroscience Concentration | 2 |
| Biochemistry-Molecular Biology | 1 |
| Biochemistry, Cell and Molecular Biology | 1 |
| Biochemistry; Anthropology | 2 |
| Biochemistry; Applied Mathematics | 1 |
| Biochemistry; Biochemistry, Molecular and Structural Biology | 1 |
| Biochemistry; Biology | 5 |
| Biochemistry; Biology; Biophysics | 1 |
| Biochemistry; Biology; Microbiology / Bacteriology | 1 |
| Biochemistry; Biology; Pre-Medical | 1 |
| Biochemistry; Biomedical Science | 1 |
| Biochemistry; Biophysics | 5 |
| Biochemistry; Biophysics; Economics | 1 |
| Biochemistry; Chemistry | 4 |
| Biochemistry; Chemistry; Academy for Young Scholars | 1 |
| Biochemistry; Chemistry; Biology | 1 |
| Biochemistry; Clinical Psychology | 1 |
| Biochemistry; Community Health | 1 |
| Biochemistry; Computer Science | 1 |
| Biochemistry; Computer Science + Mathematics | 1 |
| Biochemistry; Economics | 2 |
| Biochemistry; Genetics | 2 |
| Biochemistry; Genetics, Cell Biology and Development | 1 |
| Biochemistry; Global Health | 1 |
| Biochemistry; Healthcare Management | 1 |
| Biochemistry; Honors Program | 2 |
| Biochemistry; Human Biology | 1 |
| Biochemistry; Mathematics | 3 |
| Biochemistry; Microbiology | 1 |
| Biochemistry; Microbiology / Bacteriology | 1 |
| Biochemistry; Molecular, Cellular, and Developmental Biology | 1 |
| Biochemistry; Neuroscience | 5 |
| Biochemistry; Nutrition | 1 |
| Biochemistry; Pharmaceutical Sciences | 1 |
| Biochemistry; Philosophy | 1 |
| Biochemistry; Physics | 1 |
| Biochemistry; Physics; Biophysics | 1 |
| Biochemistry; Plan II | 1 |
| Biochemistry; Plan II Honors | 1 |
| Biochemistry; Psychology | 8 |
| Biochemistry; Public Health | 1 |
| Biochemistry; Quantitative Biosciences and Engineering | 1 |
| Biochemistry; Quantitative Economics | 1 |
| Biochemistry; Sociology | 1 |
| Biochemistry: Emphasis in Molecular Biology | 1 |
| Biochemistry/Microbiology | 1 |
| Biochemistry/Molecular Biology | 2 |
| Bioengineering | 51 |
| Bioengineering | 1 |
| Bioengineering & Biochemistry | 1 |
| Bioengineering; Biology | 1 |
| Bioengineering; Honors Program | 1 |
| Bioengineering: Bioinformatics | 2 |
| Bioengineering: Biotechnology | 2 |
| Bioengineering/Biochemistry | 1 |
| BioHealth Sciences | 1 |
| BioHealth Sciences; Honors Scholar | 1 |
| Bioinformatics | 3 |
| Bioinformatics and Computational Biology | 2 |
| Biological Chemistry | 2 |
| Biological Chemistry; Biological Sciences; Chemistry | 1 |
| Biological Chemistry; Computational and Applied Mathematics; Economics | 1 |
| Biological Chemistry; Philosophy | 1 |
| Biological Engineering | 16 |
| Biological Engineering; Electrical Engineering and Computer Science | 1 |
| Biological Foundations of Behavior: Neuroscience | 1 |
| Biological Science | 7 |
| Biological Science | 1 |
| Biological Sciences | 98 |
| Biological Sciences | 2 |
| Biological Sciences - Health and Human Disease | 1 |
| Biological Sciences (Genetics, Cell, & Dev. Bio.); Neuroscience | 1 |
| Biological Sciences (Genetics, Cell, and Dev Bio) | 1 |
| Biological Sciences (Neurobiology & Behavior) | 1 |
| Biological Sciences (Neurobiology Concentration) | 1 |
| Biological Sciences and Psychology | 1 |
| Biological Sciences- BA | 1 |
| Biological Sciences-Preprofessional | 1 |
| Biological Sciences, Spec. in Cancer Biology | 1 |
| Biological Sciences, Specialization in Immunology | 1 |
| Biological Sciences, Specialization: Endocrinology | 1 |
| Biological Sciences; Criminology and Criminal Justice | 1 |
| Biological Sciences; Global Health Studies | 1 |
| Biological Sciences; Natural and Applied Sciences | 1 |
| Biological Sciences; Natural Sciences | 1 |
| Biological Sciences; Neuroscience | 1 |
| Biological Sciences; Psychology | 2 |
| Biological Sciences; Sociology | 1 |
| Biological Sciences: Cell Biology and Genetics | 1 |
| Biological Sciences: Global and Public Health | 1 |
| Biological Sciences: Human Health and Disease | 1 |
| Biological Sciences: Neuroscience; Psychology | 1 |
| Biological Sciences: Physiology & Neurobiology | 2 |
| Biology | 789 |
| Biology | 1 |
| Biology - biomedical | 1 |
| Biology - Concentration in Human Health/Disease | 1 |
| Biology - Neuroscience | 2 |
| Biology - Physiology, Neuroscience, and Behavior | 1 |
| Biology - Quantitative Biology Track | 1 |
| Biology - Quantitative Track; Neuroscience | 1 |
| Biology (Biochemistry & Biophysics Concentration) | 1 |
| Biology (Biochemistry & Biophysics) | 1 |
| Biology (Biochemistry Concentration) | 1 |
| Biology (Biomedical Sciences concentration); Neuroscience | 1 |
| Biology (Cellular, Molecular, Genetics) | 1 |
| Biology (Computational) | 1 |
| Biology (Concentration in Neurobiology) | 1 |
| Biology (emphasis in Immunobiology) | 1 |
| Biology (General) | 1 |
| Biology (Honors Program) | 1 |
| Biology (Molecular, Cellular, Developmental) | 1 |
| Biology (Neurobiology) | 1 |
| Biology (neuroscience track); Spanish | 1 |
| Biology (Option: Genetics and Genomics) | 1 |
| Biology (Physical Sciences track) | 1 |
| Biology (Physiology) | 1 |
| Biology & Society | 1 |
| Biology and Chemistry | 1 |
| Biology and Psychology | 1 |
| Biology and Society | 3 |
| Biology Emphasis in Human Anatomy and Physiology | 1 |
| Biology for Health Sciences; Chemistry | 2 |
| Biology Medical Sciences | 1 |
| Biology modified with Quantitative Social Sciences | 1 |
| Biology of Global Health | 2 |
| Biology pre-med | 1 |
| Biology Pre-Medical | 1 |
| Biology Sciences | 1 |
| Biology w/ a Specialization in Neuroscience | 1 |
| Biology w/ Concentration in Immunity and Microbes | 1 |
| Biology with a Concentration in Biomedical Science; Biochemistry | 1 |
| Biology with a Concentration in Cell and Molecular | 1 |
| Biology with a concentration in Neuroscience ; Educational Studies | 1 |
| Biology with a specialization in Anatomy | 1 |
| Biology with a Specialization in Cancer Biology | 1 |
| Biology with an emphasis on Biomedical Science | 1 |
| Biology with Cell and Molecular Concentration | 1 |
| Biology with Specialization in Bioinformatics | 1 |
| Biology-Cell and Molecular genetics | 1 |
| Biology-Health Sciences | 2 |
| Biology, Cell Biology and Physiology | 1 |
| Biology, Cellular, Molecular, and Biomedical | 1 |
| Biology, emphases Anatomy and Physiology | 1 |
| Biology, Health, & Society; Movement Science | 1 |
| Biology, Health, and Society | 8 |
| Biology, Health, and Society | 2 |
| Biology, Microbiology and Infectious Diseases | 1 |
| Biology, Molecular & Cell Concentration | 1 |
| Biology, Molecular Cellular & Develop., Honors | 1 |
| Biology, Specialization in Cancer Biology | 1 |
| Biology, Specialization in Global Health Sciences | 1 |
| Biology, Specialization: Physiology & Neurobiology | 1 |
| Biology; Anthropology | 5 |
| Biology; Applied Mathematics | 1 |
| Biology; Applied Mathematics and Statistics | 1 |
| Biology; Biochemistry | 3 |
| Biology; Biochemistry; Neuroscience | 1 |
| Biology; Biology | 1 |
| Biology; Biomedical Engineering | 1 |
| Biology; Biotechnology Science Track Major | 1 |
| Biology; Brain and Cognitive Sciences | 2 |
| Biology; Chemical/Biomolecular Engineering | 1 |
| Biology; Chemistry | 13 |
| Biology; Chemistry- Biochemistry Specialization | 1 |
| Biology; Chemistry; Biochemistry | 1 |
| Biology; Cognitive Science | 1 |
| Biology; Comparative Human Development | 1 |
| Biology; Computer Science | 4 |
| Biology; Computer Science; Neuroscience | 1 |
| Biology; Cultural Anthropology | 1 |
| Biology; Economics | 10 |
| Biology; Education | 1 |
| Biology; Global Health | 6 |
| Biology; Global Health | 1 |
| Biology; Global Health Studies | 1 |
| Biology; History | 1 |
| Biology; Honors Program | 1 |
| Biology; Human Health | 2 |
| Biology; Interdisciplinary - Statistics, Biostatistics | 1 |
| Biology; Mathematics | 2 |
| Biology; Medical Anthropology | 1 |
| Biology; Medical Anthropology & Global Health | 1 |
| Biology; Microbiology / Bacteriology | 2 |
| Biology; Music | 1 |
| Biology; Natural Sciences | 2 |
| Biology; Neuroscience | 4 |
| Biology; Neuroscience and Behavior | 1 |
| Biology; Neuroscience and Behavioral Biology (NBB) | 1 |
| Biology; Nutrition | 1 |
| Biology; Philosophy | 1 |
| Biology; Physics | 1 |
| Biology; Physiology | 1 |
| Biology; Political Science | 2 |
| Biology; Pre-Medical | 2 |
| Biology; Pre-Medical Studies | 1 |
| Biology; Premedical Studies | 1 |
| Biology; Psychological & Brain Sciences | 2 |
| Biology; Psychology | 28 |
| Biology; Psychology - Cognitive Neuroscience | 1 |
| Biology; Public Health | 1 |
| Biology; Statistics | 2 |
| Biology: Biochemistry | 2 |
| Biology: Biochemistry | 1 |
| Biology: Biochemistry and Biophysics | 1 |
| Biology: Biochemisty | 1 |
| Biology: Developmental Genetics Specialization | 1 |
| Biology: Focus in Microbiology | 1 |
| Biology: Genomics and Computational Biology | 1 |
| Biology: Genomics/Computation | 3 |
| Biology: Microbiology | 3 |
| Biology: Microbiology and Biochemistry | 1 |
| Biology: Molecular Biology and Biochemistry | 1 |
| Biology: Molecular Biology and Biochemistry Track | 2 |
| Biology: Neuroscience | 10 |
| Biology: Neuroscience; Anthropology: Global Health and Environment | 1 |
| Biology: Physiology | 1 |
| Biology: Physiology & Neurobiology | 1 |
| Biology: Specialization in cancer biology | 1 |
| Biology:Cell Biology, Molecular Biology & Genetics | 1 |
| Biology:Neuroscience | 1 |
| Biology/Physics Interdisciplinary | 1 |
| Biology/Pre-Med | 2 |
| Biomechanical Engineering | 2 |
| Biomechanics | 1 |
| Biomedical and Health Sciences Engineering | 2 |
| Biomedical Computation | 1 |
| Biomedical Computation, Focus in Organ Systems | 1 |
| Biomedical Discovery and Commercialization | 1 |
| Biomedical Engineering | 189 |
| Biomedical Engineering (Biomechanics) | 1 |
| Biomedical Engineering (Mechanical Emphasis); Philosophy | 1 |
| Biomedical Engineering; Applied Mathematics and Statistics | 2 |
| Biomedical Engineering; Biochemistry | 1 |
| Biomedical Engineering; Biology | 1 |
| Biomedical Engineering; Cell Biology and Neuroscience | 1 |
| Biomedical Engineering; Chemical and Biological Engineering | 1 |
| Biomedical Engineering; Chemical Engineering | 1 |
| Biomedical Engineering; Chemistry | 1 |
| Biomedical Engineering; Cognitive Neuroscience | 1 |
| Biomedical Engineering; Computer Science | 3 |
| Biomedical Engineering; Computer Science; Applied Mathematics and Statistics | 1 |
| Biomedical Engineering; Electrical and Computer Engineering | 1 |
| Biomedical Engineering; Environmental Sociology | 1 |
| Biomedical Engineering; Global Health Studies | 1 |
| Biomedical Engineering; Healthcare Management | 1 |
| Biomedical Engineering; Integrated Science Program | 1 |
| Biomedical Engineering; Mathematics | 2 |
| Biomedical Engineering; Neuroscience | 3 |
| Biomedical Engineering; Plan II Honors | 1 |
| Biomedical Engineering; Public Health | 1 |
| Biomedical Health Sciences | 2 |
| Biomedical physics | 1 |
| Biomedical Physiology Honours | 1 |
| Biomedical Science | 91 |
| Biomedical Science; Biochemistry and Molecular Biology | 1 |
| Biomedical Science; Biotechnology | 1 |
| Biomedical Science; Biotechnology and Molecular Biosciences | 1 |
| Biomedical Science; Chemical Engineering | 1 |
| Biomedical Science; Philosophy | 2 |
| Biomedical Sciences | 13 |
| Biomedical Sciences, Honours | 1 |
| Biometry & Statistics | 1 |
| Biomolecular Science | 4 |
| Biomolecular Sciences | 1 |
| Biophysics | 11 |
| Biophysics, With Highest Honors | 1 |
| Biophysics; Chemistry | 2 |
| Biophysics; Computer Science | 1 |
| Biophysics; Physics (biological sciences concentration) | 1 |
| Biophysics; Public Health | 1 |
| Biopsychology | 9 |
| Biopsychology | 1 |
| Biopsychology, Cognition and Neuroscience | 1 |
| Biopsychology, Cognition, and Neuroscience | 6 |
| Biopsychology, Cognitive, and Neuroscience; Biology, Health, and Society | 1 |
| Biopsychology/neuroscience | 1 |
| Bioscience | 1 |
| Biosciences | 7 |
| Biosciences | 2 |
| Biosciences - Cell Biology and Genetics | 1 |
| Biosciences (Biochemistry Concentration); Neuroscience | 1 |
| Biosciences (concentration in Cell Bio & Genetics) | 1 |
| Biosciences (Concentration: Biochemistry) | 1 |
| Biosciences-Cell Biology & Genetics Concentration | 1 |
| Biosciences, Cell Biology & Genetics; Health Sciences | 1 |
| Biosciences, cell biology and genetics; Psychology | 1 |
| Biosciences: Cell Biology & Genetics | 1 |
| Biosciences: Cell Biology and Genetics | 1 |
| Biostatistics | 4 |
| Biosystems Engineering | 1 |
| Biosystems Engineering; Physiology | 1 |
| Biotechnology | 3 |
| Botany | 1 |
| Brain and Cognitive Sciences | 1 |
| Brain and Cognitive Sciences ; Biology | 1 |
| BS Biological Sciences | 1 |
| Business Economics; Biology | 1 |
| Cancer Biology | 1 |
| Cell & Molecular Biology | 2 |
| Cell and Developmental Biology | 3 |
| Cell and Molecular Biology | 11 |
| Cell Biology | 2 |
| Cell Biology & Neuroscience | 1 |
| Cell Biology & Physiology | 1 |
| Cell Biology and Molecular Genetics | 2 |
| Cell Biology and Molecular Genetics; Psychology | 1 |
| Cell Biology and Neuroscience | 4 |
| Cell Biology and Neuroscience; Honors Program | 1 |
| Cell Biology and Physiology | 5 |
| Cell Biology, Molecular Biology and Genetics | 1 |
| Cell, Molecular, and Developmental Biology | 2 |
| Cellular and Molecular Biology | 1 |
| Cellular and Molecular Biomedical Science | 1 |
| Cellular Biology; Biomedical Physiology | 1 |
| Cellular, Molecular, and Microbial Biology | 1 |
| Celular-Molecular Biology | 1 |
| Chemical & Biomolecular Engineering | 1 |
| Chemical & Biomolecular Engineering; Molecular & Cellular Biology | 1 |
| Chemical & Biomolecular Engineering; Psychology | 1 |
| Chemical and Biological Engineering | 5 |
| Chemical and Biomolecular Engineering | 5 |
| Chemical and Physical Biology | 4 |
| Chemical Biology | 7 |
| Chemical Biology; Immunology & Molecular Medicine | 1 |
| Chemical Biosciences | 2 |
| Chemical Engineering | 38 |
| Chemical Engineering; Biomedical Engineering | 1 |
| Chemical Physics | 2 |
| Chemical Technician; General Studies; Medical Laboratory Sciences | 1 |
| Chemical-Biological Engineering | 1 |
| Chemistry | 144 |
| Chemistry | 1 |
| Chemistry - Specialization in Biochemistry; Global Studies - Global Public Health | 1 |
| Chemistry (Biochemistry Track) | 1 |
| Chemistry (BS): Biochemistry | 1 |
| Chemistry (Emphasis: Biochemistry) | 1 |
| Chemistry (Flex in Biomedical Engineering) | 1 |
| Chemistry (Research) | 1 |
| Chemistry and Biology | 5 |
| Chemistry and Chemical Biology | 1 |
| Chemistry and Physics | 2 |
| Chemistry and Physics; Computer Science | 1 |
| Chemistry Honors | 1 |
| Chemistry Medical Sciences ; Biology Medical Science | 1 |
| Chemistry Pre-Med | 1 |
| Chemistry Pre-Medical Sciences; Biology Pre-Medical Sciences | 1 |
| Chemistry Pre-Medicine | 1 |
| Chemistry with a Biochemistry Concentration | 2 |
| Chemistry with a Concentration in Biochemistry | 1 |
| Chemistry with a specialization in Biochemistry | 1 |
| Chemistry with an Emphasis in Biology | 1 |
| Chemistry with Biochemistry Concentration | 1 |
| Chemistry-ACS, Biochemistry Emphasis | 1 |
| Chemistry, Organic Specialization | 1 |
| Chemistry; Anthropology | 1 |
| Chemistry; Anthropology and Human Biology | 1 |
| Chemistry; Biochemistry | 1 |
| Chemistry; Bioinformatics | 1 |
| Chemistry; Biological Science | 1 |
| Chemistry; Biology | 12 |
| Chemistry; Biology Pre-Health Professions | 1 |
| Chemistry; Biology; Integrated Science | 1 |
| Chemistry; Chemical Engineering | 1 |
| Chemistry; Economics | 1 |
| Chemistry; Emphasis in Biochemistry | 1 |
| Chemistry; Honors Program | 1 |
| Chemistry; Human Physiology | 1 |
| Chemistry; Microbiology / Bacteriology | 1 |
| Chemistry; Molecular Biology | 1 |
| Chemistry; Molecular Biology and Biochemistry | 1 |
| Chemistry; Neurobiology; Biochemistry | 1 |
| Chemistry; Neuroscience | 1 |
| Chemistry; Physics | 1 |
| Chemistry; Psychology | 2 |
| Chemistry; Public Health | 1 |
| Chemistry; Thomas Hunter Special Honors Program Major | 1 |
| Chemistry: Biochemistry Concentration | 2 |
| Chemistry/Biochemistry Emphasis | 1 |
| Chemistry/Biochemistry; Music | 1 |
| Chemistry/Materials Science | 1 |
| Civil Engineering | 2 |
| Clinical & Applied Health Sciences | 1 |
| Clinical Microbiology | 1 |
| Clinical Neuroscience | 2 |
| Clinical Psychology | 1 |
| Clinical Psychology; Human Biology | 1 |
| Cognitive and Behavioral Neuroscience | 1 |
| Cognitive Brain Science | 1 |
| Cognitive Neuroscience | 11 |
| Cognitive Neuroscience and Evolutionary Psychology | 1 |
| Cognitive Science | 9 |
| Cognitive Science; Computer Science; Biopsychology, Cognition, and Neuroscience | 1 |
| Cognitive Science; Psychology | 1 |
| Cognitive Sciences | 4 |
| Cognitive Systems | 1 |
| Cognitive- Neuroscience | 1 |
| Combined Major in Science | 1 |
| Community and Global Public Health; Neuroscience | 1 |
| Computation and Cognition | 2 |
| Computation and Neural Systems | 1 |
| Computational and Systems Biology | 5 |
| Computational Biology | 8 |
| Computational Biology | 1 |
| Computational Biology & Genomics | 1 |
| Computational Biology, Applied Math Track | 1 |
| Computational Neuroscience | 1 |
| Computer Engineering | 2 |
| Computer Science | 36 |
| Computer Science (Artificial Intelligence) | 1 |
| Computer Science & Biology | 1 |
| Computer Science + Math | 1 |
| Computer Science and Biology | 1 |
| Computer Science and Math | 1 |
| Computer Science and Mathematics | 1 |
| Computer Science and Mathematics; Biochemistry and Molecular Biology | 1 |
| Computer Science and Molecular Biology | 2 |
| Computer Science and Molecular Biology | 1 |
| Computer Science B S Comp Sc; University Honors Program Major | 1 |
| Computer Science, Data Science Track | 1 |
| Computer Science; Applied Mathematics - Economics | 1 |
| Computer Science; Applied Mathematics & Statistics | 1 |
| Computer Science; Biology | 4 |
| Computer Science; Chemistry | 1 |
| Computer Science; Cognitive Science | 1 |
| Computer Science; Economics | 2 |
| Computer Science; Neuroscience | 2 |
| Computer Science; Neuroscience; Chemistry | 1 |
| Cultural Anthropology | 1 |
| Dairy Science | 1 |
| Data Science | 1 |
| Developmental and Regenerative Biology | 1 |
| Developmental Biology/Genetics | 1 |
| Diagnostic Medical Ultrasound Technology | 1 |
| Earth & Environmental Science; Biology | 1 |
| Ecology & Evolutionary Biology | 3 |
| Ecology & Evolutionary Biology; Neuroscience | 1 |
| Ecology and Evolutionary Biology | 4 |
| Economics | 34 |
| Economics (Honors) | 1 |
| Economics (Honors); Biochemistry | 1 |
| Economics with Specialization in Data Science | 1 |
| Economics, Specialization in Business Economics; Biological Sciences | 1 |
| Economics; Biological Sciences | 1 |
| Economics; Biology | 2 |
| Economics; Biology modified with Mathematics | 1 |
| Economics; Biology; Zoology | 1 |
| Economics; Biomolecular Science | 1 |
| Economics; Chemistry | 1 |
| Economics; Molecular and Cell Biology | 1 |
| Economics; Neuroscience | 1 |
| Economics; Pharmaceutical Chemistry | 1 |
| Economics; Pre-Health | 1 |
| Electrical and Computer Engineering | 2 |
| Electrical and Computer Engineering; Computer Science | 1 |
| Electrical Engineering | 6 |
| Electrical Engineering; Systems Science: Systems Engineering | 1 |
| Engineering as Recommended by Dept. of Mech. Engr. | 1 |
| Engineering Physics | 1 |
| Engineering Physics and Biomedical Engineering | 1 |
| Engineering Science | 1 |
| Engineering Sciences mod/ Neuroscience | 1 |
| Engineering Sciences- Materials Science; Chemistry | 1 |
| Engineering Sciences, Chemical | 1 |
| Engineering, Biomedical | 1 |
| English Literary Studies; Molecular and Cellular Biology | 1 |
| English; Biological Sciences | 1 |
| Environmental Biology | 2 |
| Environmental Engineering | 2 |
| Environmental Science | 3 |
| Environmental Studies | 3 |
| Evolution, Ecology, and Biodiversity | 1 |
| Evolutionary (Biological) Anthropology | 1 |
| Evolutionary Anthropology | 3 |
| Exercise & Sports Science | 1 |
| Exercise and Health Physiology | 1 |
| Exercise Biology | 1 |
| Exercise Science | 7 |
| Exercise Science | 1 |
| Exercise Science and Pre-Health Professions | 2 |
| Field Biology | 1 |
| Food systems, nutrition, and health | 1 |
| General Biology | 3 |
| General Science | 1 |
| General Studies Chemistry | 1 |
| Genetics | 12 |
| Genetics & Genomics | 1 |
| Genetics and Genomics Sciences | 1 |
| Genetics, Cell Biology and Development | 1 |
| Genetics, Cell Biology, and Development | 1 |
| Genetics, Cellular Biology, and Development; Physics; Computer Science | 1 |
| Genetics; Computer Science | 1 |
| Genetics/Cell Biology/Development | 1 |
| Genomics & Molecular Genetics; Neuroscience | 1 |
| Genomics and Computational Biology | 1 |
| Global Disease Biology | 2 |
| Global Health Narrative (Independent Major); Biomedical Engineering | 1 |
| Global Health Studies ; Psychology | 1 |
| Global Health; Biology | 1 |
| Global Health; Cultural Anthropology | 1 |
| Health & Human Physiology | 2 |
| Health and Disease; Psychology | 1 |
| Health and Exercise Science | 2 |
| Health and Human Biology | 2 |
| Health and Human Physiology, Health Studies Track | 1 |
| Health and Human Sciences | 4 |
| Health and Human Sciences (BA); Healthcare Decision Analysis (MS) | 1 |
| Health and Kinesiology | 1 |
| Health and Societies ; Conc. in Public Health | 1 |
| Health and the Human Sciences | 1 |
| Health Promotion & Disease Prevention | 1 |
| Health Science | 13 |
| Health Science | 1 |
| Health Science- Emphasis in Pre-Professional | 1 |
| Health Sciences | 19 |
| Health Sciences | 1 |
| Health Sciences (Honors) | 1 |
| Health Sciences (Pre-Clinical Track) | 1 |
| Health Studies - Science Emphasis | 1 |
| Health Studies: Health Science; Science; University Honors College | 1 |
| Highest Honors in Biomolecular Science | 1 |
| Highest Honors in Integrative Biology | 1 |
| History and Philosophy of Science and Medicine | 1 |
| History and Science | 1 |
| History of Science, Medicine, and Public Health | 1 |
| History of Science, Medicine, and Public Health; Molecular, Cellular, and Developmental Biology | 1 |
| Honors Anatomy and Cell Biology | 1 |
| Honors Biochemistry and Molecular Biology | 1 |
| Honors Chemistry | 1 |
| Honors in Molecular & Cell Biology | 1 |
| Honors Program; Biomedical Engineering | 1 |
| Honors Science | 1 |
| Honors Specialization in Biology | 1 |
| Honors Specialization in Health Sciences | 1 |
| Honors Specialization in Neuroscience | 1 |
| Honors Specialization in Physiology + Pharmacology | 1 |
| Honour Specialization in Neuroscience | 1 |
| Honours Anatomy & Cell Biology | 1 |
| Honours Biochemistry | 1 |
| Honours in Biology | 1 |
| Honours Interdisciplinary Health Sciences | 1 |
| Honours Life Sciences | 1 |
| Honours Neuroscience | 1 |
| Honours Spec. in Physiology and Pharmacology | 1 |
| Honours Translational and Molecular Medicine | 1 |
| Human Science | 1 |
| Human Biology | 91 |
| Human Biology | 1 |
| Human Biology & Society | 1 |
| Human Biology and Society | 11 |
| Human Biology and Society with Honors; Ecology, Behavior, and Evolution with Honors | 1 |
| Human Biology and Society- BS | 1 |
| Human Biology Health and Society | 1 |
| Human Biology, Health & Disease Specialist; Cell & Systems Biology Major | 1 |
| Human Biology, Health, and Society | 12 |
| Human Biology; Anthropology | 1 |
| Human Biology; Clinical Psychology | 1 |
| Human Biology; Economics | 3 |
| Human Biology; Foreign Language - Spanish | 1 |
| Human Biology; Genetics | 1 |
| Human Biology; Political Science; Economics | 1 |
| Human Biology; Psychology | 1 |
| Human Biology: Health & Disease; Immunology | 1 |
| Human Biology‚ÄîNeuroscience and Neurodegeneration | 1 |
| Human Development and Family Science's | 1 |
| Human Developmental & Regenerative Biology | 1 |
| Human Developmental and Regenerative Biology | 7 |
| Human Evolutionary Biology | 6 |
| Human Evolutionary Biology ; Medical Anthropology/ Global Health | 1 |
| Human Physiology | 7 |
| Human Physiology, B.S. | 1 |
| Human Science | 1 |
| Immunology | 2 |
| Industrial Engineering | 1 |
| Infectious Disease and Economic Development | 1 |
| Information Science | 2 |
| Integrated Biology | 1 |
| Integrated Sciences | 1 |
| Integrative Biology | 12 |
| Integrative Biology (Integrative Human Biology) | 2 |
| Integrative Human Biology | 2 |
| Integrative Neuroscience | 3 |
| Integrative Neuroscience - Cell and Molecular | 1 |
| Integrative Physiology | 5 |
| Integrative Physiology; Neuroscience; Psychology | 1 |
| Intensive Molecular, Cellular, and Dev. Biology | 1 |
| Interdepartmental Honours Immunology | 1 |
| Interdepartmental Studies - Health Science | 1 |
| Interdisciplinary - Translational Medicine | 1 |
| Interdisciplinary Health Sciences | 2 |
| Interdisciplinary Medical Sciences | 2 |
| Interdisciplinary Medical Sciences; Pharmacology | 1 |
| International Economics | 1 |
| Investigative and Medical Sciences | 2 |
| Joint Chemistry/Biology | 1 |
| Kinesiology | 16 |
| Kinesiology - Exercise & Movement Science | 1 |
| Kinesiology (Exercise Science) | 1 |
| Kinesiology & Health | 1 |
| Kinesiology and Health Sciences | 1 |
| Kinesiology BS | 1 |
| Kinesiology Pre-Therapy and Allied Health | 1 |
| Kinesiology- Movement Science | 1 |
| Kinesiology: Pre-Physical Therapy | 1 |
| Laboratory Medicine and Pathobiology | 2 |
| Laboratory Medicine and Pathobiology | 1 |
| Life Science | 3 |
| Life Science Specialization | 1 |
| Life Sciences | 5 |
| Life Sciences (Honours) | 1 |
| Life Sciences: Origins of Disease | 1 |
| Linguistics | 6 |
| Linguistics; Computer Science | 1 |
| Linguistics; Neuroscience | 1 |
| Major: AAAS; Concentration: Biological Sciences | 1 |
| Marine Biology | 1 |
| Materials Science and Engineering | 5 |
| Mathematical and Computational Science | 1 |
| Mathematical Sciences | 1 |
| Mathematics | 22 |
| Mathematics; Biochemistry | 1 |
| Mathematics; Biology | 3 |
| Mathematics; Chemistry | 2 |
| Mathematics; Computer Science | 1 |
| Mathematics; Neuroscience | 1 |
| Mechanical and Aerospace Engineering | 1 |
| Mechanical Engineering | 25 |
| Mechanical Engineering; Physics | 1 |
| Medical Anthropology | 2 |
| Medical Diagnostics; Public Policy | 1 |
| Medical Laboratory Science | 1 |
| Medical Laboratory Sciences | 1 |
| Medical Sciences | 11 |
| Medical Studies | 1 |
| Medicine, Health, and Society | 4 |
| Medicine, Health, and Society | 1 |
| Medicine, Science, and the Humanities | 1 |
| Microbial Biology | 2 |
| Microbiology | 14 |
| Microbiology | 7 |
| Microbiology - Biotechnology | 1 |
| Microbiology / Bacteriology | 17 |
| Microbiology / Bacteriology; Biochemistry | 2 |
| Microbiology / Bacteriology; Philosophy | 1 |
| Microbiology and Cell and Molecular Biology; Physiology | 1 |
| Microbiology and Cell Science | 2 |
| Microbiology and Cell Science; Anthropology | 1 |
| Microbiology and Cell Science; Economics | 1 |
| Microbiology and Cell Sciences | 1 |
| Microbiology Immunology Molecular Genetics | 1 |
| Microbiology with Honors; Public Health-Global Health with Honors | 1 |
| Microbiology, Immunology and Molecular Genetics | 2 |
| Microbiology, Immunology, and Molecular Genetics | 11 |
| Microbiology, Immunology, and Molecular Genetics; Economics | 1 |
| Microbiology, Immunology, Molecular Genetics | 1 |
| Microbiology; Anthropology: Global Health and Environment | 1 |
| Microbiology; Genomics & Molecular Genetics | 1 |
| Microbiology; Psychology | 1 |
| Microbiology/Immunology; Public Health | 1 |
| Molecular & Cell Bio: Biochem, Biophys & Struc Bio | 1 |
| Molecular & Cell Biology | 7 |
| Molecular & Cell Biology - Immunology | 1 |
| Molecular & Cell Biology - Neurobiology; Public Health | 1 |
| Molecular & Cell Biology; Economics | 1 |
| Molecular & Cell Biology; Integrative Biology | 1 |
| Molecular & Cell Biology; Mathematics | 1 |
| Molecular & Cell Biology; Psychology | 1 |
| Molecular & Cell Biology; Public Health | 1 |
| Molecular & Cellular Biology | 6 |
| Molecular & Cellular Biology - Neurobiology; Public Health | 1 |
| Molecular & Cellular Biology Honors Concentration; Chemistry | 1 |
| Molecular & Cellular Biology; Behavioral Biology | 1 |
| Molecular & Cellular Biology; Economics | 1 |
| Molecular & Cellular Biology; Mathematics | 1 |
| Molecular & Cellular Biology; Medicine, Health, and Society | 1 |
| Molecular & Cellular Biology; Public Health | 1 |
| Molecular & Cellular Biology; Public Health Studies | 2 |
| Molecular and Cell Biology | 43 |
| Molecular and Cell Biology - Infectious Disease; Public Health | 1 |
| Molecular and Cell Biology - Neurobiology | 1 |
| Molecular and Cell Biology (Neurobiology emphasis) | 1 |
| Molecular and Cell Biology w/ Honors | 1 |
| Molecular and Cell Biology, Immunology | 1 |
| Molecular and Cell Biology, Immunology Emphasis; Public Health, Health Policy Capstone | 1 |
| Molecular and Cell Biology, Immunology; Integrative Human Biology | 1 |
| Molecular and Cell Biology; Bioengineering | 1 |
| Molecular and Cell Biology; Data Science | 1 |
| Molecular and Cell Biology; Nutritional Sciences: Toxicology | 1 |
| Molecular and Cell Biology; Psychology | 1 |
| Molecular and Cell Biology; Public Health | 1 |
| Molecular and Cell Biology: Genetics | 1 |
| Molecular and Cell Biology: Neurobiology | 1 |
| Molecular and Cellular Biology | 57 |
| Molecular and Cellular Biology | 1 |
| Molecular and Cellular Biology - Neurobiology | 1 |
| Molecular and Cellular Biology ; Public Health Studies | 1 |
| Molecular and Cellular Biology, Honors Conc. | 1 |
| Molecular and Cellular Biology; Behavioral Neuroscience | 1 |
| Molecular and Cellular Biology; Chemistry | 1 |
| Molecular and Cellular Biology; Chemistry; Neuroscience | 1 |
| Molecular and Cellular Biology; Medicine Health and Society- Global Health | 1 |
| Molecular and Cellular Biology; Medicine, Health, and Society | 1 |
| Molecular and Cellular Biology; Psychology | 3 |
| Molecular and Cellular Biology; Public Health | 2 |
| Molecular and Medical Microbiology | 1 |
| Molecular Biochemistry and Biophysics | 1 |
| Molecular Biology | 69 |
| Molecular Biology - biochemistry | 1 |
| Molecular Biology (emphasis in Neurobiology) | 1 |
| Molecular Biology & Cellular Biology | 1 |
| Molecular Biology and Biochemistry | 5 |
| Molecular Biology and Biochemistry | 1 |
| Molecular Biology and Biochemistry (joint degree) | 1 |
| Molecular Biology and Biochemistry; ANTHROPOLOGY: GLOBAL HEALTH AND ENVIRONMENT | 1 |
| Molecular Biology; Biotechnology | 1 |
| Molecular Biology; Data Science | 1 |
| Molecular Biology; History and Philosophy of Science | 1 |
| Molecular Biology; Psychology | 1 |
| Molecular Biology; Public Health | 2 |
| Molecular Biology; Statistics | 1 |
| Molecular Biophysics & Biochemistry | 2 |
| Molecular Biophysics & Biochemistry | 1 |
| Molecular Biophysics and Biochemistry | 7 |
| Molecular Biophysics and Biochemistry; Classics | 1 |
| Molecular Biophysics and Biochemistry; History of Science, Medicine, and Public Health | 1 |
| Molecular Cell and Developmental Biology | 1 |
| Molecular Cell Biology | 1 |
| Molecular Cell Biology and Physiology | 1 |
| Molecular Cell Biology-Biochemistry | 1 |
| Molecular Cell Biology-Neurobiology; Psychology | 1 |
| Molecular Cellular & Developmental Biology | 1 |
| Molecular Cellular and Development Biology | 1 |
| Molecular Cellular and Developmental Biology | 1 |
| Molecular Cellular Biology | 1 |
| Molecular Cellular Biology, Neurology | 1 |
| Molecular Engineering; Biological Sciences | 1 |
| Molecular Environmental Biology | 4 |
| Molecular Genetics | 2 |
| Molecular Life Sciences; Music (Piano) | 1 |
| Molecular Microbiology and Immunology | 1 |
| Molecular Neuroscience | 1 |
| Molecular, Cell and Developmental Biology | 5 |
| Molecular, Cell and Developmental Biology; Psychobiology | 1 |
| Molecular, Cell, and Developmental Biology | 27 |
| Molecular, Cell, and Developmental Biology (MCDB); Anthropology | 1 |
| Molecular, Cell, and Developmental Biology; Neuroscience | 1 |
| Molecular, Cellular and Developmental Biology | 6 |
| Molecular, Cellular and Developmental Biology | 1 |
| Molecular, Cellular, & Developmental Biology | 2 |
| Molecular, Cellular, & Developmental Biology; Biochemistry | 1 |
| Molecular, Cellular, and Dev. Biology (Intensive) | 1 |
| Molecular, Cellular, and Development Biology | 1 |
| Molecular, Cellular, and Developmental Biology | 45 |
| Molecular, Cellular, and Developmental Biology | 2 |
| Molecular, Cellular, and Developmental Biology; Biochemistry | 1 |
| Molecular, Cellular, and Developmental Biology; Biopsychology, Cognition, and Neuroscience | 1 |
| Molecular, Cellular, and Developmental Biology; Ecology and Evolutionary Biology | 1 |
| Molecular, Cellular, and Developmental Biology; History | 1 |
| Molecular, Cellular, and Developmental Biology; Spanish | 1 |
| Molecular, Cellular, Developmental Biology | 3 |
| Molecular, Cellular, Developmental Biology; Applied Behavioral Science - ECE&I | 1 |
| Molecular, Cellular, Organismal Biology | 1 |
| Molecular/ Cellular Neuroscience; Biochemistry | 1 |
| Movement Science | 4 |
| Movement Science | 1 |
| Music (Ballet) and an Outside Field (Biology) | 1 |
| Natural Science | 1 |
| Natural Sciences | 2 |
| Natural Sciences-Health Sciences | 1 |
| Natural Sciences; Psychology | 1 |
| Neural Science | 4 |
| Neurobiology | 18 |
| Neurobiology - Molecular and Cell Biology | 1 |
| Neurobiology and Music Performance | 1 |
| Neurobiology and Physiology | 1 |
| Neurobiology and Physiology ; Psychology | 1 |
| Neurobiology, Physiology and Behavior | 3 |
| Neurobiology, Physiology and Behavior | 2 |
| Neurobiology, Physiology and Behavior; Psychology | 1 |
| Neurobiology, Physiology, & Behavior | 1 |
| Neurobiology, Physiology, and Behavior | 8 |
| Neurobiology, Physiology, and Behavior; Psychology | 1 |
| Neurobiology; Biochemistry | 1 |
| Neurobiology; Biophysics | 1 |
| Neurobiology; Global Health | 1 |
| Neurobiology; Nutritional Science | 1 |
| Neuroscience | 457 |
| Neuroscience | 1 |
| Neuroscience - B.S. | 1 |
| Neuroscience - Biology Track | 1 |
| Neuroscience (Undergraduate); Bioinformatics (Master's) | 1 |
| Neuroscience [BS]; Global Health [BA] | 1 |
| Neuroscience & Behavior; Economics | 1 |
| Neuroscience & Behavioral Biology | 4 |
| Neuroscience and Behavior | 24 |
| Neuroscience and Behavior | 2 |
| Neuroscience and Behavioral Biology | 11 |
| Neuroscience and Behavioral Biology | 1 |
| Neuroscience and Behavioral Biology; Anthropology and Human Biology | 1 |
| Neuroscience and Behavioral Biology; Chemistry | 1 |
| Neuroscience and Behavioral Biology; Economics | 1 |
| Neuroscience and Behavioral Biology; French | 1 |
| Neuroscience and Cognitive Science | 1 |
| Neuroscience Bachelor's, Bioengineering Master's | 1 |
| Neuroscience Honors; University Honors | 1 |
| Neuroscience Scholars | 1 |
| Neuroscience with Highest Honors | 1 |
| Neuroscience, Cellular & Molecular; Neuroscience, Cognitive | 1 |
| Neuroscience, Cognitive/Behavioral Concentration; Psychology | 1 |
| Neuroscience, Honors; Psychology | 1 |
| Neuroscience; Anthropology | 1 |
| Neuroscience; Applied Mathematics (Data Science & Statistics) | 1 |
| Neuroscience; Applied Mathematics and Statistics | 1 |
| Neuroscience; Biochemistry | 2 |
| Neuroscience; Biology | 8 |
| Neuroscience; Cell and Molecular Biology | 1 |
| Neuroscience; Chemistry | 2 |
| Neuroscience; Child Development | 1 |
| Neuroscience; Cognitive Science | 2 |
| Neuroscience; Computational and Systems Biology | 1 |
| Neuroscience; Computer Science | 2 |
| Neuroscience; Economics | 6 |
| Neuroscience; Genetics, Cell Biology, and Development | 1 |
| Neuroscience; Global Health Studies | 2 |
| Neuroscience; Human Biology | 2 |
| Neuroscience; Human Biology and Society | 1 |
| Neuroscience; Human Physiology | 1 |
| Neuroscience; Information Science | 1 |
| Neuroscience; Mathematics | 1 |
| Neuroscience; Mathematics and Computer Science | 1 |
| Neuroscience; Medical Anthropology | 1 |
| Neuroscience; Medicine | 1 |
| Neuroscience; Medicine Health and Society | 1 |
| Neuroscience; Molecular and Cellular Biology | 1 |
| Neuroscience; Molecular Life Sciences (Developmental & Cellular) | 1 |
| Neuroscience; Molecular/Cellular Biology | 1 |
| Neuroscience; Pathobiology | 1 |
| Neuroscience; Philosophy | 2 |
| Neuroscience; Physics | 1 |
| Neuroscience; Plan II Honors | 1 |
| Neuroscience; Plan II Honors Program | 2 |
| Neuroscience; Psychology | 13 |
| Neuroscience; Public Health | 3 |
| Neuroscience; Sociology | 3 |
| Neuroscience; Statistics and Data Science | 1 |
| Neuroscience: Cell & Molecular | 1 |
| No Major; Anthropology | 1 |
| No Major; Economics | 1 |
| Nuclear Engineering | 1 |
| Nursing combined with Public Health | 1 |
| Nutrition | 6 |
| Nutrition and Food Science | 1 |
| Nutrition and Food Science: Dietetics | 1 |
| Nutrition Science | 1 |
| Nutrition Science | 1 |
| Nutrition Science and Research | 1 |
| Nutrition Sciences | 1 |
| Nutrition; Biochemistry | 1 |
| Nutrition; Economics | 1 |
| Nutrition; Human Biology | 1 |
| Nutritional Biochemistry and Metabolism | 1 |
| Nutritional Science | 1 |
| Nutritional Science - Physiology and Metabolism | 1 |
| Nutritional Sciences | 3 |
| Nutritional Sciences and Toxicology | 1 |
| Nutritional Sciences; Microbiology and Cell Science | 1 |
| Nutritional Sciences: Physiology and Metabolism | 1 |
| Operations Research and Financial Engineering | 1 |
| Paramedicine | 1 |
| Pathology | 1 |
| Pharmaceutical Science | 1 |
| Pharmaceutical Sciences | 7 |
| Pharmacology | 6 |
| Pharmacology & Toxicology; Biology | 1 |
| Pharmacology; Interdisciplinary Medical Science | 1 |
| Pharmacology; Interdisciplinary Medical Sciences | 1 |
| Pharmacology; Physiology | 1 |
| PHIL-NEURO-PSYCH: COGNITIVE NEUROSCIENCE | 1 |
| Philosophy-Neuroscience-Psychology | 6 |
| Philosophy-Neuroscience-Psychology: Cog Neuro | 1 |
| Philosophy-Neuroscience-Psychology: Cognitive Neur | 1 |
| Philosophy, Neuroscience, and Psychology | 1 |
| Philosophy, Politics, and Economics | 1 |
| Philosophy, Politics, Economics and Law (PPEL) | 1 |
| Philosophy; Biochemistry | 1 |
| Philosophy; Biology | 2 |
| Philosophy; Neurobiology, Honors in the Major | 1 |
| Physics | 16 |
| Physics; Brain & Cognitive Science | 1 |
| Physics; Computer Science | 1 |
| Physics; Mathematics | 1 |
| Physics; Neuroscience | 1 |
| Physics; Psychology | 1 |
| Physiological & Organismal Biology | 1 |
| Physiological Science | 19 |
| Physiological Science | 1 |
| Physiological Sciences | 5 |
| Physiology | 21 |
| Physiology & Neurobiology | 2 |
| Physiology & Pharmacology | 1 |
| Physiology and Developmental Biology | 2 |
| Physiology and Developmental Biology | 1 |
| Physiology and Medical Sciences | 1 |
| Physiology and Medical Sciences; Mathematics | 1 |
| Physiology and Neurobiology | 2 |
| Physiology and Neurobiology | 1 |
| Physiology and Neurobiology; Economics | 1 |
| Physiology and Neuroscience | 3 |
| Physiology specialist | 1 |
| Physiology Specialist; Cell & Molecular Biology Major | 1 |
| Physiology; Cellular, Physiological and Pharmacological Scienc | 1 |
| Physiology; Human Biology | 2 |
| Physiology; Interdisciplinary Medical Sciences | 1 |
| Physiology; Interdisciplinary Medical Sciences (IMS) | 1 |
| Physiology; Pathology | 1 |
| PNP (Philosophy, Neuroscience, Psychology) | 1 |
| Pre-Medical | 13 |
| Pre-Medical - Biology | 1 |
| Pre-Medical Biology | 1 |
| Pre-Medical; Biochemistry | 2 |
| Program II: Medicine, Music, and Mind | 1 |
| Psych & Brain Sci: Cognitive Neuroscience | 1 |
| Psych and Brain Sciences: Cognitive Neuroscience | 1 |
| Psych. (Cog. Neuroscience & Evolutionary Psych.); Human Evolutionary Biology | 1 |
| Psychobiology | 25 |
| Psychobiology; Economics | 1 |
| Psychological & Brain Sciences | 1 |
| Psychological and Brain Sciences | 1 |
| Psychology | 109 |
| Psychology (Concentration in Neuroscience) | 1 |
| Psychology & Brain Sciences:Cognitive Neuroscience | 1 |
| Psychology and Brain Sciences | 1 |
| Psychology with Biological Emphasis; Pharmaceutical Chemistry | 1 |
| Psychology with Neuroscience Area of Emphasis | 1 |
| Psychology, Neuroscience & Behaviour | 1 |
| Psychology; Anthropology | 1 |
| Psychology; Biochemistry & Molecular Biology | 1 |
| Psychology; Biology | 3 |
| Psychology; Cell Biology and Neuroscience | 1 |
| Psychology; Chemistry | 1 |
| Psychology; Child Studies and Human Development | 1 |
| Psychology; Cognitive Science | 1 |
| Psychology; Global Health | 2 |
| Psychology; Global Health | 1 |
| Psychology; Health & Nutrition Sciences | 1 |
| Psychology; Human Biology | 1 |
| Psychology; Human Development & Family Science | 1 |
| Psychology; Molecular & Cell Biology | 1 |
| Psychology; Molecular and Cell Biology - Neurobiology | 1 |
| Psychology; Molecular Biology | 1 |
| Psychology; Natural Sciences | 1 |
| Psychology; Neuroscience | 3 |
| Psychology; Neuroscience-Biology | 1 |
| Psychology; Pre-Health | 1 |
| Psychology; Pre-Medical | 1 |
| Psychology; Public Health | 1 |
| Public Health | 30 |
| Public Health Science | 1 |
| Public Health Sciences | 1 |
| Public Health Studies | 3 |
| Public Health w/ Cell Science Emphasis | 1 |
| Public Health w/ Conc in Medicine Sciences; Molecular and Cell Biology (with Honors) | 1 |
| Public Health; Biochemistry | 1 |
| Public Health; Biology | 2 |
| Public Health; Health Behavior | 1 |
| Public Health; History of Science, Medicine, and Technology | 1 |
| Public Health; Microbiology / Bacteriology | 1 |
| Public Health; Molecular & Cell Biology emphasis in Neurobiology | 1 |
| Public Health; Molecular and Cell Biology | 2 |
| Public Health; Molecular and Cellular Biology | 1 |
| Public Health; Molecular Biology | 1 |
| Public Health; Natural Sciences | 2 |
| Public Health; Neuroscience | 2 |
| Public Health; Plan II Honors | 1 |
| Public Health; Plan II Honors | 1 |
| Quantitative Biology | 2 |
| Quantitative Biosciences and Engineering | 1 |
| Quantitative Science Concentration in Neuroscience | 1 |
| Quantitative Social Science | 1 |
| Quantitative Social Science (Public Policy focus) | 1 |
| Quantitative/Data Science with a Biology Focus | 1 |
| Science (Other Biology) | 6 |
| Science (Other Biology), Health Sciences | 1 |
| Science (Other Biology); Neuroscience | 1 |
| Science and Technology Studies | 1 |
| Science General | 2 |
| Science Pre-Professional Studies | 1 |
| Science Research Fellows | 2 |
| Science-Business | 1 |
| Science-Computing | 1 |
| Shaped Study in Public Health; Biochemistry | 1 |
| Social Anthropology; Neuroscience | 1 |
| Sociology | 14 |
| Sociology (Intensive) | 1 |
| Sociology; Biology | 2 |
| Sociology; Molecular Biology | 1 |
| Sociology; Psychology | 1 |
| Sociology/Anthropology; Biology | 1 |
| Sp Maj: Neuroscience | 1 |
| Special Major in Medical Anthropology | 1 |
| Sport and Exercise Science | 1 |
| Sports Medicine | 1 |
| Sports Medicine | 1 |
| Sports Medicine & Exercise Physiology | 2 |
| Sports Medicine and Exercise Physiology | 5 |
| Statistical Science | 1 |
| Statistics | 5 |
| Statistics and Molecular and Cellular Biology | 1 |
| Statistics; Chemistry | 1 |
| Statistics; Human Biology | 1 |
| Veterinary Medicine | 1 |
| Zoology | 1 |
| Neuroscience; Molecular, Cell & Developmental Biology | 1 |
| Program Honors in Biological Sciences | 1 |

| **Non-STEM majors** | **Number reporting this major** |
| --- | --- |
| Accounting | 1 |
| Architecture | 1 |
| Athletic Training | 1 |
| Biology, Health, and Society; Gender and Health | 1 |
| Business | 3 |
| Business Administration | 3 |
| Business Administration, Finance | 1 |
| Business Analytics | 1 |
| Business Fellows | 1 |
| Business Fellows; Finance | 1 |
| Business Fellows; Management | 1 |
| Business management | 1 |
| Business Management w/ conc. in Healthcare | 1 |
| Business, Concentration in Finance | 1 |
| Business; Medicine | 1 |
| Cello Performance | 1 |
| Classical Civilizations | 1 |
| Classics | 3 |
| Classics; History | 1 |
| Clinical Professions | 1 |
| Communications | 1 |
| Community Health | 1 |
| Comparative Literature | 2 |
| Comparative Literature (French concentration) | 1 |
| Creative Writing | 2 |
| Criminal Justice | 1 |
| Criminology | 1 |
| Criminology/Criminal Justice | 2 |
| Dance - Pedagogy | 1 |
| Department of History | 1 |
| Dietetics | 1 |
| East Asian Studies and Government | 1 |
| Education | 1 |
| Emergency Administration and Management | 1 |
| Emergency and Disaster Management | 1 |
| Emergency Medical Services Management; Pre-Medical | 1 |
| English | 18 |
| English; Mass Communications; (12 months graduate coursework) Product Innovation | 1 |
| Evolution of Consciousness | 1 |
| Fibers | 1 |
| Film Production | 1 |
| Finance | 7 |
| Finance, Investment, and Banking ; Real Estate and Urban Land Economics | 1 |
| Finance; Accounting | 1 |
| Finance; Economics | 1 |
| Foreign Language | 4 |
| Foreign Language - Spanish | 1 |
| Forensic Science | 1 |
| Francophone Studies | 1 |
| Gender & Sexuality Studies; Social Policy | 1 |
| Gender Studies | 1 |
| Geography | 3 |
| Global and Community Health | 1 |
| Global Cultural Studies | 1 |
| Global Health | 4 |
| Global Studies, conc. in Health & Environment | 1 |
| Government and Legal Studies, Comparative Politics | 1 |
| Health and Society | 3 |
| Health Care Management and Policy | 1 |
| Health Care Policy | 1 |
| Health Education and Behavior | 1 |
| Health Equity | 1 |
| Health Management | 1 |
| Health Management & Policy | 1 |
| Health Promotion and Disease Prevention | 1 |
| Health Promotion and Disease Prevention Studies | 1 |
| Health Society and Policy | 1 |
| Health, Society and Policy | 1 |
| Healthcare Administration | 1 |
| Healthcare Management | 1 |
| Healthcare Management & Policy | 1 |
| Healthcare Management; Organization and Strategic Management | 1 |
| Hispanic Studies | 1 |
| History | 12 |
| History & Literature | 1 |
| History and Science | 2 |
| History and Science, cum laude | 1 |
| History of Science | 1 |
| History of Science, Medicine, and Public Health; German Studies | 1 |
| History; International Relations | 1 |
| Honors Co-Major; German Honors | 1 |
| Honors Program; Nutrition; International Board Certified Lactation Consultant | 1 |
| Honors Specialization in One Health | 1 |
| Human and Organizational Development; Medicine, Health, and Society | 1 |
| Human Development | 2 |
| Human Development and Family Studies | 1 |
| Humanities | 1 |
| Industrial and Labor Relations | 1 |
| Interdisciplinary Humanities Studies | 1 |
| Interdisciplinary Project in the Humanities | 1 |
| International Business | 1 |
| International Relations | 1 |
| International Studies | 2 |
| International Studies (Political Science) | 1 |
| Japanese | 1 |
| Jazz Piano Performance | 1 |
| Jewish Studies; French; Religious Studies | 1 |
| Journalism | 2 |
| Liberal Arts Music | 1 |
| Literature and Culture Studies | 1 |
| Magnetic Resonance Imaging | 1 |
| Management | 1 |
| Management; Clifton Builders | 1 |
| Marketing/Business | 1 |
| Medical Humanities | 1 |
| Medicine, Health, & Society | 1 |
| Medicine, Health, & Society; Spanish | 1 |
| Medicine, Health, and Society; Spanish | 1 |
| Middle East and Islamic Studies | 1 |
| Music | 4 |
| Music Performance, Strings Track (Violin) | 1 |
| Music; Pre-Medical | 1 |
| Musical Arts | 1 |
| Narratives of Health: Global Perspectives | 1 |
| Nursing | 14 |
| One Health | 1 |
| Philosophy | 14 |
| Philosophy of Science | 1 |
| Philosophy-Neuroscience-Psychology; Foreign Language: Spanish | 1 |
| Philosophy; Classics | 1 |
| Physician Assistant | 1 |
| Political Science | 10 |
| Political Science; Communications | 1 |
| Political Science; Environmental Studies | 1 |
| Political Science; History | 1 |
| Politics; Business Administration | 1 |
| Pre-Health; Family Studies & Human Development | 1 |
| Pre-Professional | 1 |
| Public Policy | 3 |
| Public Policy (Global Public Health conc.) | 1 |
| Religion | 2 |
| School of Public and International Affairs | 1 |
| Science & Technology Studies | 1 |
| Science Business | 2 |
| Science-Business | 1 |
| Science, Technology & Society | 1 |
| Science, Technology, and International Affairs | 1 |
| Social Studies | 1 |
| Spanish | 4 |
| Stage Management | 1 |
| Studio Art | 1 |
| the study of religion | 1 |
| Theatre | 1 |
| Theatre Arts (and Cinema) | 1 |
| University Scholar | 1 |
| Violin Performance | 1 |
| Women's and Gender Studies | 1 |
| Women's, Gender, and Sexuality Studies; History | 1 |
| Accounting; Finance | 2 |

| **Major combination** | **Number reporting this major** |
| --- | --- |
| "Biochemistry (B.S.)"; "Liberal Studies: Medical Scholar (B.A.)" | 1 |
| African-American Studies; Psychology | 1 |
| Anthropology; Black Studies | 1 |
| Anthropology; Molecular Biology; Interdisciplinary Studies | 1 |
| Anthropology: Global Health and Environment; Foreign Language: French | 1 |
| Anthropology: Global Health and the Environment; Healthcare Management | 1 |
| Art History; Biology | 1 |
| Baylor Business Fellows; Economics | 1 |
| Biochemistry (Molecular Biology); Foreign Language (French) | 1 |
| Biochemistry & Biophysics; Art and the History of Art | 1 |
| Biochemistry & Cell Biology; Business Psychology | 1 |
| Biochemistry and Molecular Biology; Biology; Classics | 1 |
| Biochemistry and Molecular Biology; Religion and Health Care | 1 |
| Biochemistry and Molecular Biology; Spanish | 2 |
| Biochemistry, Cell & Molecular Biology; Spanish | 1 |
| Biochemistry; Biology; Sociology | 1 |
| Biochemistry; Biology; Women's Studies | 1 |
| Biochemistry; Biophysics; Hispanic Studies | 1 |
| Biochemistry; Chinese Language and Cultural Studies | 1 |
| Biochemistry; Dance | 2 |
| Biochemistry; Finance | 1 |
| Biochemistry; French | 1 |
| Biochemistry; Hispanic Studies | 1 |
| Biochemistry; Hispanic Studies | 1 |
| Biochemistry; History | 1 |
| Biochemistry; International Studies | 1 |
| Biochemistry; Literature | 1 |
| Biochemistry; Medicine, Health, and Society | 1 |
| Biochemistry; Music Performance-Harp | 1 |
| Biochemistry; Political Science | 1 |
| Biochemistry; Spanish | 2 |
| Biochemistry; Spanish, Supplementary | 1 |
| Biochemistry; Women's, Gender, and Sexuality Studies | 1 |
| Biological Science Education; Biology, Health, and Society | 1 |
| Biological Sciences; Classics | 2 |
| Biological Sciences; French and Francophone Studies | 1 |
| Biological Sciences; Music | 1 |
| Biological Sciences; Near Eastern Studies | 1 |
| Biological Sciences; Polish, Russian & CEE Studies (Russian Studies) | 1 |
| Biological Sciences; Theology (Supplemental Major) | 1 |
| Biological Sciences: General Biology; Spanish Language, Literature and Culture | 1 |
| Biological Sciences: General; English: British and American Literature | 1 |
| Biological Sciences: Physiology and Neurobiology; Philosophy, Politics, and Economics | 1 |
| Biology; American Studies | 1 |
| Biology; Archaeology | 1 |
| Biology; Art History | 1 |
| Biology; Business | 1 |
| Biology; Business Management | 1 |
| Biology; Community Health | 2 |
| Biology; English | 2 |
| Biology; English and Comparative Literature | 1 |
| Biology; Entrepreneurship | 1 |
| Biology; Finance | 1 |
| Biology; Finance; Healthcare Management | 1 |
| Biology; Fine Arts | 1 |
| Biology; Foreign Language - Spanish | 1 |
| Biology; Foreign Language; Honors Program | 1 |
| Biology; French | 5 |
| Biology; French Studies | 1 |
| Biology; Greek and Latin Studies | 1 |
| Biology; Health and Societies | 1 |
| Biology; Health Policy and Management | 1 |
| Biology; Health Studies | 1 |
| Biology; Healthcare Management | 1 |
| Biology; Interdisciplinary Studies | 1 |
| Biology; International Relations | 1 |
| Biology; Latin American, Latinx, and Caribbean Studies | 1 |
| Biology; Music (Piano) and an Outside Field (Mol. Life Sci); Neuroscience | 1 |
| Biology; Philosophy | 2 |
| Biology; Political Science (American) | 1 |
| Biology; Public Policy | 1 |
| Biology; Russian Language and Literature | 1 |
| Biology; Sociology - American Studies | 1 |
| Biology; Spanish | 9 |
| Biology; Spanish Studies (Secondary Major) | 1 |
| Biology; Studio Arts | 1 |
| Biology; Theatre Arts (and Cinema) | 1 |
| Biology; Theatre: Musical Theatre Track | 1 |
| Biology; Theology | 1 |
| Biology: Neuroscience; Chinese Language and Culture | 1 |
| Biology: Neuroscience; Japanese Language and Culture | 1 |
| Biomedical Engineering; Humanities | 1 |
| Biomedical Engineering; Religion | 1 |
| Biomedical Science; Healthcare Management | 1 |
| Biomedical Science; International Studies | 1 |
| Biomedical Science; Modern Languages German Emphasis | 1 |
| Biomedical Science; Music | 1 |
| Biomedical Science; Spanish | 1 |
| Biomedical Sciences; Spanish for the Health Professions | 1 |
| Biomolecular Science; Middle Eastern and North African Studies | 1 |
| Biopsychology, Cognition, and Neuroscience; Biology, Health, and Society | 1 |
| Biopsychology; French | 1 |
| Biosciences (Concentration in Biochemistry); Classical Studies | 1 |
| Biosciences: Cell Biology and Genetics; History | 1 |
| Business Fellows; Economics | 1 |
| Cell and Molecular Biology; Honors Arts and Letters | 1 |
| Chemical Biology ; Chinese | 1 |
| Chemistry, Emphasis in Biochemistry; Music, Emphasis in Outside Area-Instrumental | 1 |
| Chemistry; Art | 1 |
| Chemistry; Art History | 1 |
| Chemistry; English | 1 |
| Chemistry; Foreign Language | 1 |
| Chemistry; German | 1 |
| Chemistry; Global Studies | 1 |
| Chemistry; Hispanic Studies | 2 |
| Chemistry; Latin American, Latino, and Iberian Studies | 1 |
| Chemistry; Medicine, Health, and Society | 1 |
| Chemistry; Music | 1 |
| Chemistry; Philosophy | 4 |
| Chemistry; Piano Performance | 1 |
| Chemistry; Religious Studies | 1 |
| Chemistry; Spanish | 4 |
| Classics; Cell and Systems Biology | 1 |
| Classics; Molecular & Cellular Biology; Medicine, Science, and the Humanities | 1 |
| Classics; Neuroscience | 1 |
| Cognitive Neuroscience; Spanish | 1 |
| Cognitive Science; Bioethics and Medical Humanities | 1 |
| Community Health; Spanish Major in Literary Studies | 1 |
| Computation and Neural Systems; English | 1 |
| Computational Biology; East Asian Language & Civilization | 1 |
| Computer Science; Biology; Classics | 1 |
| Computer Science; Philosophy | 1 |
| Economics; Pre-Health Studies (Supp.); Spanish (Supp.) | 1 |
| Ecosystem Science & Policy | 1 |
| English (Creative Writing conc.; Middle East mod.); Neuroscience with Honors | 1 |
| English; Biological Sciences | 1 |
| English; Molecular, Cellular, Dev Bio (Int) | 1 |
| English; Neuroscience | 2 |
| Finance; Computational Biology; Mathematics | 1 |
| Finance; Operations Management | 1 |
| Foreign Language; Biology | 2 |
| French and Francophone Studies; Chemistry | 1 |
| Gender, Sexuality, and Feminist Studies; Global Health | 1 |
| Global Health and Environment; Women, Gender and Sexuality Studies | 1 |
| Health and Disease; Immunology | 1 |
| Health and Human Biology; Education Studies | 1 |
| Health Promotion & Disease Prevention | 1 |
| Health Sciences; Social Policy Analysis | 1 |
| History of Science, Medicine & Public Health | 1 |
| History of Science, Medicine, and Public Health; Molecular, Cellular, and Developmental Biology | 1 |
| History of Science; Chemistry | 1 |
| History; Anthropology | 1 |
| History; Biology | 1 |
| History; Biosciences | 1 |
| History; Computational and Systems Biology | 1 |
| History; Global Health | 1 |
| HSSP: Health, Science, Society and Policy; Biology | 1 |
| Human and Organizational Development; Medicine, Health, and Society | 1 |
| Human Biology & Society | 1 |
| Human Biology Health & Society | 1 |
| Human Biology; Educational Psychology | 1 |
| Human Biology; Ethnic Studies | 1 |
| Human Biology; French | 1 |
| Human Evolutionary Biology; Slavic Languages and Literatures | 1 |
| Human Physiology; Spanish | 1 |
| Integrative Physiology; Spanish Language and Literature | 1 |
| Interdisciplinary - Computer Science; Music | 1 |
| International Studies, Global Environment & Health; Middle Eastern and North African Studies | 1 |
| International Studies; Medical Anthropology | 1 |
| Kinesiology ; Spanish | 1 |
| Legal Studies; Biology | 1 |
| Linguistics; Psychology: Behavioral and Cognitive Neuroscience | 1 |
| Mathematics; Asian Studies | 1 |
| Medical Sciences; Spanish | 1 |
| Medicine, Health, and Society; Computer Science | 1 |
| Medicine, Health, and Society; Neuroscience | 2 |
| Medicine, Health, and Society; Philosophy | 1 |
| Medieval Cultures; Biology | 1 |
| Microbial Biology (Microbio); Spanish | 1 |
| Microbiology (Medical); Philosophy (Science, Nature, and Mind) | 1 |
| Microbiology and Cell Science; Music Performance (Euphonium) | 1 |
| Microbiology; English | 1 |
| Microbiology; Music | 1 |
| Microbiology; Spanish | 1 |
| Molecular and Cell Biology- Immunology ; Business Administration | 1 |
| Molecular and Cell Biology; Legal Studies | 1 |
| Molecular and Cell Biology; Middle Eastern Language and Culture | 1 |
| Molecular and Cellular Biology; Architecture and the Built Environment | 1 |
| Molecular and Cellular Biology; French | 1 |
| Molecular and Cellular Biology; Medicine, Health, & Society | 1 |
| Molecular and Cellular Biology; Medicine, Health, and Society | 2 |
| Molecular Biology; East Asian Studies | 1 |
| Molecular Biology; Foreign Language | 1 |
| Molecular Biology; Medicine, Health, and Society | 1 |
| Molecular, Cell, and Developmental Biology; Communication | 1 |
| Molecular, Cellular and Developmental Biology; Spanish | 1 |
| Molecular, Cellular, and Developmental Biology; Philosophy | 1 |
| Molecular, Cellular, Developmental Biology; Global Affairs | 1 |
| Music - Performance Concentration; Chemistry | 1 |
| Music; Anthropology | 1 |
| Music; Classics; Accounting; Chemistry; Human Biology | 1 |
| Music; Neurobiology, Physiology, and Behavior | 1 |
| Music; Neuroscience | 1 |
| Neurobiology & Music Composition | 1 |
| Neuroscience and Behavior; Theology | 1 |
| Neuroscience and Behavioral Biology (NBB); Music Composition | 1 |
| Neuroscience and Behavioral Biology; Religion | 1 |
| Neuroscience; Biology; French and Francophone Studies | 1 |
| Neuroscience; Classics | 3 |
| Neuroscience; Dance | 1 |
| Neuroscience; Fine Arts | 1 |
| Neuroscience; Flute Performance | 1 |
| Neuroscience; Gender and Health | 1 |
| Neuroscience; Gender Studies | 1 |
| Neuroscience; German | 1 |
| Neuroscience; Latin American Studies | 1 |
| Neuroscience; Literature | 1 |
| Neuroscience; Medicine, Health, and Society | 10 |
| Neuroscience; Medicine, Science and the Humanities | 1 |
| Neuroscience; Music | 1 |
| Neuroscience; Philosophy | 5 |
| Neuroscience; Religious Studies | 1 |
| Neuroscience; Spanish | 7 |
| Neuroscience; Women's Studies | 1 |
| Nutrition Science and Research; Asian & Middle Eastern Studies: Chinese | 1 |
| Nutritional Science; Art History | 1 |
| Philosophy; Biology | 2 |
| Philosophy; Natural Sciences | 1 |
| Philosophy; Neuroscience | 1 |
| Philosophy; Neuroscience and Behavior | 1 |
| Philosophy; Psychology | 1 |
| Physics in Medicine; Chinese | 1 |
| Physics; Music | 1 |
| Physics; Secondary Education | 1 |
| Physiology and Neurobiology; History | 1 |
| Planning; Biology | 1 |
| Political Science; Behavioral Neuroscience | 1 |
| Political Science; Biological Sciences | 1 |
| Political Science; Physiology & Medical Sciences | 1 |
| Pre-Medical; Foreign Language: Spanish; Neuroscience | 1 |
| Psychological & Brain Sciences (PBS); Women, Gender, and Sexuality Studies (WGSS) | 1 |
| Psychology; Biology; German | 1 |
| Psychology; English | 1 |
| Psychology; Flute Performance | 1 |
| Psychology; History | 1 |
| Psychology; Medicine, Health, and Society | 1 |
| Psychology; Religion | 1 |
| Psychology; Religious Studies | 1 |
| Public Health Studies; International Studies | 1 |
| Religion; Chemistry | 1 |
| Religious Studies ; Biology | 1 |
| Social Relations and Policy; History, Philosophy, and Sociology of Science | 1 |
| Sociology; Biology, Health, and Society | 1 |
| Sociology; International Studies | 1 |
| Spanish; Molecular and Cellular Biology | 1 |
| Spanish; Neuroscience | 1 |
| Spanish; Philosophy | 1 |
| Spanish; Sociology (Honors) | 1 |
| Sports Medicine & Exercise Physiology; Spanish & Portuguese | 1 |
| Sports Medicine and Exercise Physiology; Linguistics; Biosciences | 1 |
| Studio Art; Chemistry | 1 |
| Women's Studies; Biology | 1 |

| **Major** | **Number reporting this major** |
| --- | --- |
| General Studies | 1 |
| Honors Program | 2 |
| Individualized Major | 1 |
| Interdisciplinary Studies | 5 |
| No Major | 26 |
| University Scholar | 1 |
| University Scholars | 3 |
| (blank) | 38 |
